# A transdiagnostic group exercise intervention for mental health outpatients in Germany (ImPuls): results of a pragmatic, multi-site, block-randomised, phase 3 controlled trial

**DOI:** 10.1101/2023.12.16.23300028

**Authors:** Sebastian Wolf, Britta Seiffer, Johanna-Marie Zeibig, Anna Katharina Frei, Thomas Studnitz, Jana Welkerling, Edith Meinzinger, Leonie Louisa Bauer, Julia Baur, Stephanie Rosenstiel, David Victor Fiedler, Florian Helmhold, Andreas Ray, Eva Herzog, Keisuke Takano, Tristan Nakagawa, Mia Maria Günak, Saskia Kropp, Stefan Peters, Anna Lena Flagmeier, Lena Zwanzleitner, Leonie Sundmacher, Ander Ramos-Murguialday, Martin Hautzinger, Gorden Sudeck, Thomas Ehring

## Abstract

**Background:** This study aimed to assess the efficacy of ImPuls, a transdiagnostic group exercise intervention, plus treatment as usual (TAU) compared to TAU alone in outpatients with various mental disorders.

**Methods:** In this pragmatic, two-arm, multi-site randomised controlled trial in Germany, 10 outpatient rehabilitative and medical care facilities were involved as study sites. Participants were outpatients diagnosed according to ICD-10 with one or more of the following disorders based on structured clinical interviews: major depression, primary insomnia, PTSD, panic disorder, or agoraphobia. Blocks of six participants were randomly allocated to ImPuls plus TAU or TAU alone, stratified by study site. TAU was representative of typical outpatient health care in Germany, allowing patients access to any standard treatments. The primary outcome was global symptom severity at 6 and 12 months after randomisation, measured using self-report on the Brief Symptom Inventory (BSI-18) and analysed in the intention-to-treat sample. Safety was assessed in all participants. The trial was registered with the German Clinical Trials Register (ID: DRKS00024152, 05/02/2021).

**Findings:** Of the 400 eligible participants, 284 (71%) self-identified as female; mean age was 42·20 years (SD 13·23; range 19–65). 287 (71·75%) participants met the criteria for depression, 81 (20·25%) for primary insomnia, 37 (9·25%) for agoraphobia, 46 (11·50%) for panic disorder, and 72 (18%) for PTSD. 199 participants were allocated to the intervention and 201 to the control group. 38 (19·10%) participants did not receive the minimum ImPuls intervention dose. ImPuls plus TAU demonstrated superior efficacy to TAU alone in reducing global symptom severity, with an adjusted difference on BSI-18 of 4·11 (95% CI 1·74 to 6·48; *d*=0·35 [0·14–0·56]; p=0·001) at 6 months and 3·29 (95% CI 0·86–5·72; *d*=0·28 [0·07–0·50]; p=0·008) at 12 months.

**Interpretation:** ImPuls is an efficacious transdiagnostic adjunctive treatment in outpatient mental health care.

**Research in context:** *Evidence before this study:* There is strong evidence that exercise interventions are efficacious in reducing the symptoms of a range of highly prevalent mental disorders, including depression, insomnia, agoraphobia, panic disorder, and post-traumatic stress disorder (PTSD). Most research to date, however, has focused on disorder-specific outcomes and interventions in patient samples with specific mental disorders. In contrast, there is a lack of evidence on transdiagnostic exercise interventions and their effects on global symptom severity in samples of patients with various mental disorders. Furthermore, evidence on the disorder-specific effects of exercise interventions is mostly based on studies without long-term follow-up assessments and with small sample sizes, making it difficult to draw strong conclusions about their long-term efficacy. Before conducting the present study, we searched PubMed in March 2020 without date or language restrictions using the following search terms: ((exercise[Title/Abstract]) OR (physical activity[Title/Abstract])) AND ((transdiagnostic[Title/Abstract])) AND ((intervention) OR (treatment)) AND ((depression) OR (anxiety) OR (panic) OR (agoraphobia) OR (insomnia) OR (PTSD)). Our search had no date or language restrictions, and it used the filter for randomised controlled trials. Three relevant papers were identified: two consisted of study protocols published in 2015 and 2016, and one from 2017 reported the results of a clinical trial. The study protocols described trials designed to evaluate the efficacy of transdiagnostic interventions in which physical activity constituted only one component. The clinical trial investigated the effects of exercise and strength training on disorder-specific and transdiagnostic outcomes, albeit only in patients with anxiety-related disorders.

*Added value:* Our large randomised controlled trial assessed the efficacy of a group exercise intervention (ImPuls) plus treatment as usual (TAU) compared to TAU alone in reducing global transdiagnostic symptom severity in a real-world outpatient context for adult patients diagnosed with depression, insomnia, agoraphobia, panic disorder, or PTSD. Of the 400 patients successfully recruited to the study and randomised to the intervention or control group, 77% were already receiving a standard outpatient treatment (pharmacotherapy or psychological treatment) at baseline. We found larger improvements in global symptom severity in the intervention group compared to control at 6 months after baseline, with persistent benefits seen at the 12-month follow-up assessment. An evaluation of reliable clinical change showed that the effects were clinically meaningful both at 6 and 12 months. We also found indications of benefits for disorder-specific mental health symptoms, including those of depression, panic disorder, general anxiety, and PTSD. These clinical effects were maintained for depression, general anxiety, and panic disorder symptoms up to the 12-month assessment. Mean self-reported exercise increased in the intervention group from 17 weekly minutes on average to the intended dose of more than 90 weekly minutes at 6 months and dropped to 69 minutes at 12 months. Increases in self-reported exercise partially explained the treatment effects for participants who adhered to the minimum intervention dose. After concluding this study in September 2023, we updated our literature search. Apart from our own feasibility study, however, we found no additional studies specifically evaluating the efficacy of exercise on global symptom severity in a sample of patients diagnosed with depression, insomnia, anxiety disorders, or PTSD, underscoring the novelty of our study. Our results provide strong evidence that exercise therapy is a feasible and efficacious transdiagnostic adjunctive treatment in real-world mental health care contexts for outpatients with various mental disorders.

*Implications of all the available evidence:* Strong evidence that exercise is efficacious in treating specific mental disorders, as well as new evidence from our study that exercise has transdiagnostic effects when combined with TAU, suggests that exercise therapy should be used in outpatient mental health care (a) as an alternative to standard treatment or (b) as an adjunctive treatment for disorders such as depression, insomnia, panic disorder, agoraphobia, and PTSD. Transdiagnostic group exercise interventions hold great promise because they allow for the simultaneous treatment of multiple patients. By optimizing the use of healthcare resources and potentially reducing waiting times for treatment, these interventions could ameliorate the existing disparity in care provision between the many individuals in need of evidence-based treatment and the few who are actually receiving it.

## Introduction

Globally, mental disorders are a major health concern, accounting for 4·9% of all disability-adjusted life years (DALYs) and ranking as the seventh leading cause of all DALYs.^1,2^ Depression, insomnia, anxiety, and stress-related disorders are some of the most prevalent mental disorders worldwide. They are frequently comorbid and share aetiological and maintenance factors.^3^

Although evidence-based interventions exist, several serious challenges remain. First, a substantial subgroup of patients does not respond to pharmacotherapy and psychological treatment, with non-response rates ranging from 30–50% across different mental disorders.^4,5^ Second, only a minority of patients worldwide have access to evidence-based standard treatments.^6^ Even in a high-income country like Germany, only 10% of individuals with mental disorders receive evidence-based treatment, and only 2·5% receive psychological treatment, often facing waiting times of up to nine months.^7^ Third, psychological treatment is usually delivered in an individual format, making it inefficient from a public health perspective. Lastly, the use of psychotropic medication can have detrimental effects on physical health.

Exercise at moderate-to-vigorous intensity has demonstrated disorder-specific therapeutic effects in patients with specific mental disorders.^8^ Recent meta-analyses investigating the effects of exercise on depression have found large effects compared to treatment as usual (TAU).^9,10^ Additionally, moderate effects have been reported for exercise in addition to TAU for depression.^11^ In the case of PTSD and insomnia, meta-analyses have found small to moderate effects and large effects, respectively, for exercise interventions compared to passive control groups or TAU.^12,13^ For anxiety and stress-related disorders, combined meta-analytic evidence indicates small to moderate anxiolytic effects.^14^ The efficacy of exercise across these disorders might be explained by its effects on transdiagnostic aetiological and maintenance mechanisms, such as anxiety sensitivity, self-efficacy, and stress reactivity.^15,16^

To date, however, there is a lack of evidence on the efficacy of transdiagnostic group exercise programmes in reducing global symptom severity among patients with various mental disorders. This is unfortunate given that such programmes allow for the simultaneous treatment of multiple patients in heterogeneous groups, optimising the use of healthcare resources. Evaluating these programmes in real-world outpatient settings is crucial to assess their potential as alternatives or adjuncts to standard treatment and help bridge the current treatment gap.

The ImPuls programme, a recent transdiagnostic group exercise intervention, was developed for outpatients with various mental disorders based on current evidence regarding the efficacy of exercise and its long-term maintenance.^18^ Because patients with mental disorders often have difficulties motivating themselves to perform regular exercise, ImPuls explicitly included motivational and volitional elements based on evidence that these can increase exercise maintenance.^17^ A feasibility study of ImPuls demonstrated large immediate and moderate long-term effects of the intervention compared to a passive control group in patients awaiting psychological treatment.^19,20^ In this paper, we report the results of a large-scale, multi-site, pragmatic randomised controlled trial that aimed to investigate the long-term efficacy of ImPuls plus TAU compared to TAU alone in a real-world outpatient mental health care setting in Germany. We tested the following preregistered hypotheses^1^:

1. Participants in the intervention group receiving ImPuls plus TAU will show reduced global symptom severity and more instances of clinically significant change at 6 months and 12 months compared to a control group receiving TAU only.

2a. The intervention will lead to significantly higher volumes of self-reported exercise and moderate-to-vigorous intensity physical activity (MVPA) as measured by accelerometry at 6 months and 12 months compared to the control group.

2b. The effect of the intervention in reducing the primary outcome, global symptom severity, from baseline to 6 months and from baseline to 12 months will be mediated by increases in self-reported exercise and MVPA.

3. Participants in the intervention group will show reduced disorder-specific symptoms compared to participants in the control group at 6 months and 12 months.

## Methods

### Study design

We conducted a pragmatic, multi-site, block-randomised, controlled phase 3 trial with two treatment arms (ImPuls plus TAU vs. TAU alone) and three points of assessment (baseline, 6 months, 12 months). The trial was carried out across 10 different outpatient rehabilitative and medical care facilities in south-west Germany (appendix p 56). The study was approved by the local ethics committee (ID: 888/2020B01, 02/11/2020). The study protocol has been published.^21^

### Participants and recruitment

Eligible participants were adults who were 1) aged between 18 and 65 years, 2) insured by one of the two cooperating statutory health insurers, Allgemeine Ortskrankenkasse Baden-Württemberg (AOK BW) and Techniker Krankenkasse (TK), 3) fluent in German, 4) without medical contraindications for exercise, and 5) diagnosed according to ICD-10 with at least one of the following disorders: major depressive disorders of at least moderate severity (F32·1, F32·2, F33·1, F33·2), insomnia (F51·0), agoraphobia (F40·0, F40·01), panic disorder (F41·0), or PTSD (F43·1). Participants were excluded if they had 1) engaged in at least 30 minutes of exercise of at least moderate intensity more than once a week for six weeks within the three months before study diagnosis or 2) a current diagnosis of mental and behavioural disorders caused by psychotropic substances, eating disorders, bipolar disorder, schizophrenia, or acute suicidality (F-codes see appendix p 16). Gender data were based on self-reporting (female, male, other).

Patients were recruited from various settings (appendix pp 38, 58) and screened for somatic contraindications to exercise using the Physical Activity Readiness Questionnaire (PAR-Q). Additionally, patients had to provide a health provider’s referral for ImPuls before the baseline assessment. After obtaining written informed consent from patients to participate in the study, qualified psychologists conducted a structured clinical interview (SCID-5-CV) to confirm eligibility. Once six patients at a study site were found to be eligible, they received online questionnaires through the web-based data management system REDCap. Additionally, participants were given an accelerometer-based physical activity sensor (MOVE 4; movisens GmbH) to wear for seven consecutive days (appendix pp 22–23; 38–40)

### Randomisation and masking

Each group of six eligible patients was randomly assigned to either ImPuls plus TAU or TAU only. The randomisation sequence was generated by an external data manager using a varying-size permuted block design, stratified by study site. Randomisation codes were generated digitally and concealed on a secure system. The research team responsible for data collection and management in direct contact with patients, were blinded to the randomisation sequence. The data analyst remained blinded to treatment allocation until the primary statistical analyses were complete.

### Procedures

After randomisation and before the intervention started, global symptom severity was reassessed to verify that patients met the cut-off criteria for a mental disorder. All measures taken during the baseline assessment were repeated at 6 and 12 months after randomisation. The intervention group received ImPuls plus TAU, whereas the control group received TAU only. TAU consisted of any available standard intervention typically provided in the German outpatient setting. Upon completing all assessments, patients in the control group were compensated with €450.

The design and components of ImPuls are shown in Figure 1 and appendix pp 17–19.^18,20,21^ Exercise therapists were required to have an academic degree or comparable qualification with at least three years of training as physical activity or exercise professionals, along with a specific therapeutic qualification. All therapists also received training in ImPuls (appendix pp 16, 57).

**Figure 1.**
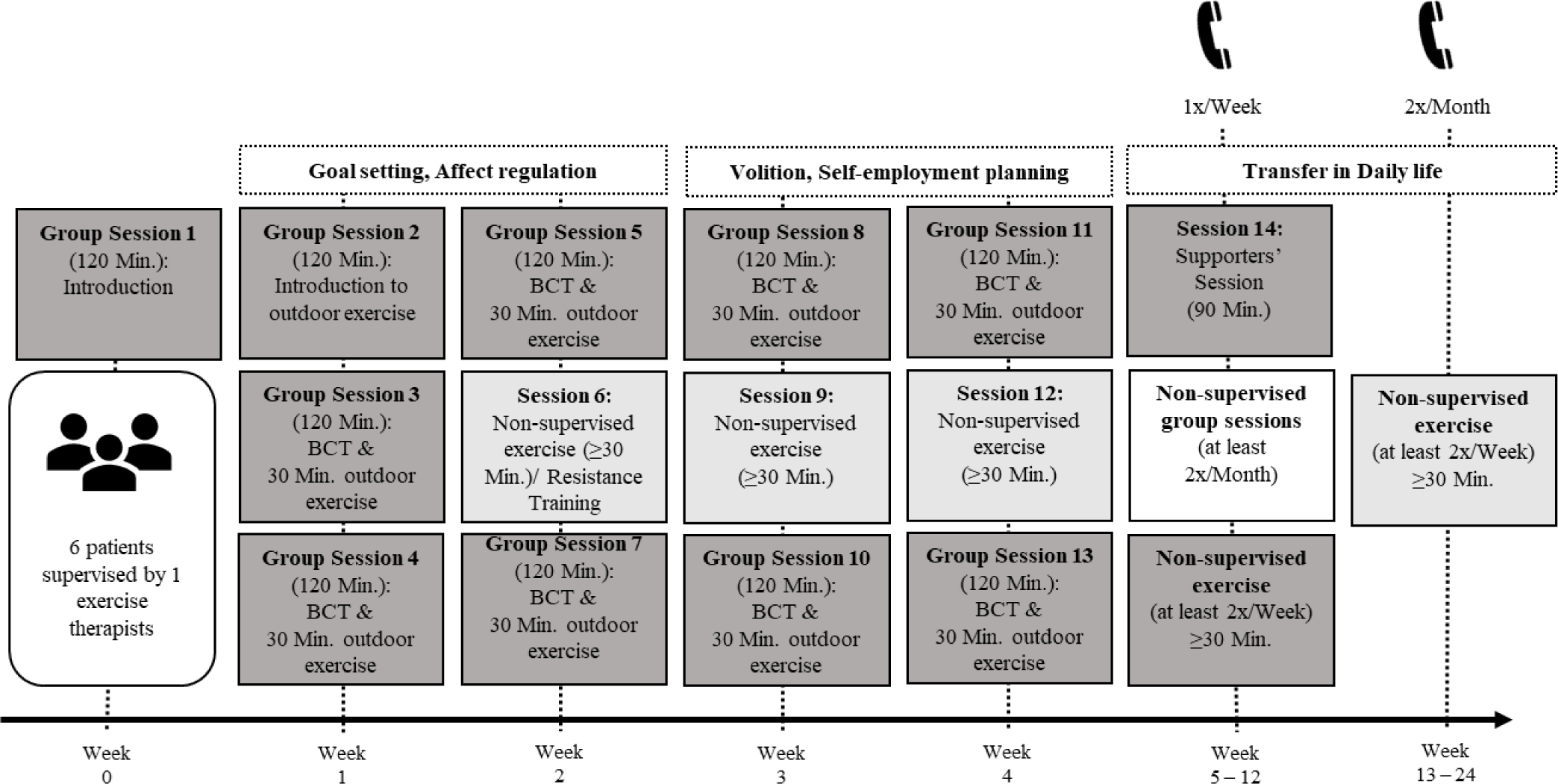
presents the design and components of the ImPuls intervention. The dark grey boxes indicate supervised sessions with group meetings (“Group Session”) and 30 minutes of moderate-to-vigorous aerobic exercise performed as outdoor running along with a supporters’ session in Week 5. Intensity was controlled by a heart rate monitor (SIGMA iD.FREE) combined with a chest strap (SIGMA R1 Bluetooth Duo Comfortex+) and the Borg Rating of Perceived Exertion (RPE) Scale. Moderate-to-vigorous intensity was defined as at least 64% of maximum heart rate, subtracting age from 220 and a self-report of at least 13 points of the RPE Scale. Behavioural change techniques (BCT) such as goal setting or barrier management were integrated to improve motivational and volitional skills for exercise maintenance. These were delivered by exercise therapists in the group sessions and supported by the ImPuls smartphone application. The medium grey boxes depict non-supervised aerobic exercise, allowing patients to choose type of exercises independently based on their own interests. The white box shows optional non-supervised group sessions from week 5 to 24. Telephone calls in combination with the ImPuls smartphone application were used for long-term exercise monitoring. The participants had access to the ImPuls smartphone application during the whole trial, which could be used to track exercise duration and intensity.

Treatment dropout criteria were defined as missing two entire weeks during weeks 1–4. Attendance during the supervised (weeks 0–4) and partially supervised (weeks 5–24) periods was tracked. Study dropouts were defined as intentional discontinuation of the entire study or of entire assessments (appendix pp 20–21; 29–30).

The fidelity of intervention delivery was evaluated through video recordings, with 10% of all sessions randomly selected for evaluation. Adherence to the treatment manual was assessed by two reviewers independently. Details of the assessment process can be found in the appendix (pp 28–29, 59). The overall fidelity score was expected to be ≥90%.

### Outcomes

Global symptom severity served as the primary outcome and was measured using the Global Severity Index (GSI) derived from the validated German version of the Brief Symptom Inventory (BSI-18).^22^ The GSI encompasses ratings of general mental distress across symptom scales for somatisation, depression, and anxiety. Each scale consists of six items, contributing to a total of 18 items in the assessment. Participants rate each item on a 5-point Likert scale (range: 0–4). The total score is calculated for each of the three scales, and the GSI is determined by summing these three scores. Higher scores on the GSI indicate greater levels of distress, with a clinical cut-offset at 12.^22^

Secondary outcomes (appendix pp 23–27) were depressive symptoms assessed with the Patient Health Questionnaire (PHQ-9), non-organic insomnia symptoms assessed with the Insomnia Severity Index (ISI), sleep quality assessed with the global sleep quality score of the Pittsburgh Sleep Quality Index (PSQI), anxiety symptoms assessed with the Generalized Anxiety Disorder Scale (GAD-7), panic disorder and agoraphobia symptoms assessed with the three-item panic subscale of the BSI-18, and symptoms of PTSD were assessed with the PTSD Checklist for DSM-5.

Self-reported exercise was measured in minutes per week using the Exercise Activity Index of the Physical Activity, Exercise, and Sport Questionnaire (BSA). Weekly minutes spent in moderate-to-vigorous physical activity (MVPA; ≥3MET) were measured using accelerometer-based sensors (Move 4, movisens GmbH).

Additional assessments included calculating mean objective exercise intensity, expressed as a percentage of maximum heart rate (HRmax). HRmax is calculated by subtracting the participant’s age from 220. Heart rate data were collected using heart rate monitors (SIGMA iD.FREE). Using the ImPuls smartphone application, perceived exertion was rated by participants with the Borg Rating of Perceived Exertion (RPE) scale and mean session duration (minutes/session) was tracked during the supervised period (weeks 1–4) (appendix pp 17, 30).

Patients’ outcome expectations, motivation, and satisfaction with the intervention, as well as exercise therapists’ motivation and satisfaction with the intervention, were assessed with validated scales (appendix pp 30–36). On these scales, means falling into the upper quarter, the second or third quarters, or the lowest quarter were considered high, moderate, and low scores, respectively.

Adverse events (AE) were assessed at baseline, 6 months, and 12 months. Serious adverse events (SAE) could be reported at any time and were assessed through structured interviews at each assessment point. All SAEs were reported to an independent Data Safety and Monitoring Board (appendix pp 46–49).

### Choice of primary outcome measure

The BSI-18 is a short and resource-efficient version of the Symptom Checklist-90 (SCL-90). The questionnaire showed excellent fit with the composition of our transdiagnostic sample because the GSI derived from it encompasses ratings across depression, anxiety, and somatisation symptoms. In German outpatients with various mental disorders, the GSI has shown good internal consistency (Cronbach’s alpha=0·89). Moreover, it has demonstrated construct validity in patients with affective disorders and anxiety disorders, with correlation coefficients of r=0·71 and r=0·67, respectively.^23^

### Statistical methods

The minimum sample size for the study (N=375) was determined a priori through a power analysis. This analysis was based on the smallest post-treatment effect size of exercise versus TAU/waiting list reported in earlier meta-analyses, which was d=-0·348 for PTSD symptoms.^13^ We assumed a two-sided t-test, alpha level of 0·05, a test power of 80%, an equal cell population, and a dropout rate of 30%. To ensure sufficient statistical power for further predefined analyses, as published in a separate study protocol for the process evaluation of ImPuls, a maximum sample size of N=600 was pre-registered.^24^ Due to recruitment delays caused by the COVID-19 pandemic, the original target of 600 participants was not met. However, we ultimately recruited 400 participants, which means that the study was sufficiently powered for the main analyses.

All analyses adhered to the pre-established statistical analysis plan published before database lock^21^ and were conducted using R (Version 4). Descriptive statistics were calculated for baseline characteristics and outcome measures at 6 and 12 months. The primary and secondary outcomes (Hypotheses 1, 2a, and 3) were analysed using linear mixed models (restricted maximum likelihood estimation; appendix pp 43-44). These models incorporated categorical fixed factors for time (baseline, 6 months, 12 months), groups (ImPuls plus TAU and TAU), and their interaction. The analysis included data from all randomised participants on an intention-to-treat basis, except for one individual who withdrew consent for data analysis (n=399). Missing values were addressed using multilevel multiple imputation. The models included random intercepts to account for between-person and between-site variation and random slopes for time-related between-person variation. The significance level was set at alpha=0·05.

The normality assumption of residuals was checked using QQ plots, and non-normally distributed data were log-transformed. Effect sizes for adjusted group differences at each follow-up time point were calculated, divided by the standard deviation estimated through the mixed models. Standardised within-group effect sizes were computed from adjusted mean within-group changes relative to baseline. A reliable change index based on the Jacobson–Truax method was calculated for the GSI score at 6 months and again for the GSI score at 12 months on the non-imputed dataset (appendix p 44). The Mann–Whitney U test was performed to assess differences in ordinal scores (recovered, improved, unchanged, deteriorated) between groups. Analyses for the primary and secondary outcomes were repeated on the completer sample, defined as those who completed at least two full weeks of the supervised intervention (appendix pp 20–21).

To test Hypothesis 2b, mediation analyses were performed on the simple change scores of GSI (outcome) and of MVPA and self-reported exercise (as the mediators). Two path models were estimated: one for changes from baseline to 6 months and the other for changes from baseline to 12 months. We used full-information maximum likelihood estimation to handle missing observations and bootstrapped standard errors for direct, indirect, and total effects. The same mediation analyses were repeated on the completer sample.

The study was registered with the German Clinical Trial Register (ID: DRKS00024152, 05/02/2021). The progress of the study and its final results were discussed with the Data Safety and Monitoring Board (DSMB).

## Results

Between January 1, 2021, and May 31, 2022, a total of 600 patients provided informed consent and were recruited to the study (Figure 2). Of these patients, 199 were excluded based on the inclusion and exclusion criteria. Additionally, one patient withdrew consent during the baseline assessment. The remaining 400 patients were then randomly assigned to ImPuls plus TAU (n=199) or TAU alone (n=201).

**Figure 2.**
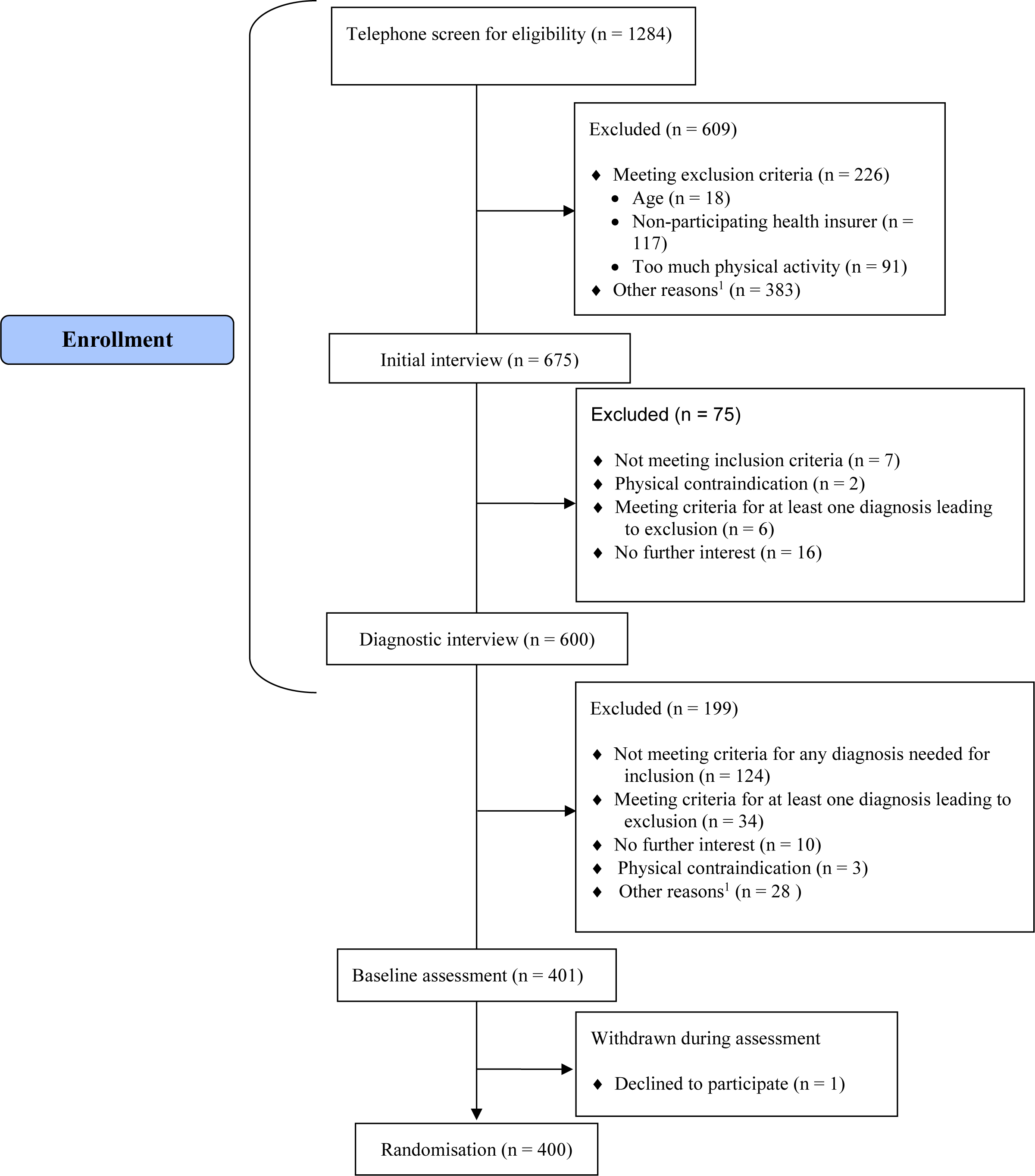
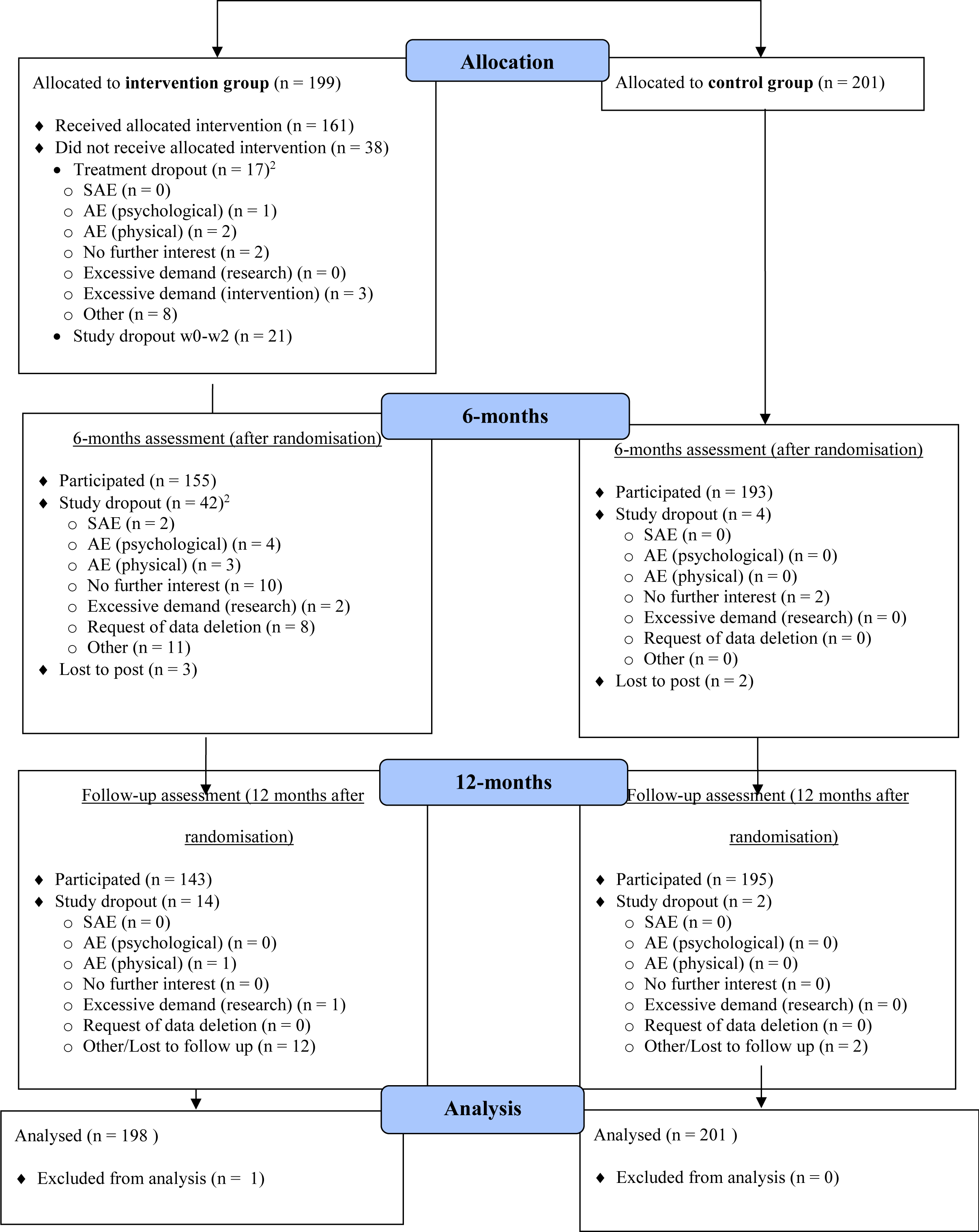
presents the patient flowchart based on CONSORT. ^1^ Other reasons include organisational problems, relocation, no more contact possible, physical constraints, language problems, unknown. ^2^ Multiple answers possible; AE = adverse events; SAE = serious adverse events

Demographic and baseline data for these 400 participants are shown in Table 1 (appendix pp 53–54). Their mean age was 42·20 years (SD 13·23, range 19–65), and 71% reported being female, 26·50% male, and 2·25% of other gender. Regarding diagnoses, 71·75% of participants were diagnosed with depression, 20·25% with primary insomnia, 9·25% with agoraphobia, 11·50% with panic disorder, and 18% with PTSD. Altogether 24·5% of participants additionally had at least one of the other inclusion diagnoses, and 49% had at least one additional psychiatric diagnosis that was not among the inclusion diagnoses.

**Table 1.**
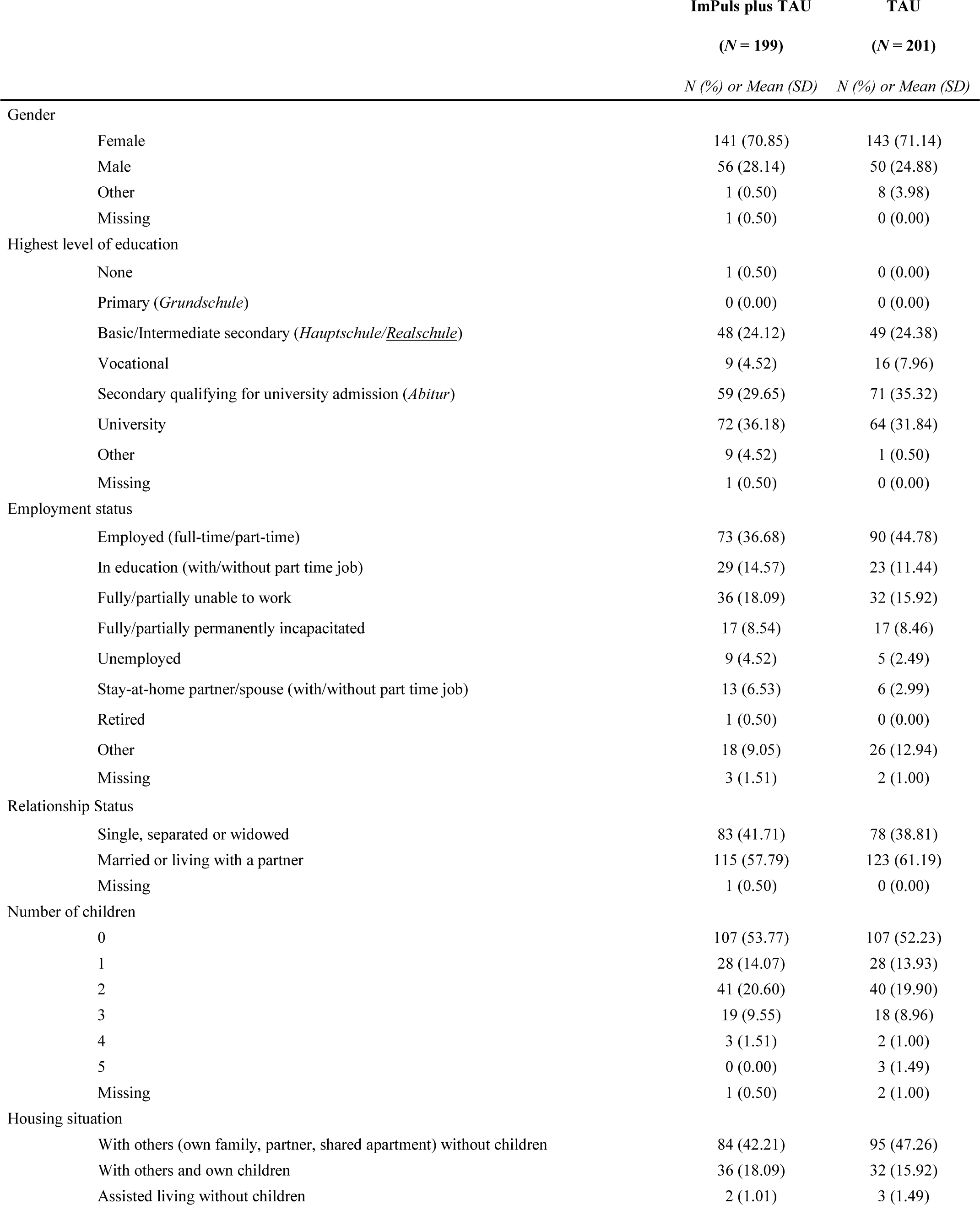
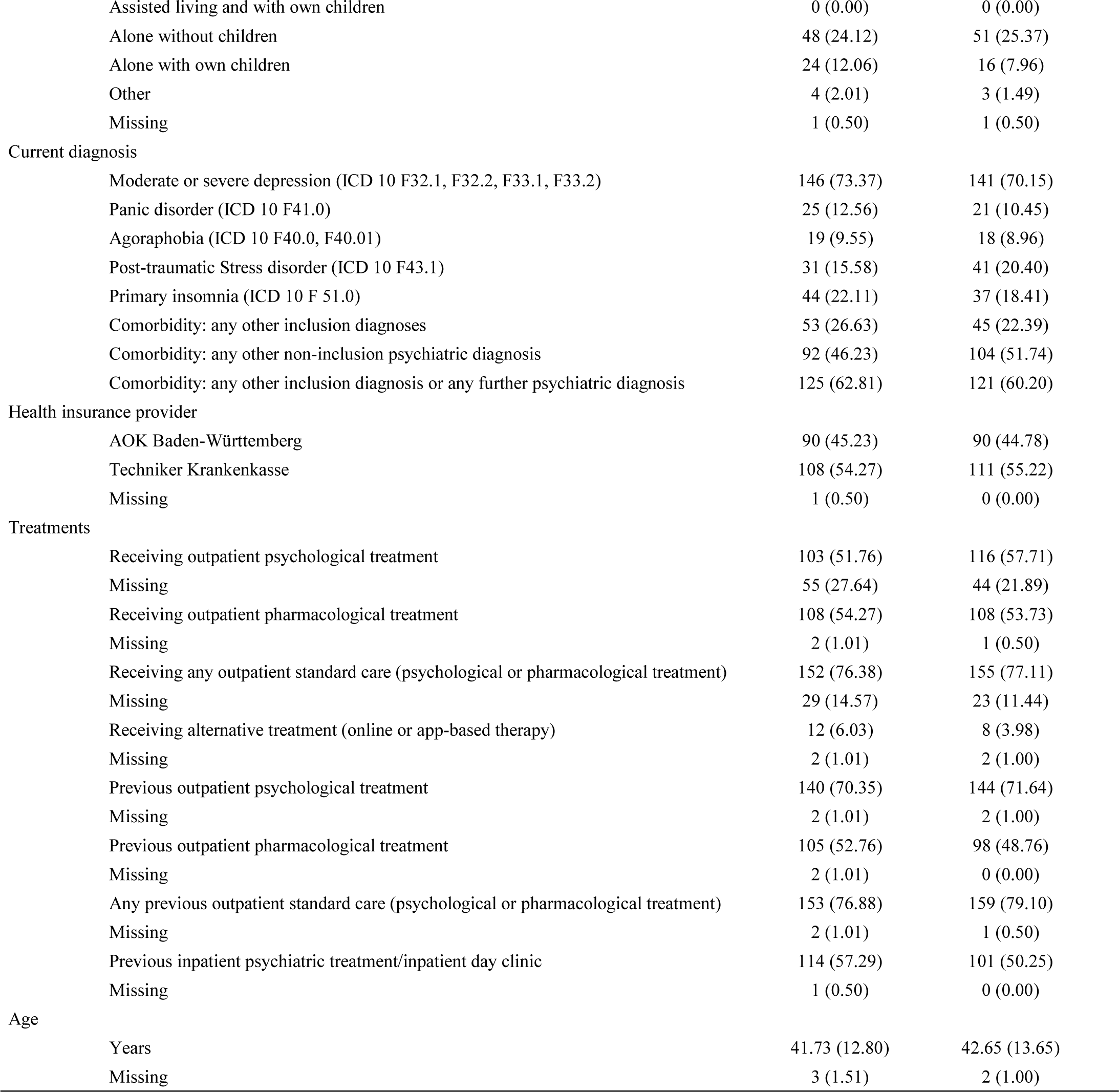
Baseline demographic and clinical characteristics of the intention-to-treat population (*n*=400). All characteristics were reported in N (%), except for age, which was presented as mean (SD). TAU = treatment as usual;

|  | ImPuls plus TAU | TAU |
| --- | --- | --- |
|  | (*N* = 199) | (*N* = 201) |
|  | *N* (%) or Mean (SD) | *N* (%) or Mean (SD) |
| Gender |  |  |
| Female | 141 (70.85) | 143 (71.14) |
| Male | 56 (28.14) | 50 (24.88) |
| Other | 1 (0.50) | 8 (3.98) |
| Missing | 1 (0.50) | 0 (0.00) |
| Highest level of education |  |  |
| None | 1 (0.50) | 0 (0.00) |
| Primary (*Grundschule*) | 0 (0.00) | 0 (0.00) |
| Basic/Intermediate secondary (*Hauptschule/Realschule*) | 48 (24.12) | 49 (24.38) |
| Vocational | 9 (4.52) | 16 (7.96) |
| Secondary qualifying for university admission (*Abitur*) | 59 (29.65) | 71 (35.32) |
| University | 72 (36.18) | 64 (31.84) |
| Other | 9 (4.52) | 1 (0.50) |
| Missing | 1 (0.50) | 0 (0.00) |
| Employment status |  |  |
| Employed (full-time/part-time) | 73 (36.68) | 90 (44.78) |
| In education (with/without part time job) | 29 (14.57) | 23 (11.44) |
| Fully/partially unable to work | 36 (18.09) | 32 (15.92) |
| Fully/partially permanently incapacitated | 17 (8.54) | 17 (8.46) |
| Unemployed | 9 (4.52) | 5 (2.49) |
| Stay-at-home partner/spouse (with/without part time job) | 13 (6.53) | 6 (2.99) |
| Retired | 1 (0.50) | 0 (0.00) |
| Other | 18 (9.05) | 26 (12.94) |
| Missing | 3 (1.51) | 2 (1.00) |
| Relationship Status |  |  |
| Single, separated or widowed | 83 (41.71) | 78 (38.81) |
| Married or living with a partner | 115 (57.79) | 123 (61.19) |
| Missing | 1 (0.50) | 0 (0.00) |
| Number of children |  |  |
| 0 | 107 (53.77) | 107 (52.23) |
| 1 | 28 (14.07) | 28 (13.93) |
| 2 | 41 (20.60) | 40 (19.90) |
| 3 | 19 (9.55) | 18 (8.96) |
| 4 | 3 (1.51) | 2 (1.00) |
| 5 | 0 (0.00) | 3 (1.49) |
| Missing | 1 (0.50) | 2 (1.00) |
| Housing situation |  |  |
| With others (own family, partner, shared apartment) without children | 84 (42.21) | 95 (47.26) |
| With others and own children | 36 (18.09) | 32 (15.92) |
| Assisted living without children | 2 (1.01) | 3 (1.49) |
| Assisted living and with own children | 0 (0.00) | 0 (0.00) |
| Alone without children | 48 (24.12) | 51 (25.37) |
| Alone with own children | 24 (12.06) | 16 (7.96) |
| Other | 4 (2.01) | 3 (1.49) |
| Missing | 1 (0.50) | 1 (0.50) |
| Current diagnosis |  |  |
| Moderate or severe depression (ICD 10 F32.1, F32.2, F33.1, F33.2) | 146 (73.37) | 141 (70.15) |
| Panic disorder (ICD 10 F41.0) | 25 (12.56) | 21 (10.45) |
| Agoraphobia (ICD 10 F40.0, F40.01) | 19 (9.55) | 18 (8.96) |
| Post-traumatic Stress disorder (ICD 10 F43.1) | 31 (15.58) | 41 (20.40) |
| Primary insomnia (ICD 10 F 51.0) | 44 (22.11) | 37 (18.41) |
| Comorbidity: any other inclusion diagnoses | 53 (26.63) | 45 (22.39) |
| Comorbidity: any other non-inclusion psychiatric diagnosis | 92 (46.23) | 104 (51.74) |
| Comorbidity: any other inclusion diagnosis or any further psychiatric diagnosis | 125 (62.81) | 121 (60.20) |
| Health insurance provider |  |  |
| AOK Baden-Württemberg | 90 (45.23) | 90 (44.78) |
| Techniker Krankenkasse | 108 (54.27) | 111 (55.22) |
| Missing | 1 (0.50) | 0 (0.00) |
| Treatments |  |  |
| Receiving outpatient psychological treatment | 103 (51.76) | 116 (57.71) |
| Missing | 55 (27.64) | 44 (21.89) |
| Receiving outpatient pharmacological treatment | 108 (54.27) | 108 (53.73) |
| Missing | 2 (1.01) | 1 (0.50) |
| Receiving any outpatient standard care (psychological or pharmacological treatment) | 152 (76.38) | 155 (77.11) |
| Missing | 29 (14.57) | 23 (11.44) |
| Receiving alternative treatment (online or app-based therapy) | 12 (6.03) | 8 (3.98) |
| Missing | 2 (1.01) | 2 (1.00) |
| Previous outpatient psychological treatment | 140 (70.35) | 144 (71.64) |
| Missing | 2 (1.01) | 2 (1.00) |
| Previous outpatient pharmacological treatment | 105 (52.76) | 98 (48.76) |
| Missing | 2 (1.01) | 0 (0.00) |
| Any previous outpatient standard care (psychological or pharmacological treatment) | 153 (76.88) | 159 (79.10) |
| Missing | 2 (1.01) | 1 (0.50) |
| Previous inpatient psychiatric treatment/inpatient day clinic | 114 (57.29) | 101 (50.25) |
| Missing | 1 (0.50) | 0 (0.00) |
| Age |  |  |
| Years | 41.73 (12.80) | 42.65 (13.65) |
| Missing | 3 (1.51) | 2 (1.00) |

Regarding treatment history, 76·75% of the sample was receiving a standard pharmacological or psychological treatment at baseline. Additionally, 78% reported a previous standard treatment, and 53·75% had a history of inpatient or day clinical treatment. Directly after randomisation, global symptom severity in both groups (mean 20·67 [SD 11·55]) was comparable to the German clinical norm sample (mean 20·23 [SD 12·19]).^25^

Treatment fidelity achieved an overall score of 87%, which was only slightly lower than intended (>90%) and can still be considered high (appendix pp 59–60). Demographic characteristics of the exercise therapists can be found in appendix p 57. Therapist motivation (scale 1–5, mean 4·25 [SD 0·57], n=19), satisfaction with the intervention (scale 1–5, mean 3·39 [SD 0·91], n=15), and expectations of intervention success (scale 1–5, mean 3·88 [SD 0·55], n=19) at baseline were moderate to high. Patient motivation (scale 4–16, mean 14·05 [SD 2·50], n=354) and expectations of intervention success (scale 1–5, mean 3·03 [SD 0·96], n=350) were also moderate to high. Satisfaction with the intervention after the four-week supervised period (scale 6–30, mean 21·96 [SD 3·88], n=156) and after the six-month intervention period (scale 6–30, mean 22·28 [SD 5·11], n=135) was moderate. The overall attendance rate was high at 84%. Mean objective exercise intensity, indexed by percentage of individual maximum heart rate, was 71% ([SD 14%], n=38) and thus aligned with expectations, as did the mean subjective rating of perceived exertion (scale 6–20, mean 13·96 [SD 1·44], n=79) and mean exercise session duration (32·88 minutes [SD 9·19], n=82) averaged across all exercise sessions within the supervised period (appendix p 60).

In total, 61 participants (15·5% out of n=399) dropped out of the study (ImPuls plus TAU: n=55 [27·6%]; TAU: n=6 [3%])). In the ImPuls plus TAU group, 161 patients (80·9% of n=199) completed the minimum intervention dose, indicating a treatment drop-out rate of 19·1%. Data for the primary outcome (GSI) were available for 398 participants at baseline (TAU: n=201; ImPuls plus TAU: n=197), 338 participants at 6 months (TAU: n=190; ImPuls plus TAU: n=148), and 329 participants at 12 months (TAU: n=194; ImPuls plus TAU: n=135). Performing Little’s test on the primary and secondary outcomes led to the rejection of the missing completely at random (MCAR) assumption (X^2^[179]=352, p<0·001). Baseline predictors of attrition in the ImPuls plus TAU group indicated a higher likelihood of study discontinuation among individuals with agoraphobia (ICD-10 F40·00) and those who had received previous psychiatric treatment (appendix pp 66–67).

Descriptive statistics were calculated for the primary and secondary outcome measures at all assessment points, along with the results of the mixed model analyses, and are summarised in Table 2 (appendix pp 61–65). Results for the primary outcome are depicted in Figure 3. In the primary intention-to-treat analysis of 399 participants, ImPuls plus TAU was superior to TAU alone, showing an adjusted difference on the BSI-18 of 4·11 (95% CI 1·74–6·48; *d*=0·35 [0·14–0·56]; p=0·001) at 6 months and 3·29 (95% CI 0·86–5·72; *d*=0·28 [0·07–0·50]; p=0·008; Table 2, Figure 3) at 12 months. In the ImPuls plus TAU group, effect sizes were moderate at both 6 months (*d*=-0·57) and 12 months (*d*=-0·60). Based on the Jacobson–Truax method, a greater number of participants in the intervention group achieved clinically significant changes from baseline to 6 months (W=11551, p=0·002) and from baseline to 12 months (W=11084, p=0·009]; appendix pp 44, 61) compared to TAU alone.

**Figure 3.**
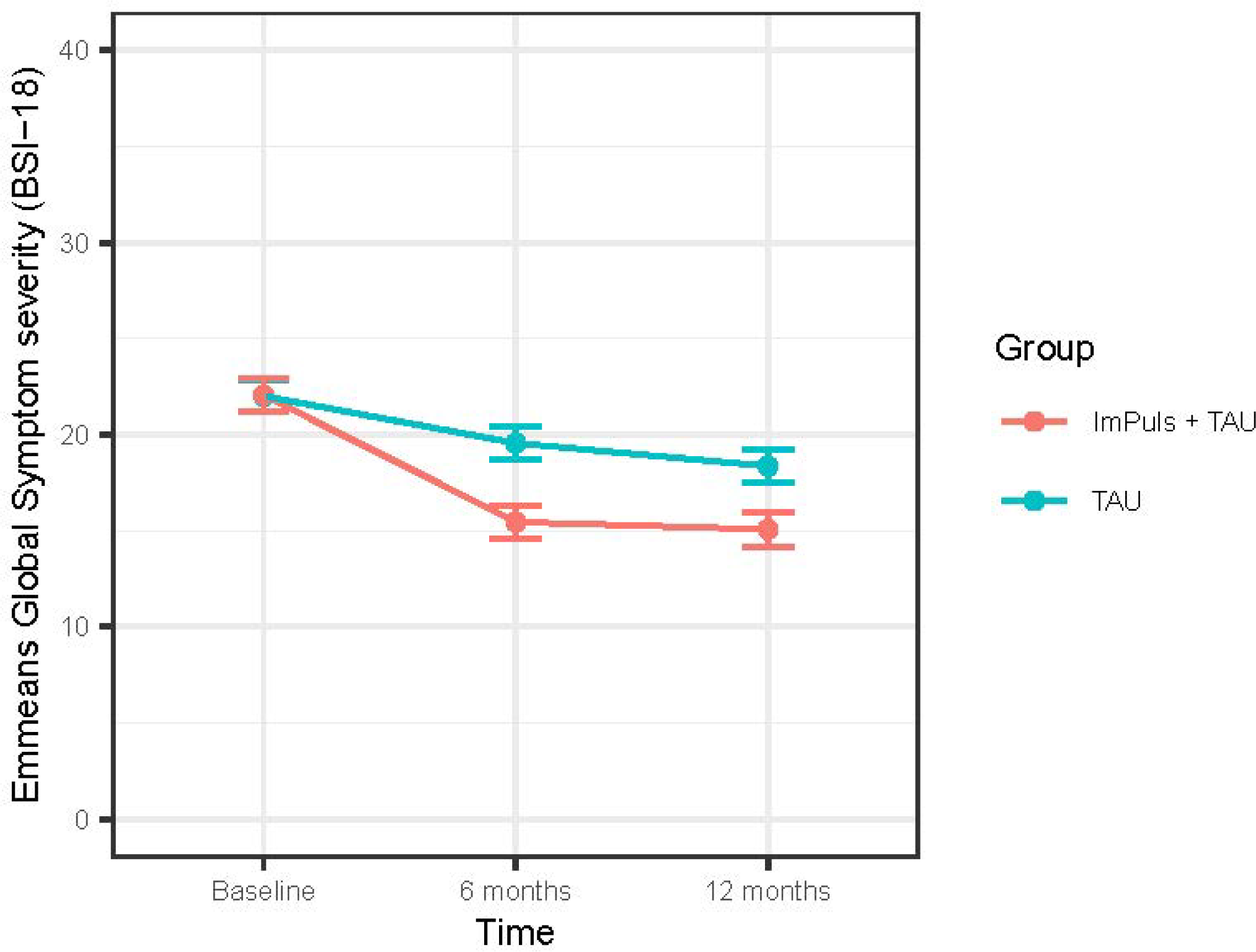
Plot of Emmeans derived from the mixed models on global symptom severity at all measurement points. Baseline global symptom severity was recorded before randomisation. Error bars show standard errors; BSI-18 = Brief Symptom Inventory 18

**Table 2.** Descriptive summaries of primary and secondary outcome measures and results of the mixed models on the ITT sample including unadjusted means, adjusted differences, standardised between and within-group differences for the primary and all secondary outcomes. TAU = treatment as usual; SD = standard deviation; CI = confidence interval; BSI-18 = Brief Symptom Inventory 18; PHQ-9 = Patient Health Questionnaire-9; ISI = Insomnia Severity Index; PSQI = Pittsburgh Sleep Quality Index; GAD-7 = Generalized Anxiety Disorder scale; PCL-5 = Posttraumatic Stress Disorder Checklist 5; MVPA = weekly minutes spent in moderate-to-vigorous physical activity based on metabolic equivalent of task values (≥3 MET) derived from accelerometry sensors worn for 7 consecutive days; BSA = self-reported exercise in minutes per week based on the athletic exercise index of the Physical Activity, Exercise, and Sport Questionnaire; * Adjusted and standardised estimates are based on log-transformed data due to a skewed distribution of raw data; Significant result: p<0.05

|  | Unadjusted mean (SD); N |  | Adjusted difference (95% CI) | p value | Standardised between-group effect size (95% CI) | Standardised within-group effect size (95% CI) |  |
| --- | --- | --- | --- | --- | --- | --- | --- |
|  | TAU plus ImPuls | TAU |  |  |  | TAU plus ImPuls | TAU |
| Global Severity Index (BSI-18) |  |  |  |  |  |  |  |
| Baseline | 22.08 (11.75); 197 | 22.01 (10.55); 201 |  |  |  |  |  |
| 6 months | 14.03 (11.28); 148 | 19.61 (12.36); 190 | 4.108 (1.736 to 6.480) | 0.001 | 0.351 (0.142 to 0.561) | -0.566 (-0.707 to -0.425) | -0.209 (-0.341 to -0.078) |
| 12 months | 12.65 (11.20); 135 | 18.40 (12.45); 194 | 3.291 (0.863 to 5.720) | 0.008 | 0.282 (0.068 to 0.495) | -0.599 (-0.750 to -0.448) | -0.312 (-0.445 to -0.179) |
| Depression (PHQ-9) |  |  |  |  |  |  |  |
| Baseline | 13.76 (5.04); 197 | 13.79 (4.99); 200 |  |  |  |  |  |
| 6 months | 10.33 (5.61); 147 | 12.15 (5.92); 190 | 1.259 (0.109 to 2.408) | 0.032 | 0.228 (0.014 to 0.442) | -0.521 (-0.675 to -0.368) | -0.292 (-0.433 to -0.151) |
| 12 months | 9.07 (5.67); 134 | 11.72 (6.26); 192 | 1.468 (0.237 to 2.700) | 0.020 | 0.266 (0.038 to 0.494) | -0.635 (-0.810 to -0.461) | -0.367 (-0.512 to -0.223) |
| Insomnia (ISI) |  |  |  |  |  |  |  |
| Baseline | 15.14 (5.68); 197 | 14.50 (6.37); 201 |  |  |  |  |  |
| 6 months | 11.51 (5.50); 148 | 13.19 (6.55); 190 | 1.245 (-0.024 to 2.515) | 0.054 | 0.204 (-0.016 to 0.424) | -0.498 (-0.658 to -0.339) | -0.210 (-0.356 to -0.063) |
| 12 months | 10.51 (6.09); 135 | 12.46 (6.26); 194 | 0.955 (-0.331 to 2.242) | 0.145 | 0.157 (-0.065 to 0.379) | -0.575 (-0.742 to -0.408) | -0.334 (-0.485 to -0.183) |
| Sleep Quality (PSQI) |  |  |  |  |  |  |  |
| Baseline | 10.10 (3.74); 182 | 9.93 (3.87); 196 |  |  |  |  |  |
| 6 months | 8.67 (3.81); 125 | 9.02 (3.59); 167 | 0.418 (-0.435 to 1.271) | 0.336 | 0.109 (-0.121 to 0.340) | -0.328 (-0.483 to -0.173) | -0.174 (-0.318 to 0.031) |
| 12 months | 8.27 (3.87); 115 | 9.10 (4.02); 166 | 0.436 (0.302 to -0.394) | 0.302 | 0.114 (-0.111 to 0.338) | -0.353 (-0.525 to -0.182) | -0.194 (-0.332 to -0.057) |
| Anxiety (GAD-7) |  |  |  |  |  |  |  |
| Baseline | 10.54 (4.87); 196 | 10.67 (4.78); 201 |  |  |  |  |  |
| 6 months | 7.49 (4.58); 148 | 9.40 (4.92); 190 | 1.284 (0.268 to 2.300) | 0.013 | 0.261 (0.048 to 0.473) | -0.487 (-0.638 to -0.337) | -0.252 (-0.389 to -0.115) |
| 12 months | 6.56 (4.64); 133 | 8.93 (5.12); 192 | 1.221 (0.142 to 2.299) | 0.027 | 0.248 (0.023 to 0.473) | -0.573 (-0.746 to -0.399) | -0.351 (-0.488 to -0.213) |
| Panic (BSI-18) |  |  |  |  |  |  |  |
| Baseline | 2.81 (2.79); 197 | 2.59 (2.47); 201 |  |  |  |  |  |
| 6 months | 1.38 (2.16); 148 | 2.18 (2.42); 190 | 0.595 (0.096 to 1.094) | 0.019 | 0.245 (0.035 to 0.456) | -0.498 (-0.657 to -0.339) | -0.164 (-0.306 to -0.022) |
| 12 months | 1.16 (2.03); 135 | 2.13 (2.33); 194 | 0.573 (0.075 to 1.071) | 0.024 | 0.236 (0.026 to 0.446) | -0.516 (-0.676 to -0.357) | -0.192 (-0.333 to -0.050) |
| PTSD (PCL-5) |  |  |  |  |  |  |  |
| Baseline | 29.10 (16.59); 196 | 30.08 (14.94); 200 |  |  |  |  |  |
| 6 months | 20.20 (16.37); 148 | 26.27 (17.09); 187 | 3.937 (0.632 to 7.242) | 0.020 | 0.240 (0.032 to 0.447) | -0.410 (-0.540 to -0.281) | -0.231 (-0.354 to -0.109) |
| 12 months | 18.37 (16.22); 133 | 24.15 (16.40); 192 | 3.087 (-0.384 to 6.558) | 0.081 | 0.188 (-0.029 to 0.405) | -0.469 (-0.612 to -0.327) | -0.342 (-0.467 to -0.217) |
| Self-reported exercise (BSA) * |  |  |  |  |  |  |  |
| Baseline | 17.90 (55.54); 196 | 19.67 (84.13); 200 |  |  |  |  |  |
| 6 months | 92.06 (109.58); 145 | 37.77 (79.92); 189 | -1.109 (-1.576 to -0.641) | 0.000 | -0.654 (0.936 to -0.373) | 1.039 (0.804 to 1.275) | 0.397 (0.183 to 0.612) |
| 12 months | 68.51 (107.73); 131 | 54.35 (125.80); 192 | -0.440 (-1.001 to 0.120) | 0.123 | -0.260 (-0.594 to 0.074) | 0.727 (0.437 to 1.017) | 0.479 (0.266 to 0.693) |
| MVPA (Accelerometry data) * |  |  |  |  |  |  |  |
| Baseline | 325.78 (204.41); 172 | 336.93 (233.84); 184 |  |  |  |  |  |
| 6 months | 341.40 (239.38); 114 | 336.75 (234.98); 146 | -0.023 (-0.204 to 0.158) | 0.802 | -0.030 (-0.273 to 0.213) | 0.030 (-0.154 to 0.214) | -0.023 (-0.175 to 0.128) |
| 12 months | 342.33 (226.19); 111 | 357.40 (240.70); 172 | 0.027 (-0.146 to 0.200) | 0.761 | 0.035 (-0.198 to 0.268) | 0.041 (-0.114 to -0.195) | 0.052 (-0.106 to 0.210) |

**Figure 4.**
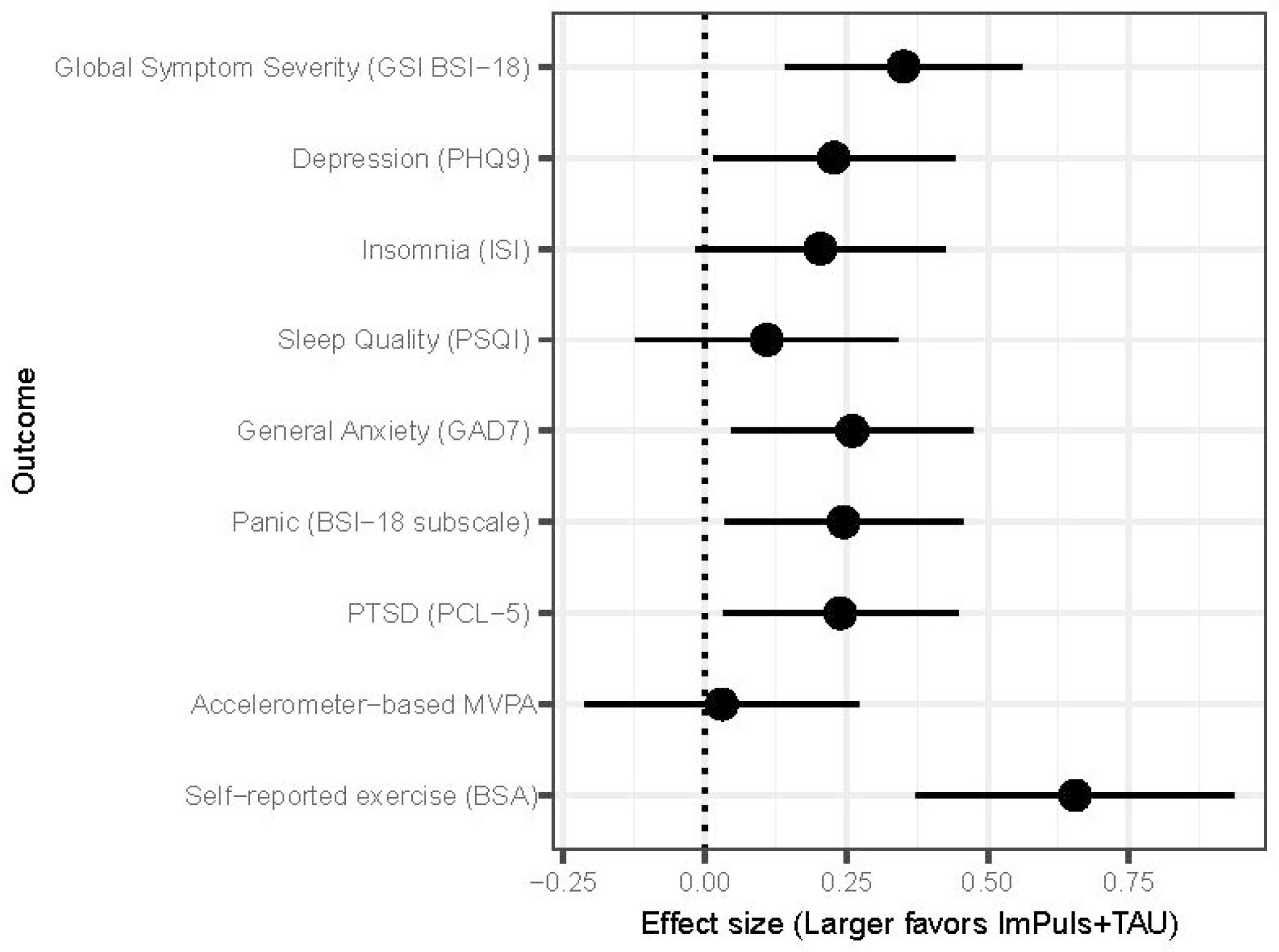
Forest Plot of standardised group differences at the 6-month assessment for the primary and all secondary outcomes between ImPuls plus TAU and TAU, whereby a standardised treatment effect greater than 0 favoured the ImPuls plus TAU group. Error bars show 95% CIs. MVPA and BSA values are based on log-transformed data. All outcomes are based on continuous scales despite MVPA, which indexes weekly minutes spent in moderate-to-vigorous physical activity based on metabolic equivalent of task values (≥3 MET) derived from accelerometry sensors worn for 7 consecutive days. TAU = treatment as usual; CI = confidence interval; BSI-18 = Brief Symptom Inventory 18; PHQ-9 = Patient Health Questionnaire-9; ISI = Insomnia Severity Index; PSQI = Pittsburgh Sleep Quality Index; GAD-7 = Generalized Anxiety Disorder scale; PCL-5 = The Posttraumatic Stress Disorder Checklist 5; MVPA = weekly moderate-to-vigorous physical activity; BSA = Physical Activity, Exercise, and Sport Questionnaire

Across secondary outcomes, ImPuls plus TAU resulted in superior improvements compared to TAU alone in measures of depression, panic, general anxiety, PTSD, and self-reported exercise up to 6 months (Table 2, Figure 3). At 12 months, the superiority of ImPuls plus TAU persisted with regard to symptoms of depression, general anxiety, and panic. The analysis of treatment completers (appendix pp 68–73) showed significant between-group differences, with increased effect sizes compared to the primary analysis at both 6 months (4·67, 95% CI 2·20–7·17; *d*=0·40 [0·18–0·63]; p<0·001) and 12 months (3·84, 95% CI 1·13–6·55; *d*=0·33 [0·09–0·57]; p=0·006). Significant long-term differences between the two groups at the 12-month assessment were seen for depression, general anxiety, panic, and PTSD symptoms.

The results of the mediation analysis indicated only marginally significant mediation effects of changes in self-reported exercise on changes in global symptom severity for both the 6-month and 12-month assessments (appendix pp 65–66). However, in the completer sample, a significant indirect effect was identified on changes in global symptom severity from baseline to 6 months (ß=-0·04, p=0·033), mediated by increases in self-reported exercise. The direct effect of the intervention remained statistically significant (ß=-0·14, p=0·006), suggesting partial mediation (appendix pp 74–75).

Over the 12-month follow-up period, a total of 220 participants reported AEs, including 85 (62·5%) of 136 patients in the ImPuls plus TAU group and 135 (70·7%) of 191 participants in the TAU-only group. Furthermore, 14 (0·07%) of 198 patients in the ImPuls plus TAU group reported 15 SAEs and 22 (10·48%) of 201 patients in the TAU-only group reported 24 SAEs (Table 3). There were no significant differences in the total number of AEs or SAEs between the two groups. Eight study drop-outs were associated with reported AEs, and two were associated with SAEs. There was one SAE (torn ligament) related to the intervention (appendix pp 76–77).

**Table 3.** Serious adverse events during the entire ImPuls trial for both groups; TAU = treatment as usual; Severity was rated by the external Data Safety and Monitoring Board (DSMB)

|  | ImPuls plus TAU<br>(*n* = 199) |  | TAU<br>(*n* = 201) |  |
| --- | --- | --- | --- | --- |
|  | Events | Participants | Events | Participants |
| **Related to intervention** | 1 | 1 | N/A | N/A |
| **Related to psychiatric events** | 10 | 9 | 15 | 14 |
| **Related to physical events** | 5 | 5 | 9 | 9 |
| **Total** | 15 | 14 | 24 | 22 |
| **Severity** |  |  |  |  |
| low | 0 | 0 | 0 | 0 |
| moderate | 3 | 3 | 4 | 4 |
| high | 12 | 11 | 20 | 19 |

## Discussion

This large pragmatic RCT demonstrated the efficacy of ImPuls, a transdiagnostic group exercise intervention, plus treatment as usual (TAU) in reducing global symptom severity and symptoms of depression, general anxiety, panic, and PTSD at 6 months after baseline compared to the control group receiving TAU alone. Treatment effects were maintained up to 12 months after baseline for global symptom severity, depression, general anxiety, and panic. Importantly, the intervention group also showed better clinical recovery and improvement at both the 6-month and 12-month assessments compared to control. While the intervention resulted in a substantial increase in self-reported exercise, this was not reflected in the accelerometer-based MVPA results at 6 months.

A sensitivity analysis focusing exclusively on patients who adhered to the minimum intervention dose confirmed the main analysis and yielded slightly larger effect sizes. Furthermore, the increase in self-reported exercise from baseline to 6 months partially mediated the treatment effects on global symptom severity in the analysis of completers, thus supporting the rationale behind the exercise intervention. The treatment drop-out rate of 19·10% was comparable to rates reported in other RCTs involving exercise interventions for outpatients with depression in mental healthcare services (19%) and anxiety and stress-related disorders (17·9%),^10,26^ as well as to rates observed for psychological treatments in real-world contexts (26%).^27^ The probability of study drop-out in the intervention group was higher among patients diagnosed with agoraphobia, which may be due to group outdoor exercise being an anxiety-provoking situation for this patient population.

In existing meta-analyses, the focus has typically been on disorder-specific interventions in disorder-specific samples. Moreover, to date, only a limited number of meta-analyses have investigated the efficacy of exercise as an adjunct to TAU in patients with depression.^11,12,14^ Our study replicates the positive findings of these analyses but shows considerably smaller effect sizes. This discrepancy may partly result from the comparison in our study between exercise as an adjunct to TAU and TAU alone, which constitutes a more stringent test than comparisons involving waitlist or passive control groups. Such control groups have often been used as comparator groups in the earlier studies underlying the meta-analyses published to date, often without clear differentiation between passive and active controls. Moreover, the use of transdiagnostic measures in our study, which tend to be less specific than disorder-specific measures, especially in samples of patients with various mental disorders, may also have contributed to our smaller effect sizes. Lastly, a recent meta-analysis of studies comparing transdiagnostic psychological treatments against passive control groups or TAU in patients with major depression and anxiety disorders reported small to moderate effects similar to those found in our study.^28^

In our study, the intervention group saw an increase in self-reported exercise to over 90 minutes at 6 months, which aligned with our intervention manual and recent evidence on the disorder-specific efficacy of exercise in mental disorders. ^8,9^ At 12 months, the average weekly exercise duration remained high at 69 minutes, indicating that patients in the intervention group, on average, continued to maintain the necessary exercise volume. Contrary to our expectations, the TAU group also experienced a substantial increase in exercise volume from baseline to 12 months, possibly reflecting heightened motivation for exercise stimulated either by participation in the study or through the effects of TAU itself.

The existing literature lacks trials that simultaneously report on exercise volume, its long-term sustainability, treatment efficacy, and the role of exercise volume as a mediator of treatment effects.^8^ Among the few trials that have considered exercise volume as an outcome, changes in exercise volume, regardless of group allocation, have been correlated with symptom reduction.^20,29^ In this context, our study adds unique insights by identifying a partial mediation effect of treatment outcomes through increases in self-reported exercise. However, this partial mediation was significant only in the completer analysis and from baseline to 6 months, suggesting that other factors contributed to the clinical efficacy observed. Evidence from our earlier feasibility study had already suggested that the long-term effects of ImPuls are not solely due to increased exercise volume but also to the deliberate use of exercise for affect regulation, which may also be true in the current trial.^19^ Interestingly, accelerometry-based physical activity data did not show differential increases and were not associated with changes in global symptom severity. This could be due to the physical presence of the accelerometer motivating all patients to engage in exercise, as reflected in the consistently high activity levels across all measurement time points. Additionally, the data captured by accelerometers lack specificity because they include routine daily activities, such as domestic and occupational tasks, which have inconsistent associations with mental health.^30^

Our study has several important limitations. First, we encountered differential drop-out rates between the groups. Comparing these rates is challenging due to the compensation provided exclusively to the control group, which may have contributed to its lower attrition rate. Here, it is also important to note that patients diagnosed with agoraphobia and those who had received previous psychiatric treatment were more likely to drop out of the intervention group.

To mitigate the influence of attrition bias, we incorporated both variables as predictors in our missing data imputation procedures. Second, similar to existing evidence-based standard treatments, our study identified a substantial proportion of non-responders, indicating that a subgroup of patients did not benefit from the exercise intervention. Lastly, our research design does not allow us to determine whether non-specific factors, such as daily routine or social support, contributed to the observed effects.

Our study also has several notable strengths. First, it achieved a high and predefined sample size, ensuring sufficient statistical power for the analyses. Second, the study exhibits strong external validity, accurately replicating real-world outpatient care conditions. This was achieved by including a diverse range of highly prevalent mental disorders and patients with substantial comorbidity, as well as by involving exercise therapists in genuine health care settings and routine care providers as the referring health professionals. Third, the inclusion of a long-term follow-up assessment six months post-intervention adds to the robustness of our findings and provides evidence of the sustained impact of the treatment. Lastly, the methodological rigour of the study is evidenced by its structured clinical eligibility assessments, intention-to-treat analysis, involvement of a blinded external statistician, and process evaluation. Together, these aspects underscore the study’s high level of internal validity. The results of the process evaluation, including high attendance rates, acceptable drop-out rates, strong treatment fidelity, and substantial intervention acceptability, support the credibility of the treatment effects identified in the present trial.^24^

Overall, the findings of our study suggest that group interventions like ImPuls, which combine exercise with behaviour change techniques, offer a viable and promising adjunctive treatment for major depression, insomnia, agoraphobia, panic disorder, and PTSD. Future research should analyse the reasons for non-response in order to refine ImPuls and similar interventions, and to improve response rates. Additionally, comparative studies are warranted to compare ImPuls to standard transdiagnostic psychological interventions. Such research could determine whether ImPuls might also serve as an alternative to standard treatment in routine care settings.

## Ethics approval and consent to participate

The study was conducted according to the guidelines of the Declaration of Helsinki of 2010 and was approved by local ethics committee for medical research at the University of Tübingen (ID: 888/2020B01, 02/11/2020).

## Availability of data and materials

Individual participant data that underlie the results reported in this article will be published after de-identification. Documents that will be shared further: Study protocol, analytic code, aggregated individual study data.

Routine/administrative data from the participating statutory health insurers will not be made available. Access to data will be provided to anyone legitimately interested in it. Analytic code and aggregated individual study data are available on OSF (https://osf.io/tpsb4). Participants gave informed consent for their data to be published after de-identification (except for the routine/administrative data from the statutory health insurers).

## Authors’ contributions

S.W., J-M.Z., B.S., N.E-K./A.L.F., L.Z., L.S., A.R-M., M.H., G.S., and T.E. contributed to the conception and the design of the study and acquisition of funding. S.W. was responsible for administration of entire project; T.E., A.R., L.Z., N.E.-K./A.L.F., S.P., G.S., T., E., and L.S. were responsible for project administration as consortium partners, S.W., J.-M.Z., B.S, J.W., L.B., T.S., A.K.F., E.M., S.R., and D.V.F. were responsible for study organisation, recruitment and assessment, training of the exercise therapists and data management. S.R., D.V.F., S.W., and G.S. were responsible for the process evaluation, development of treatment fidelity score, data management and training of the exercise therapists. F.H., A.R., and A.R-M. were responsible for the app design, development and maintenance, S.P. was responsible for the recruitment of the study sites and the qualification of the exercise therapists, A.L.F. and L.Z. were the representatives of the two statutory health insurer, providing the routine data for the health economics analysis and supporting patient recruitment, E.H., M.M.G, K.T., T.N., and T.E. were responsible for data management, data handling, the randomisation procedure, data pre-processing and analysis of treatment fidelity, K.T. was the blinded external statistician responsible for the formal statistical analysis, S.K., S.F., and L.S. were responsible for the health economic analysis. Original draft preparation was done by S.W. All authors contributed to the drafting and revision of the final study protocol.

## The use of AI and AI-assisted technologies in scientific writing

During the preparation of this work, the authors used OpenAI’s Chat GPT (version 3.5) to improve the manuscript’s wording, readability, and language quality. This tool was used only for language refinement and not for generating text. After using this tool, the authors reviewed and revised any material processed through it and take full responsibility for the content of the publication.

## Data Availability

Individual participant data that underlie the results reported in this article will be published after deidentification: text, tables, figures and appendices. Documents that will be further shared: Study protocol, statistical analysis plan, analytic code, aggregated individual study data. Routine/administrative data from health insurances will not be made available. Anyone who wishes to have access the data will have access to the data. Analytic code and aggregated individual study data will be made available on an online repository immediately after publication (or within the peer review process). Participants give informed consent to publish their data after deidentification (despite the routine/administrative data from the health insurances).

## Acknowledgements

The authors gratefully acknowledge all cooperating partners and exercise therapists who carried out the intervention, cooperated in all matters of research (e.g. documentation of sessions) and supported the recruitment process: RehaZentrum Hess (Bietigheim and Crailsheim), Therapiezentrum Heidelberg (Theraktiv GbR), VAMED Rehazentrum Karlsruhe GmbH, Karlsruhe; Universitätsklinikum Zentrum für Physiotherapie, Tübingen; VAMED Rehazentrum Ulm GmbH, Ulm; RehaZentrum Weingarten; ZAR Göppingen; Rehamed Stuttgart; Rehaklinik / ZAPR Glotterbad, Freiburg. We thank all general practitioners, psychiatrists, psychotherapists, clinics, hospitals, social media influencers, and newspapers that supported the recruitment process. We would especially like to thank all student assistants who supported us in the recruitment and training process and Prof. Dr. Felipe Schuch for critically reviewing the paper. We extend special thanks to our scientific language editor, Matthew Gaskins, for his invaluable contributions to refining the language and coherence of this manuscript.

## List of abbreviations

AE: Adverse events
AOK BW: Allgemeine Ortskrankenkasse Baden–Württemberg (German statutory health insurer)
App: (smartphone) Application
BCT: Behavioral change techniques
BSA: Physical Activity, Exercise, and Sport Questionnaire
BSI-18: Brief Symptom Inventory 18
CBT: Cognitive behavioural therapy
CONSORT: Consolidated Standards of Reporting Trials
DALY: Disability-adjusted life years
DSM-5: Diagnostic and Statistical Manual of Mental Disorders (5^th^ version)
DSMB: Data safety and monitoring board
DVGS: German Association for Health-enhancing Physical Activity and Exercise Therapy
GAD-7: Generalized Anxiety Disorder scale
ICD-10: International Classification of Diseases (version 10)
ISI: Insomnia Severity Index
ITT: Intention-to-treat
MET: Metabolic equivalent of task
MVPA: Moderate-to-vigorous physical activity
PARQ: Physical Activity Readiness Questionnaire
PCL-5: The Posttraumatic Stress Disorder Checklist 5
PHQ-9: Patient Health Questionnaire-9
PSQI: Pittsburgh Sleep Quality Index
PTSD: Post-traumatic stress disorder
RCI: Reliable change index
RCT: Randomised controlled trial
REDCap: Research Electronic Data Capture (software)
RPE: Rating of Perceived Exertion
RPE Scale: Rating of Perceived Exertion Scale
SAE: Serious adverse events
SKID-5-CV: Structured clinical interview for DSM 5 – clinical version
TAU: Treatment as usual
TK: Techniker Krankenkasse (German statutory health insurer)

### Funding

This trial is fully funded by the German Innovation Fund of the Federal Joint Committee of Germany from September 2020 to June 2024 (01NVF19022). This funding source had no role in the design of this study and will not have any role during its execution, analyses, interpretation of the data, or decision to submit results

### Trial Sponsor

University of Tuebingen Dr. Sebastian Wolf Hölderlinstraße 19 72074 Tuebingen Deutschland Tel.: +49 (0)7071 29-73613 Mail

**Organizational Structure and responsibilities:**
| Responsible | Institution | Responsibility |
| --- | --- | --- |
| Dr. Sebastian Wolf | Institute of Sports Science,<br>Education and Health Research,<br>Junior Research Group „Exercise<br>and Mental Health“, Eberhard Karls<br>University Tuebingen <sup>1</sup> | Project PI: Coordination,<br>Administration, Recruiting, Diagnostics,<br>training of therapists |
| Prof. Dr. Martin<br>Hautzinger | Department Psychology, Clinical<br>Psychology and Psychotherapy,<br>Eberhard Karls University<br>Tuebingen <sup>2</sup> | Consulting, principal investigator |
| Prof. Dr. Gorden<br>Sudeck | Institute of Sports Science,<br>Education and Health Research,<br>Eberhard Karls University<br>Tuebingen <sup>1</sup> | supervision for exercise therapists,<br>training of exercise therapists, process<br>evaluation |
| Dr. Ander Ramos Murguialday | Institute of Medical Psychology and Behavioral Neurobiology<br>Eberhard Karls University Tuebingen <sup>3</sup> | App programming, testing and maintenance |
| Prof. Dr. Thomas Ehring, | Department Psychology, Clinical Psychology and Psychotherapy, Ludwig Maximilians University Munich (LMU) <sup>4</sup> | Randomization, Data Management, Data storage, Analysis of clinical data |
| Prof. Dr. Leonie Sundmacher | Health Economics, Technical University Munich (TUM) <sup>5</sup> | Health economics, Trust center |
| Angelika Baldus<br>Stefan Peters | German Association of health-related Fitness and Exercise Therapy (German: DVGS) <sup>6</sup> | Recruitment of study sites, Qualification of exercise therapists |
| Anna Lena Flagmeier | AOK Baden-Württemberg <sup>7</sup> | Implementation of treatment structure in health insurance system, data management of routine data, patient recruitment |
| Lena Zwanzleitner | Techniker Krankenkasse <sup>8</sup> | Implementation of treatment structure in health insurance system, data management of routine data, patient recruitment |

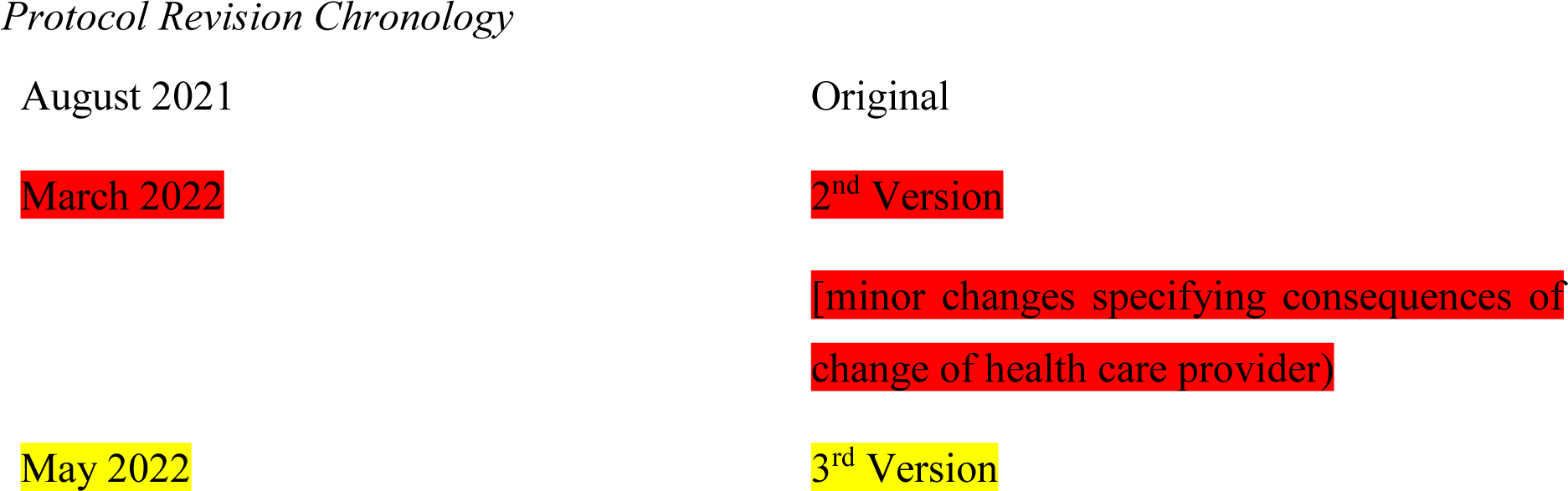
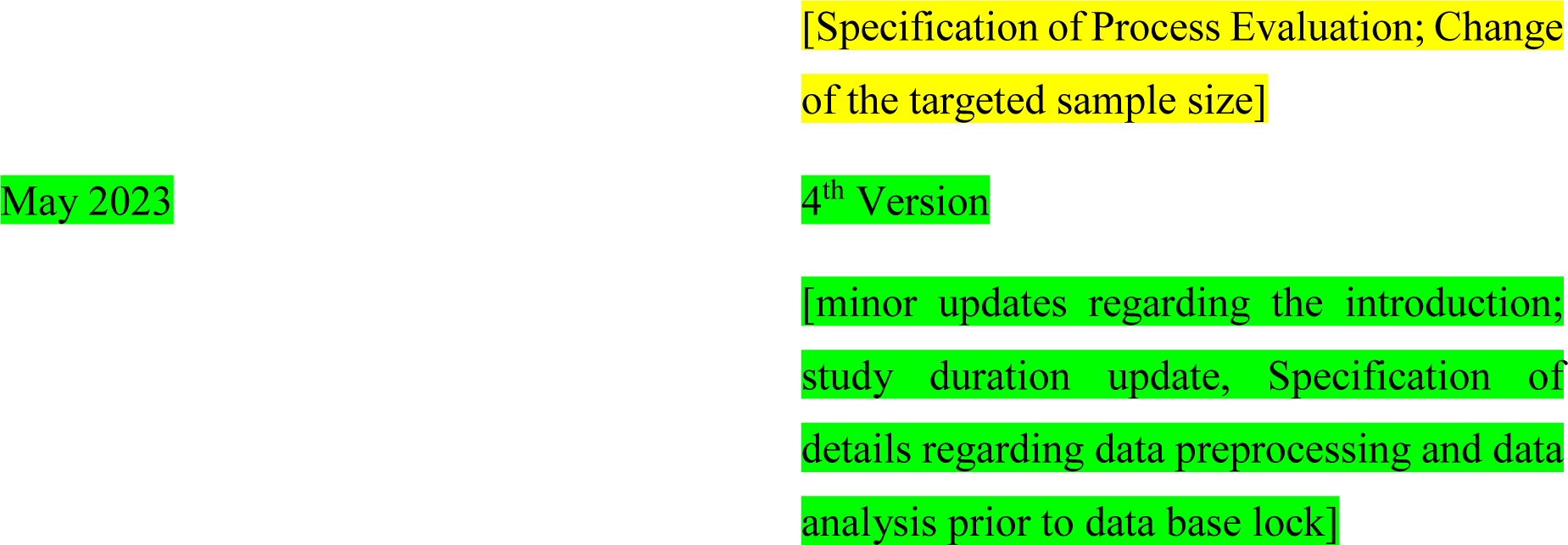

### Introduction

#### Description of research question and justification for undertaking the trial

Epidemiological data from 2019 suggests that 15.6 % of the German population suffered from any mental disorder in 2019 (point prevalence).^1^ Mental disorders result in a considerable burden of disease, for example accounting for 6.4 % of overall disability-adjusted life years (DALYS) assessed in the 2019 epidemiological survey.^2^ From 2008 to 2018, the proportion of mental disorders among all causes of death increased from 2.2 % to 6.1 %.^3^ The most prevalent disorders in Germany are anxiety disorders and trauma- and stress-related disorders (point prevalence: 7.1 %), major depressive disorders (point prevalence: 4.3 %), as well as insomnia (point prevalence: 4 %).^2,4^ Notably, these disorders often occur comorbidly and share common underlying aetiological and even maintenance mechanisms, such as the experience of stressful life events and high perceived stress, low self-efficacy, sleep disturbances, elevated anxiety sensitivity, or repetitive negative thinking.^4–14^

Worldwide, 27.2 % of the DALYS attributable to mental disorders can be explained by major depressive disorders and 16.3 % by anxiety disorders.^15^ In 2015, health care costs in Germany caused by mental disorders amounted to 44.4 billion euros.^16^ Of this, 8.7 billion euros can be attributed to major depressive disorders, 1.7 billion euros to phobic and other anxiety disorders, and 1 billion euros to insomnia. Mental disorders account for 13.1 % of total costs and represent the second highest cost group after cardiovascular disorders (46.4 billion euros, 13.7 % of total costs). Major depressive (RR = 2.63) and anxiety disorders (RR = 1.41) have also been shown to increase the risk of cardiovascular disease.^17^ Besides direct costs (e.g., treatment costs), mental disorders cause indirect costs on the German job market. With 14.4 billion euros overall, mental disorders caused the second-highest lost production costs of all diagnosis groups in 2019.^18^ They further caused 117.2 million days (16.5% of all days) of incapacity to work, which is the longest absences per sick leave of all disorders.^19^ Amongst mental disorders, major depressive disorders accounted for the most days of incapacity to work (33.9 million days), followed by trauma- and stress-related disorders (21.6 million days). Anxiety disorders accounted for 7.6 million days and insomnia for 0.5 million days.

Despite the high prevalence and severe negative impact of mental disorders, it is estimated that in Germany only 10 % of all affected individuals receive evidence-based treatment and only 2.5% receive psychological treatment.^20^ In addition, even those receiving psychological treatment often have to wait before treatment initiation; for example, 40 % of outpatients waited three to nine months to start psychotherapeutic treatment in German health care settings.^21^ Longer waiting times are associated with worsening and chronicity of symptoms and the development of comorbid conditions.^22^ The high prevalence and severe burden of mental disorders in combination with the large gap between people in need for treatment and those actually receiving it illustrates the need to develop alternative efficacious, effective and efficient treatments.^23^

Exercise, defined as physical activity that is planned, structured and repeated, with the primary aim to improve or maintain physical fitness, has revealed positive therapeutic effects for diverse mental disorders.^24,25^ Most of the studied exercise interventions include aerobic activities (i.e. running) or a combination of aerobic exercise with strengthening activities. Recent meta-analyses on major depressive disorders and insomnia have shown large effects for exercise that were comparable to those of psychological treatment and psychopharmacological treatment.^26,27^ A recent meta-analysis on PTSD found small to moderate effects; however, in two of the four studies included, the intervention comprised yoga.^28^ Looking at more recent evidence from RCTs focusing on interventions including aerobic exercise, large treatment effects were found.^29,30^ For panic disorder (with and without agoraphobia), RCTs have revealed large effects on symptomatology with both, acute exercise and structured multi-week aerobic exercise programs.^31,32^ In addition, moderate to large effects have been reported for exercise as an augmentation to TAU for major depressive disorders, panic disorder and PTSD.^30,33,34^ Exercise also seems to be efficacious in a transdiagnostic way as it targets the aforementioned underlying aetiological and maintaining factors that are present across depressive disorders, anxiety disorders and insomnia such as low self-efficacy, sleep disturbances or higher stress perception and adaptive stress coping.^35,36^ Key components of exercise interventions that have shown optimal therapeutic efficacy among patients with major depressive disorders, insomnia, panic disorder with or without agoraphobia and PTSD include aerobic exercise at a minimum of moderate intensity (MVAE) either or a combination of MVAE with resistance training, conducted two to three times per week, for 10 weeks with a session duration of at least 30 minutes, partially supervised or non-supervised.^27,35,37,38^

Exercise might not only be a promising efficacious (transdiagnostic) treatment but also carry the advantage of being highly efficient, since it can be delivered in group settings with relatively short durations, can be offered to patients with heterogeneous and burdensome mental disorders, can be expected to show a low likelihood of adverse effects, comes at a relatively low cost, and is suited to reduce the risk for cardiovascular diseases that frequently occur comorbid with mental disorders.^27,39,40^ Furthermore, exercise can be performed and continued independently without professional supervision or only remote supervision. However, individuals suffering from mental disorders often have difficulties to initiate and maintain a physically active lifestyle, which may be related in part to deficiencies in motivation and exercise-related self-regulatory skills in this population.^41,42^ Reassuringly, there is evidence showing that exercise adoption and maintenance are mediated by motivational and volitional aspects, such as intention strength, action planning and barrier management.^43^ A recent meta-analysis shows that especially self-efficacy in building intentions and action planning is crucial for sustained exercise behavior change.^44^ One possible way to promote such motivational and volitional aspects are the application of behavior change techniques (BCTs).^45,46^

The combination of behavior change techniques and exercise as a structured intervention appears highly promising in terms of initiating a sustainable exercise behavior change. Since outpatients have less supervision and contact to their therapists compared to patients in inpatient or rehabilitative mental health care settings, structured exercise interventions in combination with behavior change techniques to overcome general and disorder-specific barriers might be especially important within the outpatient mental health care setting. Indeed solely exercise on prescription (or on referral) for outpatients shows drop-out rates of nearly 80%, whereas structured exercise interventions in combination with BCTs for outpatients show lower dropout rates and stronger effects on mental health.27,47

Therefore, ImPuls was developed and evaluated as a complex exercise program with respect to the Medical Research Council (MRC) framework, specifically designed to scrutinize an additional treatment option in the outpatient mental health care system in Germany.48 It has been successfully evaluated in terms of efficacy and acceptability in a feasibility study for outpatients waiting for psychotherapeutic treatment.49,50 A broader and more comprehensive pragmatic trial was needed to explore the extent to which the intervention also achieves its effect in a real-world setting (e.g., with exercise therapists working in the outpatient setting as intervention deliverers alongside their daily business; as an add-on to treatment as usual; with a realistic referral system).51 Therefore, it is now conducted in a pragmatic multi-site randomized controlled trial to investigate efficacy and cost-effectiveness within the real-world outpatient setting.52

The complexity of ImPuls (i.e., several interacting factors [e.g., BCTs and Exercise], involvement of different actors [exercise therapists, managers of outpatient rehabilitative and medical care facilities, patients] etc.) and the future need to implement the intervention into a comprehensive health service provision prompts the necessity of research beyond a pure evaluation of efficacy, namely process evaluation. Thus, the ImPuls study is accompanied by a comprehensive process evaluation based on the MRC framework and its complement, the former of which provides comprehensive and detailed guidance.53-55 Using a mixed-methods approach may be particularly helpful to understand multiple perspectives, multiple types of causal processes and multiple types of outcomes which in turn are common aspects of implementation research.56 Process evaluation in studies evaluating exercise interventions has been slowly emerging during the last decade.57-59 However, process evaluations of exercise interventions offered to patients with mental disorders is rarely conducted. For example, one study focused solely on selected aspects like adherence rate of the participants, acceptability and feasibility, which means that only specific subcomponents of the Medical Research Council (MRC) Framework were taken into account.55,60 Other studies exclusively conducted qualitative interviews, which ideally should be complemented by quantitative methods to provide an encompassing insight into the processes relevant for implementation.56,61,62 Another study heeds the aforementioned deficiencies, yet apparently seems to omit investigation of interactions (e.g., between participants and the intervention / -deliverers) with regard to the MRC framework key component mechanisms of impact.63 Given the lack of comprehensive process evaluations accompanying exercise programs for patients with heterogeneous mental disorders, the respective evidence for implementation conditions is weak. Consequentially, further research in this area is needed.

#### Explanation for choice of comparators

The TAU condition will be modelled to represent the typical treatment patients receive in the German outpatient health care system. Therefore, patients will not be actively provided with any treatment but patients are allowed to receive any intervention that is available to them. Any evidence-based treatment provided by the outpatient mental health care system will be recorded, i.e., any psychiatric/pharmacological or psychological/psychotherapeutic intervention. Interventions, delivery or dosage of the intervention can be changed and adapted during the course of the study.

#### Specific objectives and hypotheses

Despite the promising evidence for MVAE as an intervention for patients with mental disorders, exercise programs or professional exercise therapy are currently not provided as regular health services within the outpatient mental health care system in Germany. With the aim of combining the current evidence on the efficacy of MVAE and sustained exercise behavior change with specific demands of a real-world outpatient health care setting, ImPuls was developed as a manualized group exercise intervention for physically inactive outpatients suffering from major depressive disorders, insomnia, panic disorder with or without agoraphobia and PTSD.^48^ A feasibility study found moderate long-term effects in patients waiting for psychotherapeutic treatment who completed ImPuls compared to a passive control group.^49,50^ ImPuls integrates recent findings about the optimal modalities of exercise for therapeutic efficacy, such as optimal frequency, intensity, time/duration and type of exercise (FITT criteria) for the targeted disorders and evidence regarding sustainable behavior change by integrating behavior change techniques (BCTs). The components of this intervention are further tailored towards the specific needs of outpatients with mental disorders in the current German mental health care setting. Specific features are 1) the inclusion of a broad range of heterogenous diagnoses for which prior research has demonstrated therapeutic transdiagnostic efficacy, 2) intervention delivery in group format, and 3) short duration (i.e., only 4 weeks of supervised MVAE sessions carried out inhouse in each study site). The aim of the current pragmatic trial is to investigate the efficacy and cost-effectiveness of implementing ImPuls within the outpatient mental health care setting in Baden-Württemberg, a representative state in South-West Germany. The following hypotheses will be tested:

1. Participants in the intervention condition, who have received ImPuls in addition to TAU, will show lower global symptom severity at post-treatment and follow-up assessments compared to a control condition with TAU only.
2. Overall costs in the intervention condition will represent a significant saving for the public health system compared to the control condition at post-treatment and follow-up assessments.
3a. The intervention will lead to significantly higher levels of MVAE at post-treatment and follow-up assessments compared to the control condition.
3b. The effect of condition on the reduction of the primary outcome global symptom severity will be mediated by an increase in MVAE.
4. Participants in the intervention condition will show lower disorder-specific symptoms (major depressive disorder, insomnia, panic disorder with or without agoraphobia and PTSD) compared to participants in the control condition at post-treatment and follow-up assessments

We will further assess, if participants in the intervention condition show more instances of clinically significant change compared to participants in the control condition at post-treatment and follow-up assessments (Additional analysis to Hypothesis 1 and 4).

The main objectives of our process evaluation are a) to support the findings of the ongoing pragmatic randomized controlled trial by confirming that its efficacy is truly attributable to the ImPuls intervention and b) to discover further crucial factors for the implementation of ImPuls into real-world outpatient mental health care settings.

The main research questions of the process evaluation are:

1. Implementation:
  a. To what extent do our actions empower exercise therapists (i.e., competence, acceptance) to deliver the intervention?
  b. To what extent do exercise therapists implement intervention components as intended (treatment fidelity) and what are reasons for its potential variance?
  c. Which strategies recruit the most patients and how valid are the referrals in terms of acquisition/inclusion?
  d. How do referring healthcare professionals rate the ImPuls intervention in terms of acceptability, appropriateness and feasibility?
  e. To what extent are all ImPuls sessions offered as planned and all telephone contacts made as scheduled?
2. Context:
  a. What barriers and facilitators do exercise therapists and managers experience concerning the implementation of the ImPuls intervention?
3. Mechanisms of Impact:
  a. To what extent do attitudes of exercise therapists towards the ImPuls intervention (e.g., acceptability, appropriateness) moderate the treatment effects?
  b. To what extent do patients’ integration of core components of the ImPuls intervention (e.g., amount of exercise, barrier management, goal-setting) as well as changes in respective individual behavioral determinants (e.g., action and coping plans; physical activity-related health competencies) mediate the treatment effects?
  c. To what extent do patients’ integration of motivational/volitional core components of the ImPuls intervention (e.g., barrier management, goal-setting, phone contacts) as well as changes in respective individual behavioral determinants (e.g., action and coping plans; physical activity-related health competencies) mediate its effect on their exercise adherence?
  d. To what extent do changes in patients’ transdiagnostic psychological processes (e.g., emotional regulation, repetitive negative thinking or perceived stress) mediate the treatment effects?.

### Methods

#### Description of trial design

**Figure P1:**
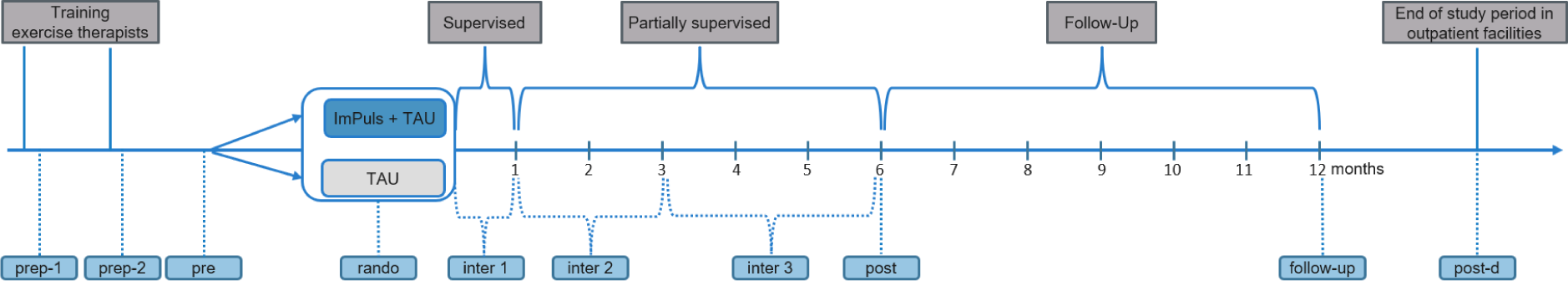
Research design and measurement points of the entire trial including process evaluation. *Note:* follow-up = 12 months after randomization; Inter-1 = once at the end of the supervised ImPuls phase (week 1-4); inter-2 = once at the end of the partially supervised ImPuls phase 1 (week 5-12); inter-3 = once at the end of the partially supervised ImPuls phase 2 (week 13-24); post = after completion of the intervention (12 months after randomization, week 24-26); post-d = end of study period in the outpatient facility; pre = prior to randomization; prep-1 = following the first training; prep-2 = following the second training; rando = after randomization, prior to intervention start in the outpatient facility.

The study will be led by researchers based at the University of Tuebingen in Germany, and will be conducted in ten different study sites across Baden-Württemberg, a region in South-West Germany. Study sites are local outpatient rehabilitative and medical care facilities which were selected to cover the different regions in Baden-Württemberg. The entire project will be conducted between September 2020 and June 2024. The study has been registered at the German Clinical Trial Register (ID: DRKS00024152, 05/02/2021) and has been approved by the local ethics committee for medical research at the University of Tuebingen (ID: 888/2020B01, 02/11/2020). A pragmatic multi-site block-randomized controlled trial with two treatment arms (ImPuls + TAU vs. TAU) and three points of assessment (pre, post, follow-up) will be conducted (see fig. 1). All outcomes will be included at all assessments. Study completion and reporting will be carried out in accordance with the Consolidated Standards of Reporting Trials (CONSORT), the Template for Intervention Description and Replication (TIDieR) and the Consensus on Exercise Reporting Template (CERT).^64,65^

#### Description of study settings

Data will be collected in Baden-Württemberg, a region south-west of Germany. Study sites are mostly big centers of psychosomatic, orthopedic or cardiological rehabilitation and one outpatient unit of physiotherapy. A list of all locations/study sites can be obtained in the registration: https://drks.de/search/en/trial/DRKS00024152

#### Eligibility criteria

##### Participants

Inclusion criteria are age between 18 and 65 years, membership of the AOK BW or TK, fluent in German, no medical contraindications for exercise, and diagnosed according to ICD-10 with at least one of the following disorders: depressive disorders (F32.1, F32.2, F33.1, F33.2), agoraphobia (F40.0, F40.01), panic disorder (F41.0), PTSD (F43.1) or insomnia (F51.0). Exclusion criteria included: Exercising of at least twice a week for at least 30 minutes each, continuously over a period of 6 weeks within the last 3 months before study diagnosis, sports-medical contraindication (medical consultation), acute mental and behavioral disorders due to psychotropic substances (F10.0, F10.2-F10.9; F11.0, F11.2-F11.9; F12. 0, F12.2-F12.9; F13.0, F13.2-F13.9; F14.0, F14.2-F14.9; F15.0, F15.2-F15.9; F16.0, F16.2-F16.9; F17.2-F17.9; F18.0, F18.2-F18.9; F19.0, F19.2-F19.9), acute eating disorders (F50); acute bipolar disorder (F31), acute schizophrenia (ICD-10 F20), acute suicidality.

### Exercise therapists/study therapists

To carry out the intervention, exercise therapists are required to have one of the following academic or comparable basic qualifications as physical activity and exercise professionals with a training period of at least 3 years: academic degree in exercise or movement science with at least 10 ECTS practice and 20 ECTS theory (e.g. Magister, Bachelor, Master, Diploma Physical Education, Exercise Science, Exercise Physiology), non-academic technical college degree Exercise and Caring/Therapeutic Gymnastics with at least 21 semester hours per week, non-academic technical college degree „Physical Educator as liberal profession“, academic and non-academic degrees in physiotherapy. Moreover, a specific additional therapeutic qualification DVGS e.V. with 5 ECTS overall is required with the following content: Physical Activity-related Health Competence (2 ECTS), Basics in Health Science and Health Pedagogy (1 ECTS), Basics in Psychiatry, Psychosomatics and Addiction (1 ECTS), Affective Disorders (1 ECTS).

All therapists will be required to attend a dedicated training for the ImPuls intervention in the future. During these training sessions, therapists will be thoroughly prepared for the implementation of the study, with a particular focus on the intervention phase. A total of three training sessions will take place in a physical, face-to-face setting. These sessions will consist of a two-day and a one-day training, both of which will be held at the Institute for Sports Science of the University of Tübingen. Following the initiation of the first group within a given center, an in-house training session will be conducted.

The content of these future training sessions will encompass various essential aspects, including the therapeutic management of specific mental illnesses included in the study, addressing clinical crises, the application and handling of the smartphone app, and, notably, the testing of specific sessions outlined in the ImPuls manual. Role-playing exercises involving actors will be utilized to practice core sessions and central elements of the intervention, which will subsequently be subjected to intensive discussions.

Key components of the role-playing exercises will include psychological goal setting, individual barriers management, imagination techniques, crisis management, and the execution of outdoor running sessions, including considerations of duration and interval methodology, in addition to the proper utilization and adjustment of heart rate monitors. It is important to note that all future training sessions will undergo continuous evaluation and adjustment in response to the preferences and feedback of the participants, thus ensuring a dynamic and adaptive training process.

#### ImPuls Intervention

The exercise intervention “ImPuls” ^48^ will be delivered to groups consisting of 6 patients and will be divided into a supervised and partially-supervised period. BCTs, such as goal setting, self-monitoring, formation of concrete exercise plans and coping planning, will be integrated to promote sustained exercise behaviour change.^45,46,66^ The intervention structure and contents are displayed in Figure P2 and Table P1. Participants will receive ImPuls in addition to TAU.

### Supervised Period (weeks 0-4)

Patients will participate in a combination of supervised MVAE sessions and group sessions with educative elements integrating BCTs (see table P1) in groups with a total duration of 120 minutes each session. Supervised MVAE will be provided twice a week and will consist of either running or fast walking. MVAE will last 30 minutes and participants can choose between a standardized interval-based or endurance method protocol. Both training methods will be conducted with at least moderate intensity, which is tracked by a heart rate monitor (SIGMA iD.FREE) combined with a chest strap (SIGMA R1 Bluetooth Duo Comfortex+) and the Borg Rating of Perceived Exertion (RPE) Scale.^67^ Moderate to vigorous intensity is defined as at least 64 % of maximum heart rate, subtracting age from 220 and at least 13 points of the RPE Scale.^67,68^ The ImPuls smartphone application (“ImPuls-App”) developed specifically for ImPuls supports the participants and therapists during MVAE. In weeks 2, 3 and 4 patients will engage in additional 30-minutes MVAE, which is chosen based on their own interests and preferences. Therapists provide a list of MVAE highlights in each study site (i.e. offers in local sport clubs, yoga studio, gyms) which can be found and selected in the “ImPuls-App”.

### Partially Supervised Period (weeks 5-24)

Participants will be asked to engage in 30-minutes non-supervised MVAE at least twice a week. Regular MVAE will be planned through specific training schedules and accompanied by activity diaries, self-monitoring of goals and volitional strategies and weekly (weeks 5-12)/biweekly (weeks 13-24) phone calls with the exercise therapist, intending to maintain motivation, volition, and adherence to exercise. Training plans and all documentation will be executed and coordinated via the “ImPuls-App”. Information will be shared with the therapists in advance prior to the phone calls. A session for patient’s supporters (e.g., friends, partner) will be scheduled in Week 5 to inform them about the possibilities to support the participants in transforming their intentions into action.

**Figure P2:**
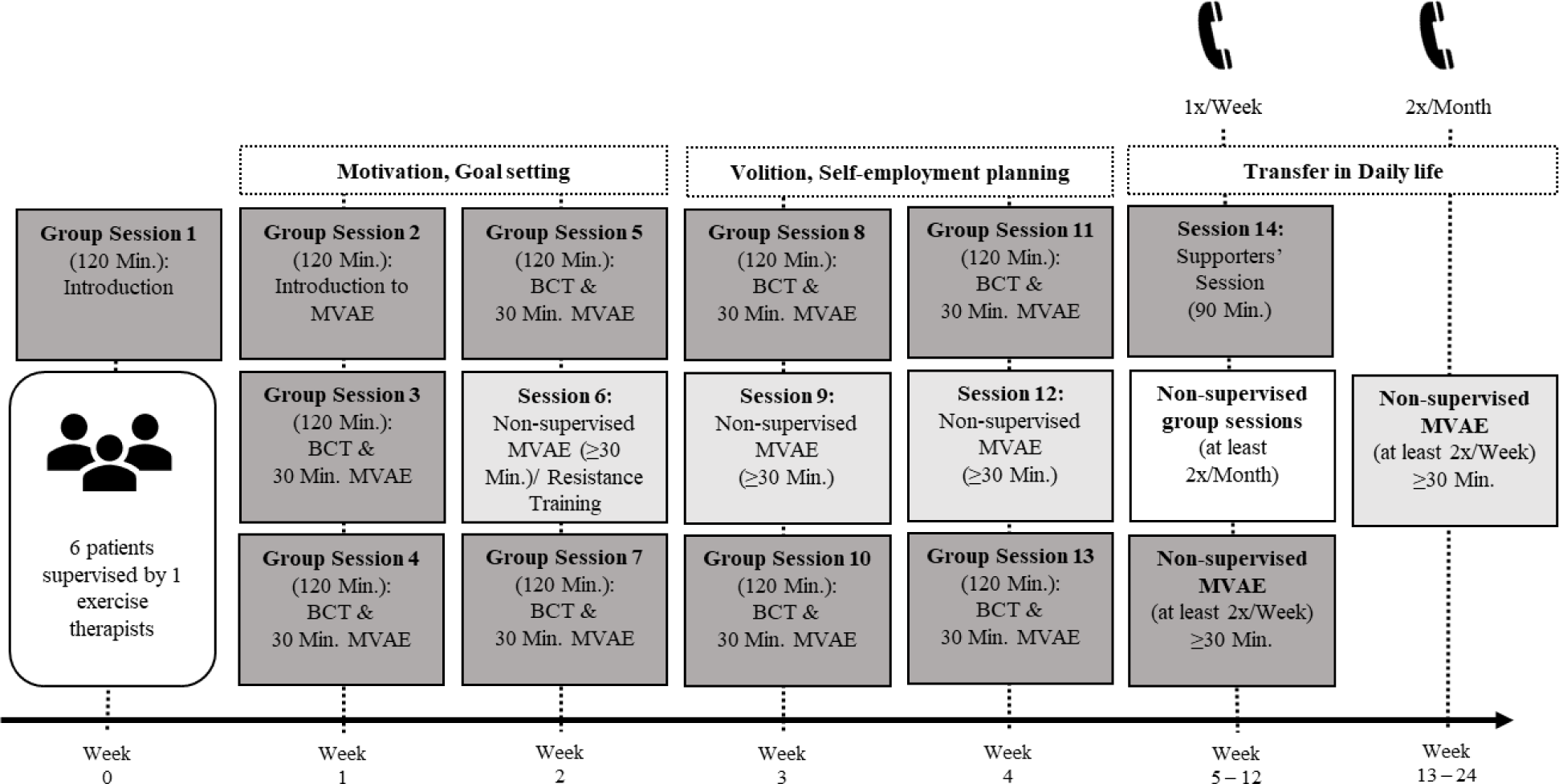
The temporal program structure and content overview of ImPuls.

The temporal program structure and content overview of ImPuls. The dark gray boxes illustrate the supervised sessions with group meetings (“Group Session”) and moderate to vigorous aerobic exercise (“MVAE”) as well as the supporters’ session in week 5. The group sessions integrate different behavioral change techniques (“BCT”) to enhance motivational and volitional skills with the long-term aim for maintenance aerobic exercise. The medium gray boxes illustrate non-supervised aerobic exercise in which the patients can choose independently any aerobic exercise that best fits to their interests and needs. The light gray box illustrates the non-supervised group sessions from week 5 to 24 in which patients complete the aerobic exercise together but without the therapist. The telephones cartoons represent telephone contacts during the non-supervised time to monitor the long-term maintenance of aerobic exercise. The entire program is supported by the ImPuls smartphone application, developed especially for ImPuls.

### ImPuls smartphone application (“ImPuls-App”)

The ImPuls App will support the participants with options for exercise planning (training plans), exercise guidance (interval training, resistance program, ratings of perceived exertion), self-monitoring of goal achievements (analysis and feedback of FITT criteria and goal achievements), mitigation of barriers and a knowledge base. A web-based application for exercise therapists will support planning and logging individual and group sessions with clients (calendar, attendance, active participation, notes). Furthermore, the participants can share some or all of their data (such as their exercise schedule or their plans for overcoming barriers) with their exercise therapists via the secure channels between the ImPuls App and the web-based platform. This will enable direct feedback of therapists to their clients. Both Apps run on Google Android and on Apple iOS. All data generated in the apps will be protected by encryption on the smartphone. All communication between smartphones (ImPuls App), browsers (Therapist’s App) and the central server will be protected by established encryption protocols, too. All components in table P1 will be documented in or provided by the ImPuls App.

**Table P1.**
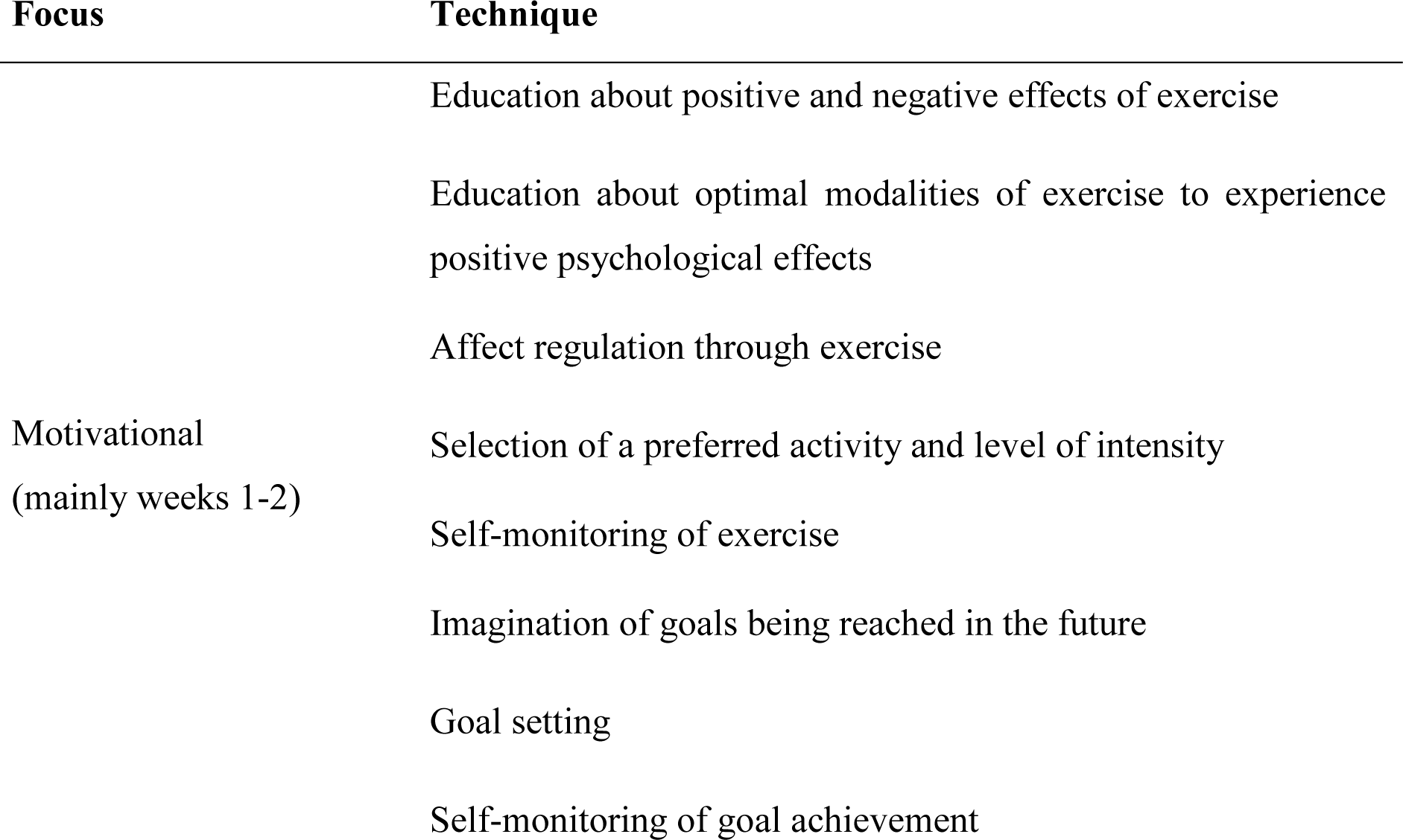
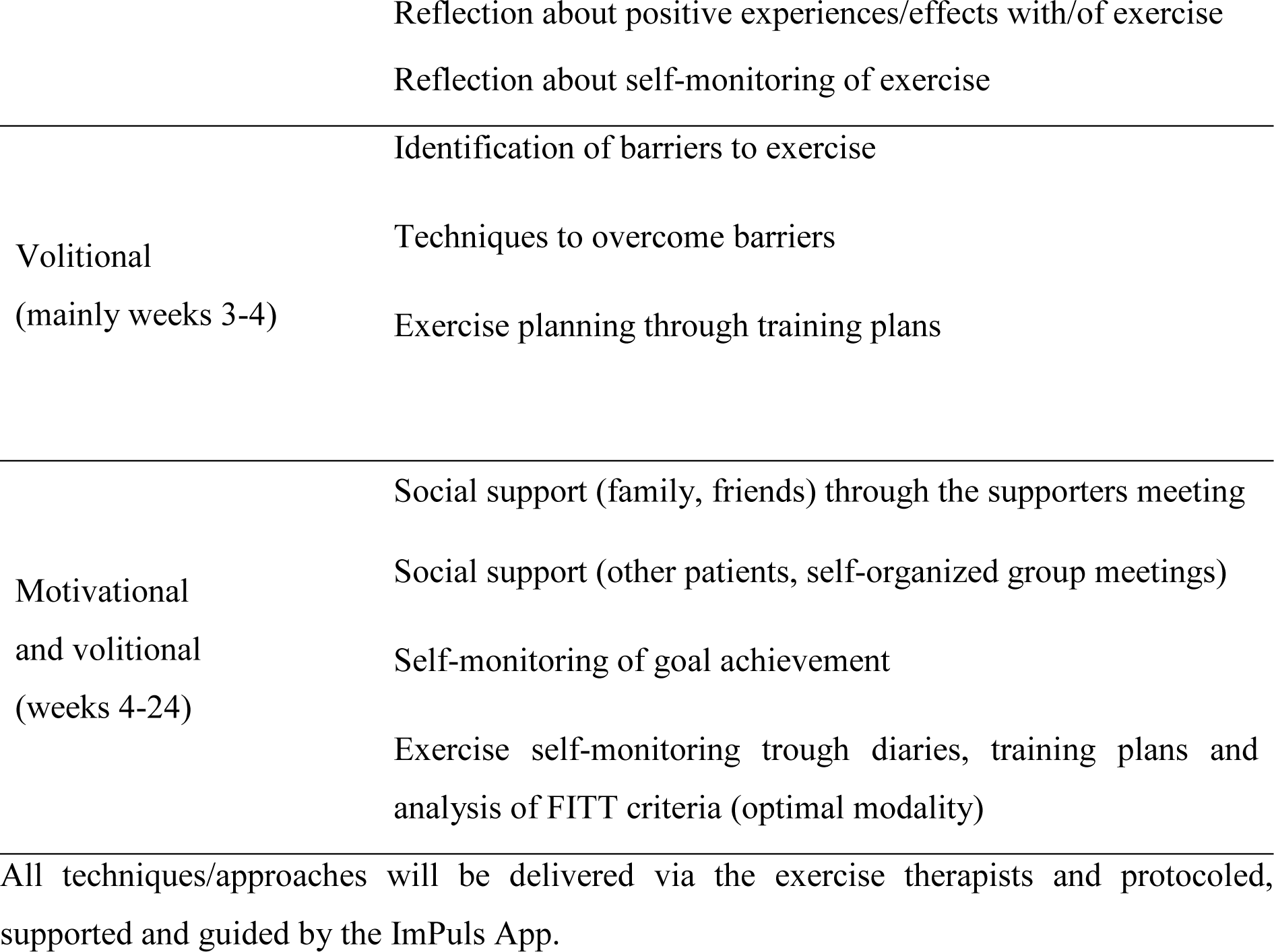
Overview of behaviour change techniques included in ImPuls.

#### Criteria for discontinuing or modifying allocated interventions

Participants who miss more than four consecutive sessions during the four weeks (two consecutive weeks) of the supervised phase (Weeks 1-4) of the intervention (≥40 %, due to any reason) but continue the study assessments are defined as treatment dropout, as they did not receive the minimum intended dose of the intervention. They can still participate in the remaining sessions of the intervention and will be asked to complete all remaining assessments. A treatment dropout is only counted in the time before an official study dropout (e.g., if a patient completely drops out of the study in Week 1, the patient is indicated as a study dropout and not as treatment dropout). If patients never attended treatment sessions but still take part in the assessments (are not defined as study drop-out), they are solely defined as treatment dropouts.

Participants who intentionally drop out of the entire study, are no longer accessible, or never/no longer participate in assessments are defined as study dropouts. It is always necessary to ask for the exact time when the patient decided to drop out of the study. Participants who intentionally discontinue the treatment or did not receive the minimum intended dose of the intervention (defined as treatment dropouts) and discontinue assessments will be also defined as study dropout. All participants agree to be asked to specify their reasons for study/treatment-dropout on a voluntary basis (once, after treatment/study dropout). If available, reasons for discontinuation will be reported. If more than 50 % of participants discontinue their participation in the intervention group during the first five weeks (Weeks 0-4) of the delivery, discontinuation of the group due to lack of economic efficiency for the study sites is possible. If only one participant of the intervention group remains, he or she can no longer receive the intervention, as delivery of the intervention to less than two participants no longer qualifies as “group-based intervention”.

All participants, including dropouts, will be asked to complete all of the following study assessments on a voluntary basis.

#### Strategies to improve adherence to intervention protocols, and any procedures for monitoring adherence

##### Intervention

To ensure adequate delivery of the intervention, all exercise therapists were trained in the ImPuls program (see eligibility criteria). All exercise therapists are offered to take part in regular supervisions by a licensed exercise therapist or supervisor in cognitive-behavioral therapy, who are also trained in the ImPuls program by the study PI. All intervention sessions will be video-taped. 10 % of the recorded sessions (randomly selected from pre-defined core-sessions) will be evaluated for treatment-fidelity by trained assessors. Every participant will set a 30-minute timer to monitor the duration of the supervised physical activity sessions. During the physical activity, intensity will be monitored through heart-rate monitors (w0-w4) as well as RPE (w0-w24). RPE will be assessed through automatic reminders of the smartphone application after 5, 15 and 25 minutes of physical activity.

The participants’ attendance will be monitored through participation lists. If participants miss a session of the intervention, they will be asked to complete 30 minutes of moderate to vigorous physical activity on their own. They will be sent the material of the patient training session. All participants are asked to schedule and track their physical activity sessions performed during the supervised as well as unsupervised phase through the smartphone application. They will receive automatic reminders to perform the scheduled physical activity. Exercise therapists will be able to access these activity logs and monitor the physical activity after the supervised phase (w0-w4) through phone contacts. If participants fail to meet their goals, exercise therapists will revise the activity goals and measures to overcome obstacles with the participants. Frequency (at least twice a week), intensity (mean intensity of least 64 %) and duration (at least 30 minutes each session) during the supervised period (4 weeks), will be assessed via the ImPuls smartphone application.

### Exercise therapists/study therapists

Study therapists are instructed to call patients in the case of absence or illness. In the case of sickness of study therapists within week 0-4 of the intervention, they can be represented by another trained therapist. In case of absence of more than 2 weeks, study therapists should be replaced be another trained therapist.

Study therapists are not allowed to take leave in week 1-4 of the intervention. In week 5 to 8 therapists should take leave for max. 1 week. Weeks 9 to 24 there are no rules regarding taking leave. In case of leave study therapists have to be represented by another trained exercise therapist.

### Assessments

Online questionnaires and the assessment of physical activity will be carried out within a period of 14 days. To ensure a good completion rate of the online surveys, automatic reminder emails, with the request to complete the questionnaires, are sent to the participants after a fixed number of days: emails were sent to the participants 5 days after receiving the survey invitation for the prep, pre, rando, inter, post and follow-up assessments and 3 days after receiving the invitation for the weekly assessments. In case surveys were still not completed, automatic emails to inform the responsible research assistants at the University of Tuebingen were sent 7, 8 and 10 days after the dispatch of the survey invitation for prep, pre, rando, inter, post and follow-up assessments. On the 7^th^ day the University of Tuebingen calls the patient, exercise therapist or manager who did not respond. If the 6^th^ day is a Saturday the University of Tuebingen calls the patient, exercise therapist or manager on the 8^th^ day.

For the weekly assessments (week one through 12), participants have 4 days to fill out questionnaires, which are sent on Saturday. On Monday: the participants are reminded to fill out the questionnaires via e-mail. If participants do not respond two weeks in a row, the LMU informs the University of Tuebingen about missing response on Tuesday morning. The participants are then contacted per telephone. If participants do not respond for three weeks in a row without dropping out, participants are contacted again. If participants do not respond for four weeks in a row, participants are not contacted via telephone until post. If participants respond to one weekly questionnaire after a period of non-response the procedure described above will be applied.

For the assessment after randomization (week 0) and the monthly assessments in the unsupervised intervention phase (week 16, week 20, week 24) participants have 7 days to fill out questionnaires, which are sent on Saturday. On Monday the participants are reminded to fill out the questionnaires via e-mail. The LMU informs the University of Tuebingen about missing response on Tuesday and Thursday morning. The participants are then contacted per telephone.

#### Relevant concomitant care and interventions that are permitted or prohibited during the trial

ImPuls will be delivered in addition to standard care/treatment as usual (TAU). Any other standard treatment (psychological or pharmacological treatment) covered by German health insurances are permitted.

#### Primary, secondary, and other outcomes

##### Primary Outcome

###### Global symptom severity

Global symptom severity will be assessed by the Global Severity Index (GSI) of the German version of the Brief Symptom Inventory [BSI-18].^69–71^ The GSI reflects the general mental distress rating on the symptom scales somatization, depression, and anxiety. Each symptom scale consists of 6 items. Thus, 18 items are rated on a 5-point Likert scale (range: 0-4). Higher scores indicate higher distress. Cut-off scores were evaluated separately for men (≥ 10) and women (≥ 13) and have high sensitivity (91.2 %) and specificity (92.6 %).^72^ Among patients with affective disorders, the GSI has demonstrated good internal consistency (α = .89) and construct validity (r = 0.71). Among patients with anxiety disorders the BSI-18 has an internal consistency of Cronbach’s α = .88 and a construct validity of r = 0.67.^73^

##### Secondary Outcomes

###### Major depressive disorder

The secondary endpoint *depressive symptoms* will be assessed with the PHQ-9 module, assessing symptoms over the last two weeks with nine items, each of them representing one of the DSM-5 (Diagnostic and Statistical Manual of Mental Disorders) criteria for a depressive episode.^74,75^ All items are rated on a 4-point Likert scale (range: 0-3). The sum of all items represents the total score (range: 0-27). Higher scores indicate higher levels of depression. Regarding depressive symptomology, individuals are classified according to the degree of depression severity: absence of depressive disorder (0-4), mild degree of severity (5-10), medium major depression (10-14), severe major depression (15-19) and most severe major depression (20-27). In medical settings the cut-off of ≥10 is used to detect a major depressive disorder.^74,76^ This cut-off was shown to have a sensitivity and specificity of 88 % and 85 %, respectively.^77^ The scale measuring depressive symptoms has an internal consistency of Cronbach’s α = .87 among a representative German sample.^74,78^

###### Insomnia / Sleep quality

Nonorganic insomnia will be assessed with the German version of the Insomnia Severity Index [ISI].^79,80^ The ISI consists of seven items and assesses the severity of sleep onset difficulties, sleep maintenance difficulties, early morning awakening, satisfaction with current sleep, interference with daytime functioning, noticeability of impairment attributed to sleep problems and degree of distress or concern caused by the sleep problem of the past two weeks. The total score ranges from 0 to 28 (range of component scores: 0-3), with a higher score reflecting greater insomnia severity. The cut-off score of ≥ 11 has shown a high sensitivity (91.4 %) and specificity (84.4 %) in identifying insomnia. The ISI has shown an internal consistency of Cronbach’s α = .83 among a representative German sample.^79^

Sleep Quality will be assessed with the global sleep quality score of the German version of the Pittburgh Sleep Quality Index [PSQI].^81^ The global sleep quality score is the sum of seven sleep component scores (range of component scores: 0-3): subjective sleep quality, sleep latency, sleep duration, habitual sleep efficiency, sleep disturbances, use of sleeping medications, and daytime dysfunction. The global sleep quality score can vary from 0 to 21 with a cut-off score of 5, identifying clinically raised sleep impairment.^81^ It has shown a high sensitivity (98.7 %) and specificity (84.4 %) in identifying insomnia.^82^

###### Panic disorder and Agoraphobia

The seven-item Generalized Anxiety Disorder scale [GAD-7] assesses symptom severity of generalized anxiety during the last 2 weeks, however shows good performance as a screening measurement for panic disorder and agoraphobia and will therefor serve as a measure for panic agoraphobia symptoms.^83–86^ Items are rated on a four-point Likert scale (range: 0-3). The sum of all items represents the total score (range: 0-21), with scores of ≥ 5 representing mild, scores of ≥ 10 moderate and scores of ≥ 15 severe anxiety symptom levels, respectively. The cut-off score of ≥ 10 has shown high sensitivity (89 %) and specificity (82 %).^86^ Among a representative German sample, the GAD-7 has an internal consistency of Cronbach’s α = .85.^87^

Besides the GAD-7, symptoms of panic disorder and agoraphobia symptoms will be assessed with the three-items subscale panic of the 6-items scale anxiety of the BSI-18.^69^ Current evidence suggests a four-factor structure of the BSI-18 that retains the somatization and depression symptom scales but splits the anxiety symptom scale in two factors: General anxiety and panic.^88,89^ The subscale panic of the BSI-18 consists of three items that are rated on a five-point Likert scale (range: 0-4). Among a German outpatient sample that was surveyed five times after the 2^nd^, 6^th^, 10^th^, 18^th^ and 26^th^ therapy session the best fitting model according to Akaike Information Criterion (AIC) was always the model with four factors, compared to one- and three-dimensional models.^88^ Among patients with anxiety disorders, the GSI of the symptom scale anxiety has demonstrated good internal consistency (α = .83) and construct validity (r = 0.67).^73^

###### Posttraumatic Stress Disorder

To assess symptoms of PTSD, the German version of the PTSD Checklist for DSM-5 [PCL-5] will be used.^90^ The questionnaire is a self-report measure that consists of 20 items corresponding to the DSM-5 criteria for PTSD. Participants report their intensity of symptoms over the past four weeks on a five-point Likert scale (0 = not at all to 4 = extremely; total range 0-80). Higher scores indicate higher levels of PTSD. The German Version shows high internal consistency (α = .95), high test-retest reliability (r = .91) and a high construct validity (r = .77). A cut-off of 33 indicates clinically relevant symptomatology.

###### Health related quality of life

Health related quality of life will be assessed by the German version of the EQ-5D-5L questionnaire.^91,92^ It consists of five items concerning the domains mobility, self-care, usual activities, pain or discomfort and anxiety or depression with five answer alternatives each (range: 1-5). The combinations of the answer alternatives can be described with a five-digit number (i.e. the pattern 11111 indicates the optimal health state). Through the EQ-5D-5L questionnaire Quality Adjusted Life Years (QALY) are captured for the economic evaluation. The EQ-5D-5L has an internal consistency of Cronbach’s α = .86 among German chronic heart failure patients. Current data concerning internal consistency in patients with mental disorders exists for the Spanish version of the EQ-5D-5L. Among Spanish patients with major depression the EQ-5D-5L has an internal consistency of Cronbach’s α = .77.^93^

###### Routine data of the health insurances/health care costs

For the economic evaluation, patients’ routine data collected 6 months before the intervention, during the time of the intervention, and six months after the intervention will be provided by the two participating statutory health insurers (AOK and TK). This data will include patients’ master data, such as gender and age, as well as patient treatment costs. Parameters for treatment costs will comprise costs of inpatient and outpatient care as well as medication, medicals aids or days of incapacity to work. Routine data for each patient will be provided for the time of the intervention as well as one year prior and one year after. The relevant costs are assessed and aggregated as quantities. In addition, cost parameters resulting from the intervention and the implementation will be considered. Subsequently, routine insurance data will be linked to primary data collected.

### Change of health insurance during study participation

In the case of a change of health insurance company during study participation, the corresponding participant is still part of the study and continues to complete the assessments. However, routine data about the health insurances/health care costs cannot be provided.

#### Exercise behavior/MVAE

The assessment of self-reported exercise duration and frequency and the assessment of accelerometer -based moderate to vigorous physical activity will serve as two proxies for MVAE. Exercise in minutes per week will be assessed using the self-report Exercise Activity Index of the Physical Activity, Exercise, and Sport Questionnaire [BSA questionnaire].^94^ Participants specify type, duration, and frequency of exercise in the last four weeks. A weekly average of more than 200 minutes of exercise will be checked again to see if it makes sense and in the event of a mistake (i.e., repeated entry of an activity already stated in the leisure time index) it will be defined as missing; in the event of an unrealistic entry, it will be defined as missing as well.

Moderate to vigorous physical activity (MVPA) will be assessed via accelerometer-based sensors (Move 4, movisens GmbH). The sensor assesses physical activity of a person based on kinematic data in three dimensions and atmospheric air pressure. This allows to estimate the amount of physical activities of different intensities for a specified time period based on validated algorithms.^95^ Patients will wear the sensors on the hip for seven consecutive days. A sensor record is considered valid and used for analysis if the sensor was worn for eight hours on at least four out of seven days.^96^ In addition, the participants complete a protocol, in which non-wear time or other problems will be indicated. In case patients indicate medical problems that resulted in sedentary behavior, data will be set manually to N/A. Raw data will be checked, if non-wear time indicated by the participants is identified by the sensors. The data of valid days (at least eight hours wearing time) are added up and divided by the number of valid days and multiplied by 7.^97^ If a person has less than four days with eight hours of sensor data each, then the data for that whole week is set to N/A and will be imputed. Volume of moderate to vigorous physical activity will be indicated in minutes/week and calculated by the daily physical activity of at least moderate intensity (≥ 3 MET)/minutes.

#### Sociodemographic data

The following sociodemographic data will be assessed during the pre-assessment: gender, age, marital status, living situation, having children, nationality status, mother tongue, highest level of education, current work situation, duration of current mental health problems, use of online therapy programs or therapy applications, use of programs to increase physical activity, current psychiatric treatment, use of psychiatric medication (current and in the past), previous physical and psychological disorders, outpatient psychological treatment (current and in the past) including number and time period, previous inpatient treatment because of psychiatric problems including number and time period, previous day-care treatment including number and time period. Due to technical reasons the variable current psychological treatment at pre-assessment needed to be reassessed by several patients at post-assessment. Therefore, we have several missing answers to this question at pre-assessment.

#### Assessments to ensure internal validity

##### Symptom severity at randomization

Symptom severity might change or fluctuate from the diagnostic interview or pre-assessment to the start of the intervention. In order to ensure clinically relevant transdiagnostic and disorder-specific symptomatology with the start of the intervention, scales that assess all primary and secondary outcomes will be presented again between randomization and group start: BSI-18, ISI, PSQI, PHQ-9, GAD-7, PCL-5.^69,74,75,79–81,85,86,90^. Participants will have 7 days to answer the questionnaires. An ImPuls group have to start at maximum 14 days after randomization.

##### (Serious) Adverse Events

Adverse events (AE) will be assessed at pre, post-1, and follow-up assessment. AEs and Serious Adverse events (SAEs) can be further reported by patients or therapists at a central phone number. At each measurement point SAEs will be assessed by structured interviews. AEs and SAEs will be documented and SAEs will be reported to an independent Data Safety and Monitoring Board, which will discuss adjustments to or discontinuation of the entire study.

##### Treatment Fidelity (adherence to protocol of study therapists)

Core elements of the manualized ImPuls intervention have been determined a priori. Separate rating forms will be developed for each session. Items will be divided into general adherence questions (e.g., “therapist discusses goals of the session”) and questions focusing on adherence regarding the core elements. All in-house sessions (see Fig. P3 and Table P1) from all therapists at all study sites will be video-taped. Outdoor MVAE within one session will not be recorded. One session video for each group out of eight recorded core sessions (sessions that comprised of intervention core elements) will be chosen for fidelity evaluation. This corresponds to 12.5 % of all core sessions and 10 % of all recorded sessions. Randomization will be done by the unblinded data manager with Excel (Microsoft Excel Version 1808 - (Microsoft Office Professional Plus 2019)). Two trained research assistants will rate the sessions with regard to adherence to the treatment manual. Raters were trained by the developers of the intervention. During the training, items of rating sheets for each session will first be explained and discussed and a test video of Session 1 will be rated collaboratively with the trainer to illustrate the procedure. In a second training step, the two raters independently rate test videos from the remaining seven core sessions, whereby test videos had previously been randomly selected and were different from the videos used later to establish adherence. Finally, a debriefing session with the raters and developers of the intervention will take place, where remaining questions will be discussed and clarified. After completion of the training, the two raters independently rate the randomly selected videos for the fidelity evaluation, with no communication between the two raters nor with the trainer or developers of the intervention. Adherence to the treatment manual will be assessed by rating with “yes” (presence) or “no” (absence) as to whether pre-defined elements of the ImPuls intervention were delivered. A general adherence score and an adherence score focusing only on core elements will be calculated and averaged to calculate the overall fidelity score, which is the percentage of all items answered with “yes”. An overall inter-rater reliability score will be calculated and reported. The overall fidelity score was assumed to be of ≥ 90 %.

##### Drop-out rate

Participants who miss more than four consecutive sessions during the four weeks (2 consecutive weeks) of the supervised phase (Weeks 1-4) of the intervention (≥40 %, due to any reason) but continue the study assessments are defined as treatment dropout, as they did not receive the minimal intended dose of the intervention. They can still participate in the remaining sessions of the intervention and will be asked to complete all remaining assessments. A treatment dropout is only counted only in the time before an official study dropout (e.g., if a patient completely drops out of the study in Week 1, the patient is indicated as a study dropout and not as a treatment dropout). If patients never attended treatment sessions but still take part in the assessments (are not defined as study dropout], they are solely defined as treatment dropouts.

Participants who intentionally drop out of the entire study, are no longer accessible, or never/no longer participate in assessments are defined as study dropouts. It is always necessary to ask for the exact time when the patient decided to drop out of the study. Participants who intentionally discontinue the treatment or did not receive the minimum intended dose of the intervention (defined as treatment dropouts) and discontinue assessments will be also defined as study dropout. All participants agree to be asked to specify their reasons for study/treatment-dropout on a voluntary basis (once, after treatment/study dropout). If available, reasons for discontinuation will be reported. If more than 50 % of participants discontinue their participation in the intervention group during the first five weeks (Weeks 0-4) of the delivery, discontinuation of the group due to lack of economic efficiency for the study sites is possible. If only one participant of the intervention group remains, he or she can no longer receive the intervention, as delivery of the intervention to less than two participants no longer qualifies as “group-based intervention”.

##### Attendance rate

Attendance within the supervised (Weeks 0-4) and partially supervised (Weeks 5-24) period will be assessed using attendance lists by the exercise therapists/study site. If the supporter’s session or a telephone contact for the whole group did not take place in a week, these appointments are not included in the attendance rate as people’s missed appointments, but are counted as appointments that never took place. Only attendance before an intentional study dropout counts in the attendance rate. Treatment dropouts are irrelevant for the attendance rate as it is defined as dose reach, therefore attendance of treatment dropouts will be calculated into the attendance rate. If patients never participated but still take part in the assessments, their attendance will not be considered for the attendance rate.

##### MVAE dose within the supervised period (patients)

Frequency (at least twice a week), intensity (mean intensity of least 64 % of maximum heart rate (HRmax) / subjective rating of perceived exertion) and duration (at least 30 minutes of MVAE in each session) of exercise during the supervised period (weeks 1-4) will be assessed via the ImPuls smartphone application. Individual heart rate will be tracked by heart rate monitors. Since heart rate trackers report only mean heart rate and not mean percentage of HRmax after an exercise session, patients enter their mean HR in the ImPuls smartphone app after each session. Percentage of HRmax will be calculated by the research team for further analysis. Maximum heart rate will be calculated by subtracting age from 220.^67,68^

##### Expectations, Motivation and Satisfaction

Validated scales adapted for use in the context of ImPuls will be employed to assess patients’ outcome expectations, motivation and satisfaction with the intervention, as well as exercise therapists’ motivation, and satisfaction with the intervention.^98–102^ On all scales, means falling into the upper quartile are considered high, means falling into the second and third quartiles are considered moderate, and means falling into the lower quartile are considered low trait expression.

### Process Evaluation

Exercise therapists, managers, referrer, and patients receive *online questionnaires* through the web-based data management system REDCap at different time points (see Figure P1, Tables P 2-6).^103,104^ All participants receive an individual web-link via E-mail to access the online survey and have 2 weeks to complete it (except weekly assessments during inter phases 1 - 2, where patients have only 1 week to complete it). Reminders are automatically sent out 5 days after receiving the survey invitation (for weekly assessments: 3 days). We primarily used existing and already validated (in German) measures. If these were not available in German, we translated them using established backward translation procedures utilizing native speakers. In order to gain a more profound insight into the processes of the ImPuls intervention, in some measurements of the exercise therapists, managers and physicians we adapted the items specifically to ImPuls or developed items ourselves. Moreover, recruitment strategies allow for a variety of healthcare professionals (e.g., psychotherapists and primary care physicians) to refer patients to ImPuls. For this purpose, we ask them via online questionnaires about their opinion regarding exercise in combination with behavior change techniques as a new treatment option for patients with mental disorders.

A *guided semi-structured interview* was developed with respect to aspects of the MRC framework (e.g., acceptability, fidelity/delivery), empirical considerations prior to the intervention and questions that arose over the course of the intervention (e.g., in conversations with exercise therapists).^105^ During the interviews, we ask exercise therapists about their experiences with patients in the ImPuls groups they conducted, their opinions about the program content regarding motivational and volitional BCTs and exercise, the perceived applicability of the program (regarding target group, general conditions in the outpatient rehabilitative and medical care facilities, ImPuls smartphone application and their own qualification) and their opinion on a possible long-term implementation of the ImPuls intervention. Possible reports of specific difficulties in implementing the program can provide us with information on why they may have had to deviate from the manual (adherence). Additionally, it can inform us regarding the areas in which they should have received more training. In summary, the interview focuses on facilitators and barriers for the implementation of the ImPuls intervention from exercise therapists’ perspectives. Interviews are conducted face to face by researchers of the process evaluation team with 20 exercise therapists, who all conducted at least one ImPuls group. The interviews have an estimated average duration of 50 minutes.

A *focus group interview* was developed analogous to the procedure mentioned above. It is supposed to provide an in-depth insight into managers’ perspectives concerning the perceived barriers and facilitators regarding the feasibility of the ImPuls intervention in the outpatient setting and its possible long-term implementation in the future. The focus group interview is conducted with 10 managers and is estimated to last 120 minutes.

Face to face interviews as well as the focus group interview are conducted once there are no further ImPuls groups in the respective outpatient rehabilitative and medical care facility. All interviews will be audio-recorded. Subsequently, the audio-records are saved on a secured network drive of the University of Tübingen and transcribed verbatim by research assistants. All mentions of personal data are masked during the transcription.

Researchers from the process evaluation team evaluate the interviews in an deductive-inductive process following the steps of a qualitative content analysis.^106^

Data from the *ImPuls smartphone application* is collected continuously during the supervised and partially supervised phase (inter 1 – 3; see figure P2 and table P1). It shows the extent to which patients use the ImPuls smartphone application (frequency) in general as well as regarding different application functions including goal setting, barrier management and training plans (patients‘ integration of core components). Repetitive negative thinking and valence of affect are measured with a self-developed self-assessment manikin prior to and after each supervised and unsupervised exercise session over the entire intervention period. We also collect data from the *web-based ImPuls interface* accompanying the ImPuls smartphone application to record the extent to which exercise therapists have used the tool to review patients’ shared information during supervised and partially-supervised phases.

We receive *documentation data* from the exercise therapists and from the project staff. Exercise therapists are required to document whether all scheduled inhouse sessions (supervised phase) and phone calls (partially supervised phase) were offered or completed. As part of the recruitment process, project staff document how patients became aware of the project, how patients are distributed among outpatient rehabilitative and medical care facilities and patient dropouts and their reasons before and during the study. Recruitment strategies include flyers and posters at the offices of the participating outpatient rehabilitative and medical care facilities, primary care physicians, psychotherapists and physiotherapists as well as a direct approach by health insurers involved in the project. We also disseminate information about the ImPuls intervention through self-help groups, daily newspapers, magazines, student mailing lists and social media.

**Table P2.** primary and secondary outcomes for patients at each time point (following SPIRIT template).

|  | pre | inter-1 | inter-2 | inter-3 | post | follow-up |
| --- | --- | --- | --- | --- | --- | --- |
| **Primary Outcomes** |  |  |  |  |  |  |
| Global symptom severity [BSI-18]<sup>69</sup> | x |  |  |  | x | x |
| Global symptom severity subscales: depressive symptoms, anxiety symptoms |  | 1x/week | 1x/week | 1x/month |  |  |
| **Secondary Outcomes** |  |  |  |  |  |  |
| Depressive symptoms [PHQ-9]<sup>74,75</sup> | x |  |  |  | x | x |
| Nonorganic insomnia [ISI]<sup>79,80</sup> | x |  |  |  | x | x |
| Sleep Quality [PSQI]<sup>81</sup> | x |  |  |  | x | x |
| Panic disorder and Agoraphobia [GAD-7; BSI-18, subscale panic]<sup>85,86</sup> | x |  |  |  | x | x |
| Posttraumatic stress disorder [PCL-5]<sup>90</sup> | x |  |  |  | x | x |
| Health related quality of life [EQ-5D-5L]<sup>91,92</sup> | x |  |  |  | x | x |
| Routine data / health care costs | x |  |  |  | x | x |
| Exercise behaviour via self-report [BSA-F]<sup>94</sup> | x |  |  |  | x | x |
| Moderate to vigorous physical activity via accelerometer-based sensors (Move 4, movisens GmbH) | x |  |  |  | x | x |
| *Note.* Inter-1 = during the supervised ImPuls phase (week 1-4); inter-2 = during the partially supervised ImPuls phase 2 (week 5-12); inter-3 = following the partially supervised ImPuls phase 2 (week 13-24) post = after completion of the intervention (supervised and partially supervised) ImPuls phases 6 months after randomization; pre = prior to randomisation. |  |  |  |  |  |  |

**Table P3.** Assessments for exercise therapists at each time point.

| ASSESSMENT EXERCISE THERAPISTS | TIMEPOINT |  |  |  |  |  |  |  |  |
| --- | --- | --- | --- | --- | --- | --- | --- | --- | --- |
|  | prep-1 | prep-2 | pre | inter-1 | inter-2 | inter-3 | post | follow-up | post-d |
| **Implementation** |  |  |  |  |  |  |  |  |  |
| Training – satisfaction [modified]<sup>107</sup> | x | x |  |  |  |  |  |  |  |
| Training – perceived skill acquisition / self-efficacy (ImPuls intervention) [adapted to Impuls]<sup>108-110</sup> | x | x |  |  |  |  |  |  |  |
| Supervision - participation rate |  |  |  | x |  |  | x |  | x |
| Supervision – satisfaction [modified]<sup>107</sup> |  |  |  | x |  |  | x |  | x |
| App - user frequency (App) |  |  |  | x | x | x |  |  |  |
| Adherence (video) |  |  |  | x |  |  |  |  |  |
| Quality of delivery (video) |  |  |  | x |  |  |  |  |  |
| Dosage - amount sessions delivered (documentation) |  |  |  | x | x | x |  |  |  |
| **Mechanisms of impact** |  |  |  |  |  |  |  |  |  |
| Global self-efficacy ImPuls [adapted to Impuls]<sup>109</sup> |  |  | x |  |  |  |  |  | x |
| Attitudes towards mental disorders (OMS-HC) [translated + modified]<sup>111</sup> |  |  | x |  |  |  |  |  | x |
| Attitudes towards manualized interventions (EBPAS-36 D)<sup>112</sup> |  |  | x |  |  |  |  |  |  |
| Motivation towards conducting ImPuls (SESSW)<sup>102</sup> |  |  | x |  |  |  | x |  |  |
| Motivation working with patients with mental disorders (self-developed items)<sup>1</sup> |  |  | x | x |  |  | x |  |  |
| Expectation of program success (PATHEV)<sup>100</sup> |  |  | x |  |  |  |  |  |  |
| Program acceptance/satisfaction (B&F-A) [translated + modified]<sup>113</sup> |  |  | x |  |  |  |  |  | x |
| Perceived barriers (B&F-A) [translated + modified]<sup>113</sup> |  |  |  | x | x |  | x |  |  |
| Satisfaction with the ImPuls group, WAI-SR [modified]<sup>107,114</sup> |  |  |  | x |  |  | x |  |  |
| Coping strategies (DPCCQ) [modified]<sup>115</sup> |  |  |  | x |  |  | x |  |  |
| App-usability (SUS) [translated + modified]<sup>116</sup> |  |  |  | x | x |  | x |  |  |
| App-functionality (MARS-G)<sup>117</sup> |  |  |  |  |  |  | x |  |  |
| App-viability (self-developed) |  |  |  |  |  |  | x |  |  |
| App-satisfaction (MARS-G)<sup>117</sup> |  |  |  | x | x |  | x |  |  |
| Context |  |  |  |  |  |  |  |  |  |
| Demographics | x |  |  |  |  |  |  |  |  |
| Qualification (self-developed) | x |  |  |  |  |  |  |  |  |
| Therapeutic experience (regarding exercise in group setting /with patients with mental disorders) (self-developed) | x |  |  |  |  |  |  |  |  |
| *Note:* follow-up = once after 12 months; Inter-1 = once at the end of the supervised ImPuls phase (week 1-4); inter-2 = once at the end of the partially supervised ImPuls phase 1 (week 5-12); inter-3 = once at the end of the partially supervised ImPuls phase 2 (week 13-24); post = after completion of the intervention (week 24-26, supervised and partially supervised) ImPuls phases; post-d = end of study period in the outpatient facility; pre = prior to intervention start in the outpatient facility; prep-1 = following the first training; prep-2 = following the second training; rando = after randomization, prior to intervention start in the outpatient facility; <sup>1</sup>Pre = 7 items (3 items with regard to project participation); inter-1 & post-1 = 4 items (working with target group)<br>*Abbreviations:* B & F – A= Barriers and facilitators assessment instrument; DPCCQ = Development of Psychotherapist Common Core questionnaire; EBPAS -36D = Evidence based practice attitude scale (German version); FPTM-40= Therapy motivation questionnaire; MARS-G = Mobile App Rating Scale (German version); OMS-HC = Opening minds scale for Health Care Providers; PATHEV = Measurement of therapy expectation and therapy evaluation of patients; SESSW = Scale for recording subjective school values; SUS = System usability scale; WAI-SR = Working Alliance Inventory – short revised (German version). |  |  |  |  |  |  |  |  |  |

**Table P4.**
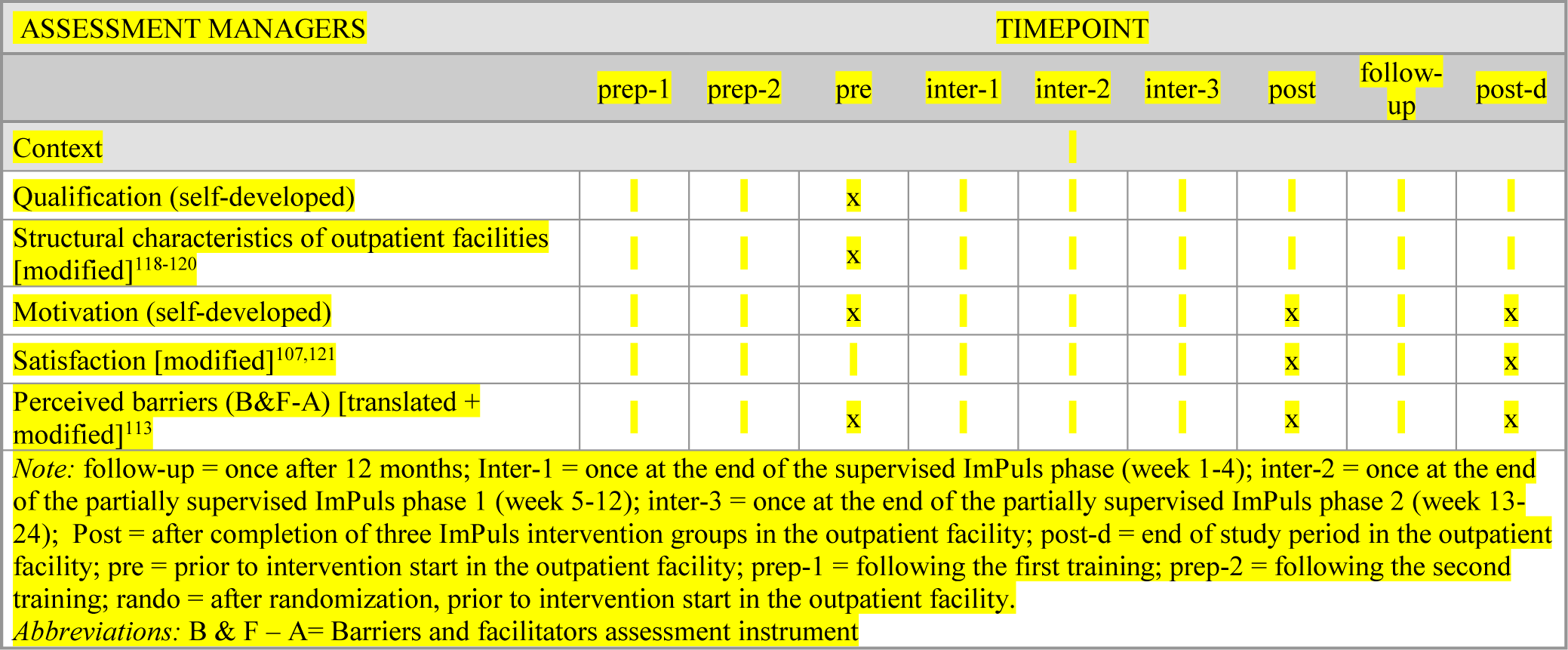
Assessments for managers at each time point.

**Table P5.**
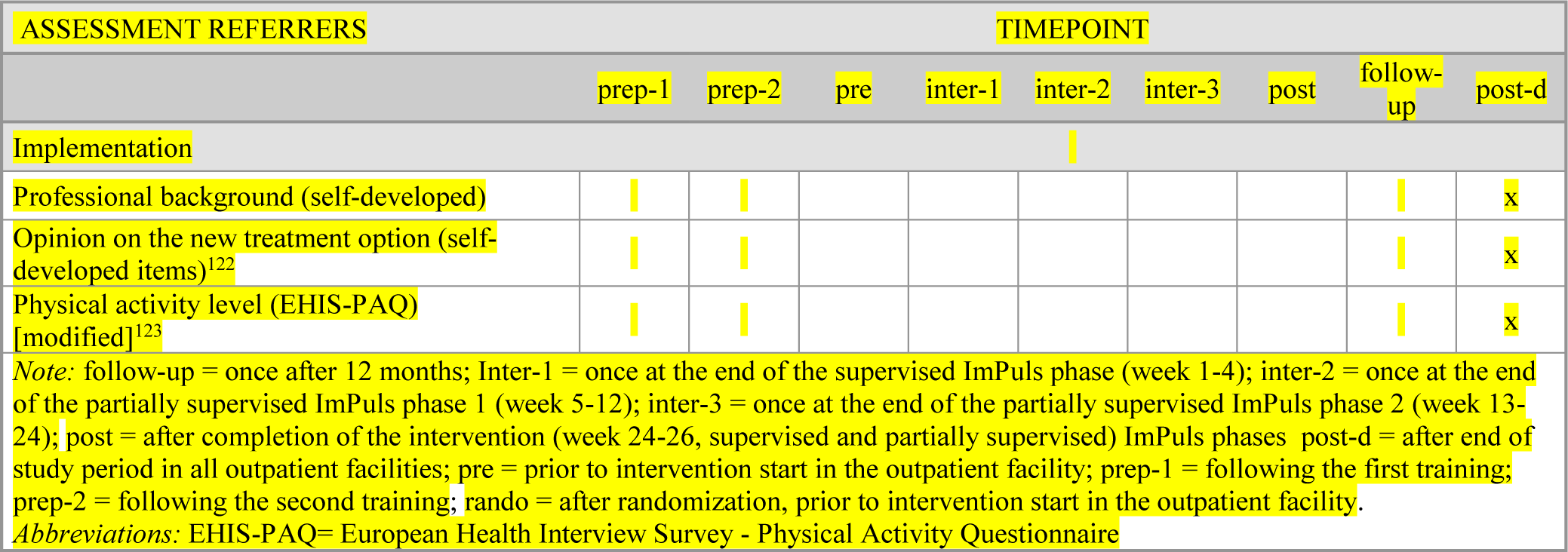
Assessments for referring health care professionals at each time point.

| ASSESSMENT REFERRERS |  | TIMEPOINT |  |  |  |  |  |  |  |  |
| --- | --- | --- | --- | --- | --- | --- | --- | --- | --- | --- |
|  |  | prep-1 | prep-2 | pre | inter-1 | inter-2 | inter-3 | post | follow-up | post-d |
| Implementation |  |  |  |  |  |  |  |  |  |  |
| Professional background (self-developed) |  |  |  |  |  |  |  |  |  | x |
| Opinion on the new treatment option (self-developed items)<sup>122</sup> |  |  |  |  |  |  |  |  |  | x |
| Physical activity level (EHIS-PAQ) [modified]<sup>123</sup> |  |  |  |  |  |  |  |  |  | x |

*Note:* follow-up = once after 12 months; Inter-1 = once at the end of the supervised ImPuls phase (week 1-4); inter-2 = once at the end of the partially supervised ImPuls phase 1 (week 5-12); inter-3 = once at the end of the partially supervised ImPuls phase 2 (week 13-24); post = after completion of the intervention (week 24-26, supervised and partially supervised) ImPuls phases post-d = after end of study period in all outpatient facilities; pre = prior to intervention start in the outpatient facility; prep-1 = following the first training; prep-2 = following the second training; rando = after randomization, prior to intervention start in the outpatient facility.
*Abbreviations:* EHIS-PAQ= European Health Interview Survey - Physical Activity Questionnaire

**Table P6.**
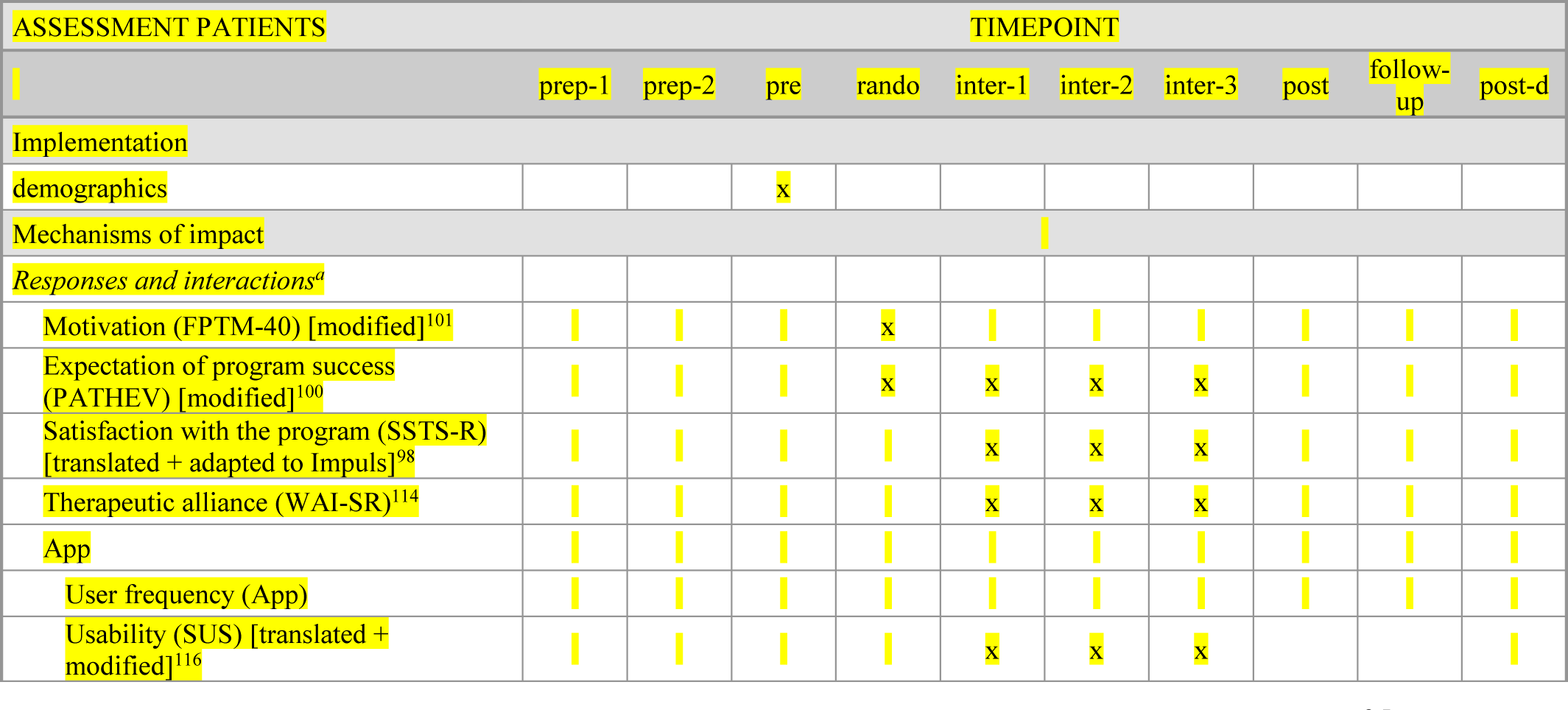
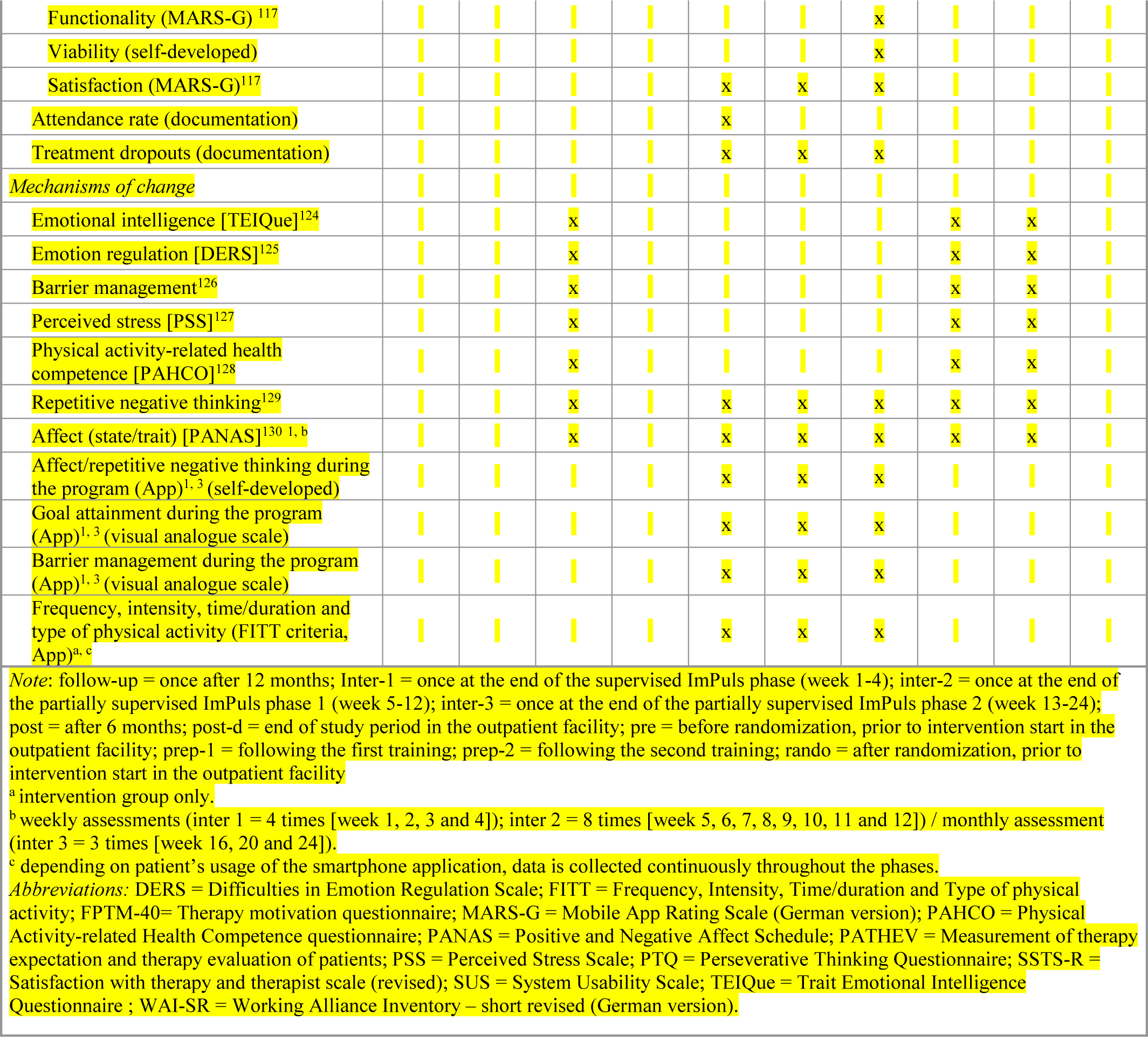
Assessments for patients at each time point.

**Figure P3:**
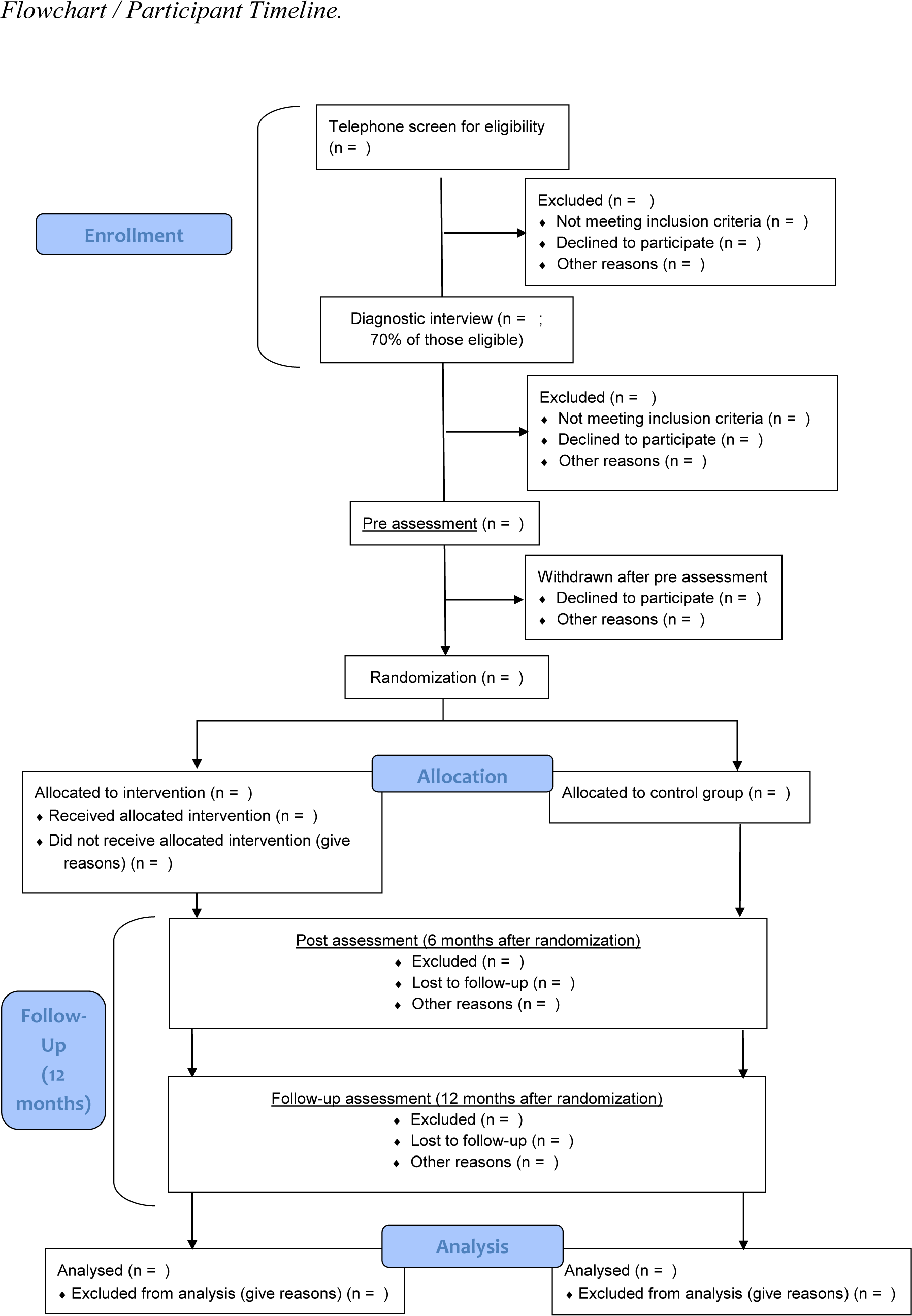
Patient Flow of the pragmatic randomized controlled trial in accordance with CONSORT

#### Estimated number of participants / Sample

The sample size was determined a priori using power analysis (G*Power, version 3.1.9.2).^131,132^ Power analysis was conservatively based on the lowest symptom-related post-treatment effect of exercise (vs. TAU/waiting list) on all included clinical disorders, namely the effect size of d = -0.348 (g = - 0.347) for symptoms of post-traumatic stress disorder.^28^ A two-sided *t*-test, alpha level of 0.05, a test power of 80%, an equal cell population, and a dropout rate of 30% were assumed. This calculation resulted in N = 375, which is conservative enough to detect the lowest expected treatment effect at the post-treatment phase. However, we regarded this sample size as the minimum and targeted a total sample of up to N = 600, in order to have enough statistical power for further analysis, e.g., looking at moderators of treatment effects. In April 2022 we decided to recruit the minimum sample size. Due to various measures to contain the COVID pandemic, recruitment was delayed and made more difficult, which is why the original target of 600 participants could not be reached.

#### Recruitment / strategies for achieving adequate participant enrolment to reach target sample size

Patients will be recruited mainly via inpatient psychiatric departments, family practices, general practitioners and psychiatric and psychotherapeutic outpatient units. The project will be conducted in collaboration with two health insurances, the AOK Baden-Württemberg (AOK) and the Techniker Krankenkasse (TK), who will support the recruitment with a targeted approach of general practitioners, psychiatrists and psychotherapists. All hospitals, clinics and practices will receive information material, such as flyers and posters, to inform their patients about the project. In addition, AOK and TK will publish articles in media distributed for their members during the course of the project. The TK further will inform eligible patients directly via phone calls. Recruitment will be additionally performed via social media posts (Instagram and Facebook), newsletters of professional associations, email distribution lists of universities, local influencers and regional newspapers and television.

Interested patients will first attend a preliminary telephone screening of eligibility criteria and will receive general information about the project (see also fig. 2). Patients will be screened for somatic contraindications for exercise via the Physical Activity Readiness Questionnaire (PARQ) and will be informed that they have to provide a physician referral for ImPuls before pre-assessment.^133^ In case of any suspicion of somatic contraindications, that might oppose participation (e.g., heart diseases or orthopedic problems), patients will be asked to provide an additional medical consult from their general practitioner or a medical specialist. During this screening, the recruitment strategy is assessed and recorded in the telephone lists in order to assess recruitment success per referrer. Institutions (i.e. clinic, psychotherapy practice, doctor’s practice) are evaluated before pure information material (distributed flyers, posters, table posters) if participants state that they were made aware of the ImPuls program by employees of these institutions. If participants state that they only received the flyer (without a doctor’s or psychotherapist’s consultation), the recruitment strategy is assigned to flyers as well as magazines, newspapers, radio reports, etc., respectively.

Eligible participants will be invited for a first inhouse meeting taking place in a study site close to their residence. Within the meeting, they will provide informed consent for study participation, receive information about the study site, create an individual pseudonym and will be screened initially for symptomatology related to the exclusion criteria to prepare for the structural diagnostic interview. Only scheduled telephone contacts (this excludes miscalls or information-only calls) and only completed interviews at study sites will be counted as telephone screenings and interview sessions, respectively. Discontinued or canceled interviews due to any reasons will be counted as exclusion after telephone screening. The telephone screening and initial interview will be performed by trained research assistants. Following this first inhouse meeting, psychologists with a M.Sc. degree undergoing a training in cognitive-behavior therapy, who will be trained by an external expert for structured clinical interviews, will conduct the structured clinical interview for DSM-5 (SKID-5-CV) to confirm eligibility.^134^ The interviewers were all trained by an external lecturer specialized in assessment using the structured clinical interview for DSM 5 (SCID-5). The entire training consisted of 2 x 3 days with a main focus on hands-on role play-based training for conducting the SCID. Teams of three were formed. The first person was the patient, the second the interviewer, and the third person acted as an external rater who also rated the items of the SICD. If the items did not match, critical items were discussed until there was 100 % agreement between raters. During the assessment phase, weekly supervision sessions with a licensed professional supervisor were held to discuss critical cases. In addition, the supervisor could be contacted at any time during or after a structured interview to discuss ambiguities in the inclusion and exclusion criteria.

Once six patients at the same site will be found to be eligible for participation, they will receive online questionnaires via the web-based data management system REDCap and a accelerometer-based Physical Activity sensors (MOVE 4; movisens GmbH) which will be worn for seven consecutive days (pre-assessment).^103,104^ The method used to collect sex/gender data is based on self-reports (female, male and other). Online questionnaires and the assessment of physical activity will be carried out within a period of 14 days. On Day 15, the six patients will be randomized as a group to either the intervention or control condition. In case of an assignment into the intervention condition, study sites will have to start the intervention within 14 days. Before the start of the intervention, global symptom as well as disorder-specific symptom severity will be assessed again to check whether participants meet the cut-off criteria for a mental disorder. In each study site, 60 patients are planned to be recruited and randomized, resulting in 10 allocations per site. The intervention group will complete the exercise intervention in addition to TAU, while the control group will receive TAU within the real-world outpatient mental health care setting in Germany. The TAU condition is intended to represent the typical treatment patients receive in the German outpatient health care system. Thus, patients will not actively be provided with any treatment but they will be allowed to receive any intervention that is available to them. The procedure of the pre-assessment phase will be repeated six months (Post) and 12 months (Follow-up) after randomization. After the completion of all assessments, patients of the control group will receive 450 € as reimbursement for their time.

#### Allocation

Six eligible patients are randomized as a group to either the intervention or control condition. The randomization sequence is generated independently of the study coordinator and the research team responsible for data collection and management. The sequence is generated using a varying-size permuted block design, stratified by study site. This procedure ensures an appropriate balance in the number of treatment and control groups per study site. Randomization codes are generated digitally and concealed on a secure system. The group-allocation sequence is concealed from the research team responsible for data collection and management until the assignment. Allocation is performed through RedCap “Randomization Module” on the day of intervention assignment with no prior access for anyone but the data manager.

#### Masking/Blinding

Patients are not blind to allocation. No assessors need to be blinded since primary and secondary outcomes are not based on clinician/assessor ratings. Accelerometer-based data is prepared and processed by trained student assistants according to predefined rules and specifications. Raw data is stored in REDCap and monitored by an external unblinded data manager. The sponsor and his research team responsible for data collection and management and any personal in contact with patients is blinded regarding the randomization procedure. The data analyst of the main analysis regarding efficacy (Hypotheses 1, 3, 4) is blinded regarding the allocation. He receives the final dataset, which is masked for the treatment condition (the “condition” variable only informs Condition A or B but no real labels of the treatment and control conditions). The data analyst will be unblinded after completion of the main analysis regarding efficacy. Analyses regarding health economics (Hypothesis 2) and further analysis regarding implementation/Process evaluation will be conducted after unblinding. An unblinded data manager handles the raw data when exporting the data from REDCap. No one other than the external data manager has access to the longitudinal data before the statistical analysis is completed.

#### Plans to promote participant retention and complete follow-up

In accordance with the Evaluation Concept and Record of Processing Activities, all data for evaluation runs through REDCap and network drives on a secured LMU internal server providing a stable and traceable coding, storage and security structure as well as data quality measures such as automatic survey reminders, alerts to researchers for telephone reminders to participants, and data type and range checks where appropriate. Missing values on surveys have to be confirmed by participants to be intentional before being able to proceed to the next questionnaire.

The process evaluation data collected via app are stored pseudonymously on a server of the Central Office of the University of Tuebingen and transmitted pseudonymously to the central data management site LMU Munich. Likewise, videos of the group sessions will be recorded by the University of Tuebingen as part of the process evaluation. These videos will be stored on a hard-drive at the Central Office of the University of Tuebingen and transferred to central data management site LMU Munich via secured network drive connection. The routine data of the health insurances (AOK/TK) include outpatient, inpatient, as well as medicines, remedies and aids six months prior and up until six months after completion of the intervention period and are transmitted to the trust center of the Technical University Munich on the basis of a pseudonym assigned by the Tue-CO, which is not known to the patients, and from there double pseudonymized to the Technical University Munich. The data stored at the central data management site LMU Munich (primary and secondary endpoints, data on progression diagnostics and process evaluation) are transmitted to Technical University via a secure connection. From there, they are transmitted to the trust center with the routine data and will be double pseudonymized.

#### Data management

##### Data Entry

Participant (patient and therapist) records are created in REDCap by Tue-MA and include pseudonym, e-mail address, study center, metadata and assessment dates.

The data that participants enter in the smartphone application during their participation in the trial are securely stored on their phones. The data are also transmitted to a central server located at University of Tuebingen. Participants are able to share some or all of their data (that is directly related to their participation such as exercise schedules or plans for goal achievement) with their exercise therapist. They can access a summary of the participant’s data via a browser-based application in order to plan their group session or provide feedback and guidance to the individuals.

Randomization by LMU personnel is automatically triggered 2 weeks after Pre-assessment. Participants enter all questionnaire data electronically via REDCap from a date-triggered REDCap invitation link. AE and Dropout-data are entered by staff of the sponsor after notification from the therapist or the participant.

##### Data Quality

Data quality is ensured through several mechanisms: Validity, data and range rules are installed for data entry in REDCap wherever possible. All manual modification and changes to the database will be documented in electronic logs in REDCap and only e-mail addresses will be deleted from logs after data collection. Participants have to confirm it to be an intentional choice if they leave items unanswered on questionnaires. Automatic date-triggered reminders are sent to participants for questionnaires. Tue-MA receive date-triggered alerts for phone-reminders if questionnaires are still missing. Additionally, regular checks by the evaluating center help detect and prevent data discrepancies. The evaluating center will regularly send reports about missing data to the University of Tuebingen.

The trust center ensures the data quality of the routine data used for the economic evaluation by controlling for completeness of the dataset and plausibility of selected variables by calculating means and distributions.

The data transmitted from the smartphone of the participants to the server are stored in a database. New records are appended to the database and do not overwrite previous records. Faulty records can thus be interpolated with the previous and the next one. Established technical measures prevent corruption of data during transmission.

##### Data Security

A data protection concept was developed together with all partners, which was approved by the data protection officers of the University of Tuebingen, the LMU, TU and from the 2 health insurances AOK and TK. The concept can be provided upon request in German language. Violations of data privacy will be documented and reported to an external data protection officer.

The participant’s data are automatically deleted upon deletion of the study smartphone application. Transport to the server is protected by established encryption protocols. Access to the server site is protected via technical and organizational measures of the University of Tuebingen. Access to the data is only granted to specific personell responsible for technical maintenance.

All study personnel are restricted to their user rights to the necessary tasks only, i.e. LMU quality control team cannot access participants’ e-mail address while Tuebingen personnel don’t have access to participants’ questionnaire data. REDCap communication channels are encrypted, and all REDCap Data is stored on a secured internal server with logging and regular backups, which is maintained by the faculty’s IT. REDCap itself is regularly updated.

Original paper versions are safely stored in Tuebingen for 10 years after the completion of the study.

Data to and from the trust center will be transmitted via a secure online connection. Only pseudonymous data is transmitted. For data quality checks the data will be stored and analyzed on an external server without any network connections. Thereafter, data will be locked on an external hard drive within a Veracrypt container and kept within a steel safe. Data is permanently erased one year after the completion of the study.

#### Statistical Methods

To test Hypotheses 1, 2, 3a and 4 we will use multilevel modeling to establish the treatment effects on the primary outcome (global symptom severity) and the secondary outcomes (major depressive disorders, insomnia, panic disorder, agoraphobia, PTSD, QALYs, exercise). In these analyses, each outcome will be predicted by the group (ImPuls + TAU vs. TAU), time (pre vs. post vs. follow-up), and their interaction. Given that the randomization is stratified by study site, we will account for the effects of study site in all analyses. All analyses will be performed on intention-to-treatment basis, using multiple imputations. Restricted maximum likelihood estimation will be used, which is implemented by R (Version 4) lme4 package. Each model will account for the three-level nested structure of the data: the within-person, between-person, and between-site levels. Random intercepts will be assumed to model the between-person and between-site, and random slopes (on time) the between-person variation. The significance level will be set to be alpha = 0·05.

To test Hypothesis 3b, mediation analyses will be performed to test the indirect effects of the treatment on the primary outcome that are mediated by the changes in the putative mediators (self-reported exercise in minutes per week, accelerometry-based moderate to vigorous physical activity (MVPA); from pre to post, pre to follow-up and post to follow-up). We will compute standardized change scores for the outcome and mediators.^135^ In a path model (specified in the framework of structural equation modeling), the group variable predicts the changes in the mediators, and these mediating factors further predict the change score in the outcome. The indirect effects are defined by the products of the group-mediator and mediator-outcome effects. In case of saturated models, we will compare model fits with and without the indirect paths in terms of AIC and BIC. As exploratory analyses, we will also examine other forms of mediations proposed by Goldsmith et al., encompassing cross-lagged effects and latent change score models.^136^ As recommended by Usami, Murayama and Hamaker, we will first test whether these models fit the data well (and which model fits the data best), and then investigate the indirect effects of the treatment on the outcome.^137^

To further analyze clinical significant changes (additional analysis for Hypothesis 4 and 5), the Reliable Change Index (RCI) will be computed for each individual between pre-, post and follow-up assessments.^138-140^ The reliable change index will be calculated each for Post and follow-up GSI scores on the non-imputed dataset. Jacobson-Truax formulation will be used, namely RCI = [GSI (post or follow-up) – GSI (pre)] / S_diff. S_diff was given by sqrt(2)*SD(GSI pre)*sqrt(1 – reliability). The cut-off for a clinically significant change was defined as 12 or smaller. The Mann-Whitney U test will be performed to examine whether the ordinal scores (recovered, improved, unchanged, and deteriorated) are significantly different between the two groups. Sensitivity analyses will be conducted with different analytic approaches. First, analyses on the primary and secondary outcomes will be repeated on the completer sample. Completers are operationalized as those who received at least two complete weeks within the supervised intervention period. Second, complete data analyses will be performed to check if multiple imputations influenced the results.

Concerning process evaluation, we use descriptive statistics to analyze objectives a), b), c), d), e) and f). Moderation analysis is planned to gain more insight concerning objective f) and g) (e. g., with multiple regression analysis) in terms of attendance, dropouts and efficacy. We will also use mediation analysis for objective h) (e.g., structural equation modeling).

Empowerment of exercise therapists (research question 1a) will be analyzed descriptively (mean, standard deviation) with respect to their self-reports on the modified training evaluation scale, the (occupational-) self-efficacy scale and concerning the frequency of supervision.^107,109^

We descriptively present the adherence score (mean, standard deviation) as well as the corresponding percentage to check whether the desired high level of adherence (>90%) in the context of the efficacy trial is achieved (research question 1b).

We descriptively present the amount of all patients acquired by each strategy as well as the amount of all patients included in the study by each strategy. We then check which recruitment strategy has the highest inclusion rate and present the corresponding percentage (research question 1c).

Referring healthcare professional’ opinion on the new treatment option is presented descriptively (mean, standard deviation) (research question 1d). To provide information about the dose delivered (research question 1e), we present documentation data of exercise therapists and the web-based ImPuls interface descriptively.

To assess barriers and facilitators (research question 2a), we use quantitative data from questionnaires (barriers and facilitators assessment instrument, satisfaction scale, as well as basic recommendations for outpatient facilities in Germany.^107,113,118-121^ Results are presented descriptively (mean, standard deviation) to provide an overview over contextual characteristics. This data is further complemented with qualitative data from the interviews.

We evaluate the interviews in a deductive-inductive process following the steps of a content-structuring qualitative content analysis.106 First, two researchers will elaborate a preliminary coding frame for coding based on the interview guideline. Subsequently, 15 % of all interviews are independently coded by those researchers. Inter-coder-reliability analysis is then performed to ensure that the elaborated coding frame is applicable. Potential discrepancies are discussed to refine the coding frame in an iterative process. This process is continued until both researchers agree that the categories are distinct and no new categories need to be added to the coding frame. Afterwards, the remaining interviews are coded. Coding and analysis are done by using the software MAXQDA 2022. In a final step, the statements from all interviews are summarized category by category and used to supplement the quantitative data.

We will conduct mediation and moderation analyses as well as subgroup analyses to gain deeper insight into the impact of the program (mechanisms of change). We check whether exercise therapist’ attitude towards mental disorders (Opening minds scale for health-care providers OMS-HC), evidence-based practice (evidence-based practice attitude scale – German version EBPAS-36D) and the program (measurement of therapy expectation and therapy evaluation of patients (PATHEV) affects treatment effects (research question 3a).^100,111,112^ In addition, patient’ application data as well as changes in respective individual behavioral determinants e.g., action and coping plans; Physical activity-related health competencies (PAHCO) will be used to determine the extent to which core components of the intervention have been used and how this affects treatment effects (research question 3b).^128^ Further analysis is done to determine the extent to which motivational/volitional core components of the ImPuls intervention (application data [barrier management, goal-setting], documentation data [phone contacts]) as well as changes in respective individual behavioral determinants (e.g., action and coping plans; [PAHCO]) affect patient’ exercise adherence (research question 3c). Finally, we want to explore whether psychological processes such as emotional intelligence (Trait emotional intelligence questionnaire (TEIQue), emotional regulation (difficulties in emotion regulation scale (DERS)), repetitive negative thinking (perseverative thinking questionnaire PTQ)) or perceived stress (perceived stress scale (PSS)) mediate the treatment effect on global symptom severity (research question 3d).^124,125,127,129^

Missing data will be handled by multiple imputations.

#### Data Monitoring

##### Composition of Data Safety and Monitoring Board

The Data Safety and Monitoring Board is composed of independent researchers who are not associated with the project but who nevertheless have expertise in clinical or medical research. All members of the Data Safety and Monitoring Board are listed in the table below.

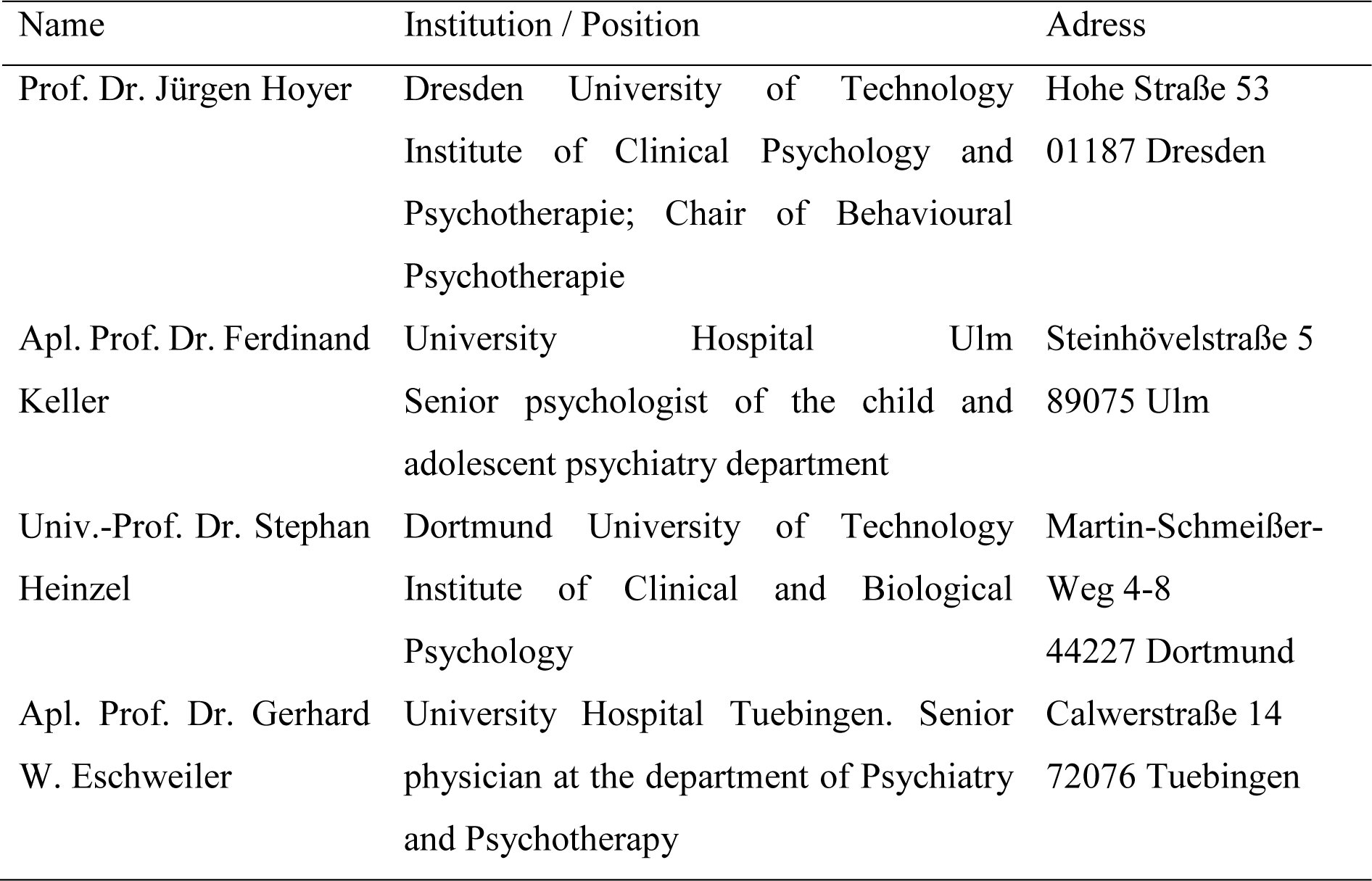

### Interim analyses

There will be no interim analysis.

### (Serious) adverse events

Adjustments to the study or discontinuation of the entire study will be discussed regularly with the independent Data Safety and Monitoring Board in light of reported Adverse Events (AE) or Serious Adverse Events (SAE).

Adverse events (AE) describe any deterioration in the mental or physical condition or behavior of a participant administered the intervention, including events not necessarily caused by or related to the intervention. Following this definition, the following categories are defined as AE for the purposes of this project:

- Onset of new psychological symptoms
- Significant worsening of pre-existing psychological symptoms
- Occurrence of new physical symptoms
- Significant worsening of pre-existing physical symptoms
- Injuries that result in not being able to perform any sports activity for at least 2 months

A serious adverse event (SAE) is any adverse event that results in death, is life-threatening, requires inpatient hospitalization or prolongation of an existing hospital stay, or results in persistent or significant disability/incapacity. Other significant events may also be considered serious if they endanger the participant or require interventions to prevent any of the above mentioned consequences. Consequently, the following events are considered SAE under this project:

- Suicide
- Suicide attempt
- Other event that resulted in death
- Intentional serious self-injury that resulted in an inpatient stay
- Third party injury
- Event that is acutely life-threatening (i.e., study participant is in acute danger of death)
- Event that results in significant physical disability
- Hospitalization due to psychiatric and somatic symptoms

The above-mentioned categories of AE are recorded during the structured assessments (post, follow-up) via online questionnaires. In addition, participants and therapists (in the case of spontaneous reports by patients) have the opportunity to report AEs to a study telephone (available daily from 9 a.m. to 5 p.m.) at the central office at the University of Tuebingen. Employees of the study center document AEs in a CRF.

Based on the above list, SAEs are recorded by the exercise therapists during the intervention phase and reported to employees of the central office of the University of Tuebingen. In addition, SAEs will be collected by employees of the University of Tuebingen at all three measurement time points (pre, post, follow-up) for the intervention and control groups. They then report SAEs to the study director (Dr. Sebastian Wolf), who consults an independent Data and Safety Monitoring Board for further action within 12 hours after the SAE. The Data Safety and Monitoring Board is composed of independent researchers who are not associated with the project but have expertise in clinical or medical research. All members of the Data Safety and Monitoring Board are listed above. The Data Safety and Monitoring Board decides within 24 hours on any actions that may be necessary (e.g., changes in the trial or similar) and passes these on to the study director. All information on AEs and SAEs will also be recorded in the REDCap system, like all other data collected.

The following data is documented for each AE/SAE:

- Pseudonym of participant
- Study site to which the participant was associated
- Date of start of SAE and associated study phase (pre, intervention phase, post or follow-up)
- Description of the SAE
  - information on whether there was a risk for other participants
  - Severity (rated by PI and DSMB)
    - **Mild** – Events require minimal or no treatment and do not interfere with the participant’s daily activities.
    - **Moderate** – Events result in a low level of inconvenience or concern with the therapeutic measures. Moderate events may cause some interference with functioning.
    - **Severe** – Events interrupt a participant’s usual daily activity and may require systemic drug therapy or other treatment. Severe events are usually potentially life-threatening or incapacitating. Of note, the term “severe” does not necessarily equate to “serious”.
  - relationship to study intervention (rated by PI and DSMB)
    - **Related** – The AE is known to occur with the study procedures, there is a reasonable possibility that the study procedures caused the AE, or there is a temporal relationship between the study procedures and the event. Reasonable possibility means that there is evidence to suggest a causal relationship between the study procedures and the AE.
    - **Not Related** – There is not a reasonable possibility that the study procedures caused the event, there is no temporal relationship between the study procedures and event onset, or an alternate etiology has been established.
  - expectedness
    - as rated by the Data Safety and Monitoring Board
  - reporting events to participants
    - yes/no
  - - Date SAE ended
  - - Status at end of SAE (resolved, improved, not improved, consequences/type of harm)

#### Auditing

Data monitoring is ensured through a combination of automatic electronic validation through REDCap and manual checks, performed by the evaluating site (LMU). A complete audit trail including any data entry, access, modification, timepoint of randomization, user rights and exports is maintained by REDCapAfter data collection, pseudomized REDCap data and logs are exported, personal information (E-mail) is cleaned from the logs and logs are stored with the codelist on a secured drive. pseudomized data is stored for the codelist data retention period wiA site initiation procedure for each site was conducted by the LMU before data collection.

#### Ethics

Ethics approval has been obtained (ID: 888/2020B01, 02/11/2020).

#### Informed consent

Interested patients are invited to a first meeting in their respective study center. In this meeting a staff member of the central office from Tuebingen informs patients about the study. All questions are clarified. Afterwards patients sign informed consent. The informed consent files are stored in lockable filing cabinets separated from all other pseudonymized study data. Only selected staff members have access to these cabinets.

#### Confidentiality

In accordance with the Record of processing activities and data protection concept, participants personal information is collected by recruiting staff at Tuebingen, who input pseudonymized records and researcher-collected data into REDCap and maintain a code list. The only exception is email address, which is also input into REDCap for automatic survey invitation and reminder e-mails, but which is not accessible to LMU data analysis personnel. At the same time, survey responses are not accessible to recruiting staff. Email addresses will be deleted from the dataset after data collection is completed, and the code list will be deleted after 10 years. Final published datasets will be anonymized, i.e., published without pseudonym. Personal patient file data (informed consent, protocols of structural clinical interviews) in written form will be stored in-house of the university of Tübingen in securely lockable filing cabinets. Files will be shredded after 10 years. Other personal data (phone lists) will be anonymized after completion of data collection.

Group session videos for process evaluation are stored separately in order to be rated regarding manual adherence. Individual participants are not identified or rated and videos are stored safely at the conducting institution and will be deleted from the network drive after rating.

Personal information of the diagnostics is stored safely and pseudonymized in files at the Tue-CO.

#### Declaration of interest

There are no financial or competing interests to declare.

#### Access to data

The principal investigator (Sebastian Wolf) and the member of the evaluation team (Keisuke Takano, Eva Herzog, Thomas Ehring) will have access to the complete final data. In addition, all other investigators involved in the project will have access to the final data for pre-specified analyses.

#### Ancillary and Post-Trial Care

No provisions and post-trial care are provided.

#### Dissemination policy

A publication committee that consists of representatives of all partners develops a dissemination policy.

The study PI proposes potential papers. PhD students or other researcher propose a topic/paper and write an abstract to the committee. The committee finally decides about acceptance and authorships.

## Supplementary information / Further Results

### Further baseline Characteristics

**Table S1.**
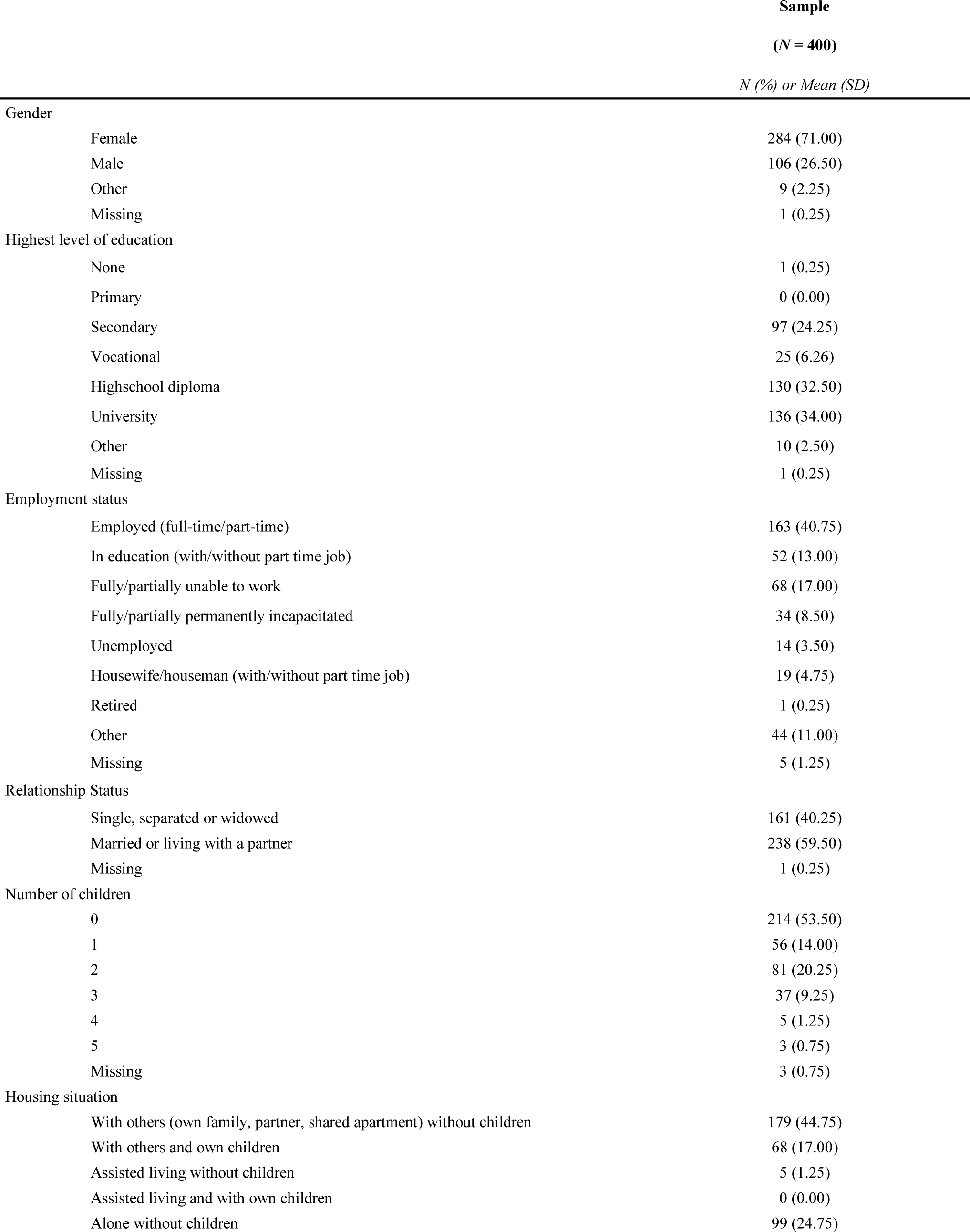
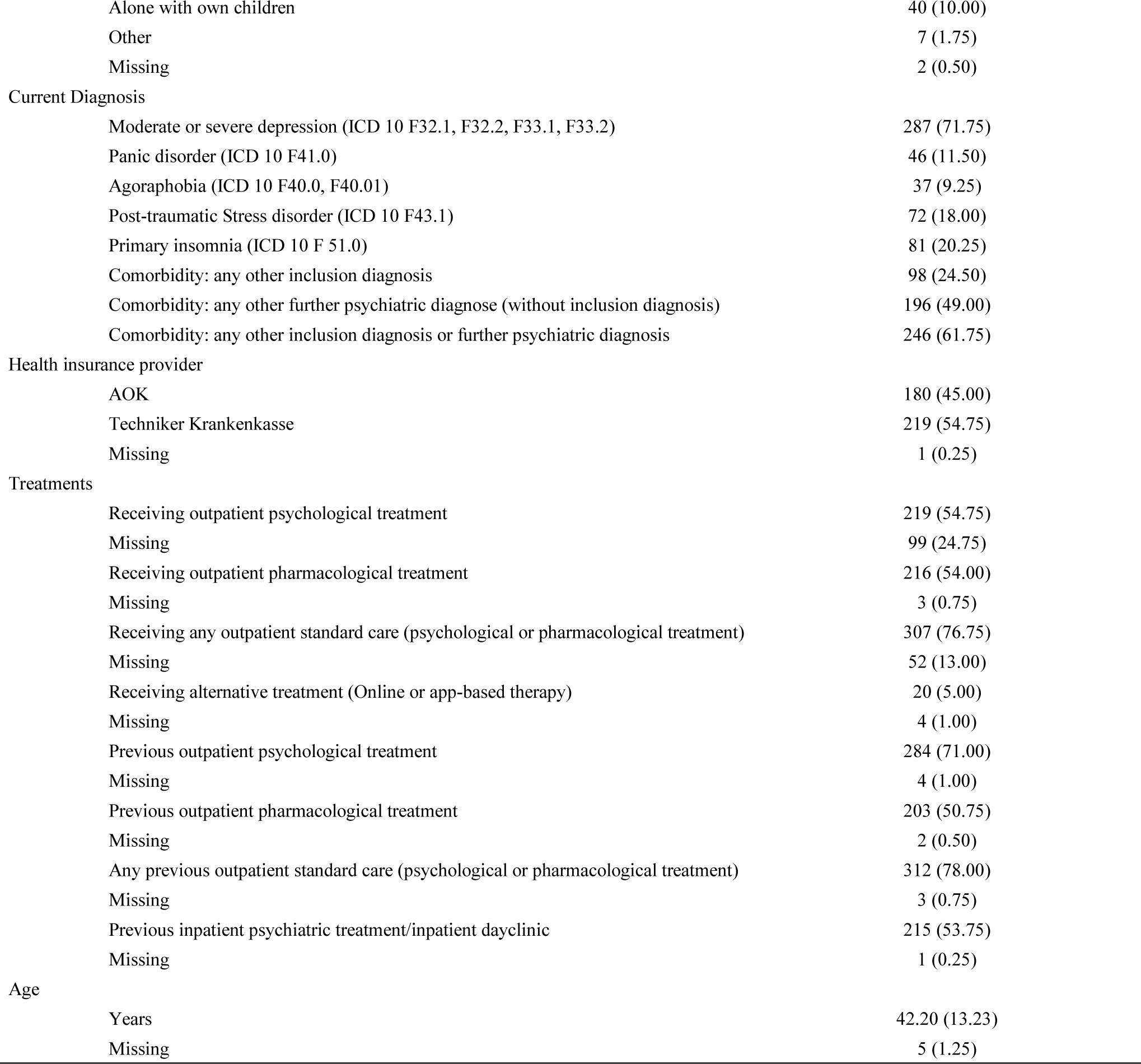
Baseline characteristics of the combined intention-to-treat population (n=400)

|  |  | Sample |
| --- | --- | --- |
|  |  | (N = 400) |
|  |  | N (%) or Mean (SD) |
| Gender |  |  |
|  | Female | 284 (71.00) |
|  | Male | 106 (26.50) |
|  | Other | 9 (2.25) |
|  | Missing | 1 (0.25) |
| Highest level of education |  |  |
|  | None | 1 (0.25) |
|  | Primary | 0 (0.00) |
|  | Secondary | 97 (24.25) |
|  | Vocational | 25 (6.26) |
|  | Highschool diploma | 130 (32.50) |
|  | University | 136 (34.00) |
|  | Other | 10 (2.50) |
|  | Missing | 1 (0.25) |
| Employment status |  |  |
|  | Employed (full-time/part-time) | 163 (40.75) |
|  | In education (with/without part time job) | 52 (13.00) |
|  | Fully/partially unable to work | 68 (17.00) |
|  | Fully/partially permanently incapacitated | 34 (8.50) |
|  | Unemployed | 14 (3.50) |
|  | Housewife/houseman (with/without part time job) | 19 (4.75) |
|  | Retired | 1 (0.25) |
|  | Other | 44 (11.00) |
|  | Missing | 5 (1.25) |
| Relationship Status |  |  |
|  | Single, separated or widowed | 161 (40.25) |
|  | Married or living with a partner | 238 (59.50) |
|  | Missing | 1 (0.25) |
| Number of children |  |  |
|  | 0 | 214 (53.50) |
|  | 1 | 56 (14.00) |
|  | 2 | 81 (20.25) |
|  | 3 | 37 (9.25) |
|  | 4 | 5 (1.25) |
|  | 5 | 3 (0.75) |
|  | Missing | 3 (0.75) |
| Housing situation |  |  |
|  | With others (own family, partner, shared apartment) without children | 179 (44.75) |
|  | With others and own children | 68 (17.00) |
|  | Assisted living without children | 5 (1.25) |
|  | Assisted living and with own children | 0 (0.00) |
|  | Alone without children | 99 (24.75) |

|  |  |
| --- | --- |
| Alone with own children | 40 (10.00) |
| Other | 7 (1.75) |
| Missing | 2 (0.50) |
| Current Diagnosis |  |
| Moderate or severe depression (ICD 10 F32.1, F32.2, F33.1, F33.2) | 287 (71.75) |
| Panic disorder (ICD 10 F41.0) | 46 (11.50) |
| Agoraphobia (ICD 10 F40.0, F40.01) | 37 (9.25) |
| Post-traumatic Stress disorder (ICD 10 F43.1) | 72 (18.00) |
| Primary insomnia (ICD 10 F 51.0) | 81 (20.25) |
| Comorbidity: any other inclusion diagnosis | 98 (24.50) |
| Comorbidity: any other further psychiatric diagnose (without inclusion diagnosis) | 196 (49.00) |
| Comorbidity: any other inclusion diagnosis or further psychiatric diagnosis | 246 (61.75) |
| Health insurance provider |  |
| AOK | 180 (45.00) |
| Techniker Krankenkasse | 219 (54.75) |
| Missing | 1 (0.25) |
| Treatments |  |
| Receiving outpatient psychological treatment | 219 (54.75) |
| Missing | 99 (24.75) |
| Receiving outpatient pharmacological treatment | 216 (54.00) |
| Missing | 3 (0.75) |
| Receiving any outpatient standard care (psychological or pharmacological treatment) | 307 (76.75) |
| Missing | 52 (13.00) |
| Receiving alternative treatment (Online or app-based therapy) | 20 (5.00) |
| Missing | 4 (1.00) |
| Previous outpatient psychological treatment | 284 (71.00) |
| Missing | 4 (1.00) |
| Previous outpatient pharmacological treatment | 203 (50.75) |
| Missing | 2 (0.50) |
| Any previous outpatient standard care (psychological or pharmacological treatment) | 312 (78.00) |
| Missing | 3 (0.75) |
| Previous inpatient psychiatric treatment/inpatient dayclinic | 215 (53.75) |
| Missing | 1 (0.25) |
| Age |  |
| Years | 42.20 (13.23) |
| Missing | 5 (1.25) |

**Table S2.** Further patient characteristic - Comorbidity assessments.

|  | Intervention group |  | Control group |  |
| --- | --- | --- | --- | --- |
|  | (N = 199) |  | (N = 201) |  |
|  | N | % | N | % |
| Comorbid Diagnosis |  |  |  |  |
| F10.1, F10.10 - Mental and behavioural disorders due to harmful use of alcohol | 6 | 3.02 | 5 | 2.49 |
| F12.1, F12.10 - Mental and behavioural disorders due to harmful use of cannabinoids | 3 | 1.51 | 2 | 1.00 |
| F32.0, F32.4 - Mild or partially remitted depressive episode | 5 | 2.51 | 6 | 2.99 |
| F32.81 - Premenstrual dysphoric disorder | 3 | 1.51 | 6 | 2.99 |
| F33.0, F33.41 - Mild or partially remitted recurrent depressive disorder | 10 | 5.03 | 11 | 5.47 |
| F34.1 - Dysthymia | 38 | 19.1 | 44 | 21.89 |
| F40.1, F40.10 - Social phobia | 12 | 6.03 | 22 | 10.95 |
| F40.2 - Specific (isolated) phobias | 15 | 7.54 | 14 | 6.97 |
| F41.1 - Generalized anxiety disorder | 6 | 3.02 | 8 | 3.98 |
| F42.0, F42.1, F42.2, F.42.3, F.42.4 - Obsessive-compulsive disorder | 10 | 5.03 | 13 | 6.47 |
| F43.1 - Posttraumatic Stress disorder, partially remitted | 1 | 0.50 | 0 |  |
| F45.0, F45.1, F45.2, F45.41 - Somatoform disorders | 9 | 4.52 | 6 | 2.99 |
| F90.0, F90.1, 90.2 - Hyperkinetic disorders | 6 | 3.02 | 5 | 2.49 |

**Table S3.** Characteristics of study sites and Allocation information.

| Study site (city) | Type of facility | Indications in outpatient care | Number of Study therapists | Allocations (68) |  |
| --- | --- | --- | --- | --- | --- |
|  |  |  |  | total | to treatment<br>N (%) |
| Bietigheim | Outpatient medical rehabilitation facility | 2 (Orthopedics, Traumatology);<br>6 (Neurology, Neuropsychology) | 1 | 6 | 3 (50) |
| Crailsheim | Outpatient medical rehabilitation facility | 2 (Orthopedics, Traumatology) | 0 | 1 | 0 (0) |
| Göppingen | Outpatient medical rehabilitation facility | 2 (Orthopedics, Traumatology);<br>5 (Gastroenterology) | 3 | 6 | 3 (50) |
| Heidelberg | Outpatient center for physiotherapy & exercise therapy | 2 (Orthopedics, Traumatology);<br>6 (Neurology, Neuropsychology);<br>9 (Geriatrics) | 2 | 10 | 6 (60) |
| Karlsruhe | Outpatient medical rehabilitation facility | 2 (Orthopedics, Traumatology);<br>3 (Rheumatology);<br>6 (Neurology, Neuropsychology) | 3 | 10 | 5 (50) |
| Tübingen | Outpatient center for physiotherapy & exercise therapy | 2 (Orthopedics, Traumatology);<br>6 (Neurology, Neuropsychology);<br>11 (Other: „Gynecology/Urology“) | 2 | 10 | 5 (50) |
| Ulm | Outpatient medical rehabilitation facility | 2 (Orthopedics, Traumatology); | 2 | 5 | 3 (60) |
| Weingarten | Outpatient center for physiotherapy & exercise therapy | 1 (Cardiology, Angiology, Hematology);<br>2 (Orthopedics, Traumatology);<br>6 (Neurology, Neuropsychology) | 3 | 6 | 3 (50) |
| Stuttgart | Outpatient medical rehabilitation facility | 2 (Orthopedics, Traumatology);<br>3 (Rheumatology) | 2 | 8 | 4 (50) |
| Freiburg | Outpatient medical rehabilitation facility | 10 (Ppsychosomatics/psychiatry) | 2 | 6 | 2 (33,3) |

**Table S4.** Characteristics of Exercise therapists at baseline (*n* = 20)

|  |  | *n* | *%* |  |  |  |
| --- | --- | --- | --- | --- | --- | --- |
| Gender | Female | 11 | 0.55 |  |  |  |
|  | Male | 9 | 0.45 |  |  |  |
|  | other | 0 | 0.00 |  |  |  |
| Age | 20-29 | 10 | 0.50 |  |  |  |
|  | 30-39 | 3 | 0.15 |  |  |  |
|  | 40-49 | 5 | 0.25 |  |  |  |
|  | 50-60 | 2 | 0.10 |  |  |  |
| Highest Education |  |  |  |  |  |  |
|  | Academic degree in exercise or movement science (e.g. Magister, Bachelor, Master) | 13 | 0.65 |  |  |  |
|  | Non-academic technical college degree | 2 | 0.10 |  |  |  |
|  | Exercise and Caring/Therapeutic Gymnastics |  |  |  |  |  |
|  | Academic or non-academic degrees in Physiotherapy<sup>1</sup> | 5 | 0.25 |  |  |  |
|  |  | *n* | *Mean* | *SD* | *Median* | *IQR* |
| Age |  |  |  |  |  |  |
|  | Years | 20 | 33.95 | 11.35 | 29.50 | 25.50 – 42.25 |
| Assessments |  |  |  |  |  |  |
|  | Working experience with patients with mental disorders (scale 0-10, “On a scale of 0-10, how much experience do you have with sport therapy for people with psychological disorders?”) | 20 | 3.59 | 3.15 | 2.55 | 1.15 – 7.15 |
|  | Working experience (years) | 20 | 8.80 | 9.81 | 4.75 | 1.50 – 12.50 |
|  | Treatment motivation at pre-assessment (SESSW<sup>2</sup>; range 1 (“Does not apply at all”) – 5 (“Strongly applies”)) | 19 | 4.25 | 0.57 | 4.22 | 4.00 – 4.72 |
|  | Satisfaction with Intervention at pre-assessment (B&F-A<sup>2</sup>, (range 1 (“Strongly disagree”) – 5 (“Strongly agree”, 6 = “Not applicable”)) | 15 | 3.39 | 0.91 | 3.67 | 3.17 – 3.92 |
|  | Expectations of intervention success at pre-assessment (PATHEV, subscale “Hope of improvement”<sup>2</sup>, range 1 (“Does not apply at all”) – 5 (“Strongly applies”)) | 19 | 3.88 | 0.55 | 4.00 | 3.50 – 4.00 |

*Note.* <sup>1</sup>*n* = 4 of the physiotherapists have completed an additional training as sports teacher or sports and gymnastic teacher. <sup>2</sup>Higher values represent greater manifestation (i.e., motivation, satisfaction, expectations).

**Table S5.** Recruitment Success by referrer.

| Categories | N | Patients randomized<br>(N=400) |
| --- | --- | --- |
|  |  | % |
| Outpatient psychotherapist practice | 135 | 33,75% |
| Telephone prospecting of health insurance company (TK) | 60 | 15,00% |
| Reports in newspapers | 32 | 8,00% |
| Report in internal magazine of health insurance company (AOK) | 29 | 7,25% |
| Outpatient general practitioner practice | 26 | 6,50% |
| University mailing lists | 23 | 5,75% |
| Friends & acquaintances | 21 | 5,25% |
| Social media posts (Instagram, Facebook) | 18 | 4,50% |
| Flyer/poster/roll-ups (without recommendation) | 16 | 4,00% |
| Not specified | 8 | 2,00% |
| TV news report | 7 | 1,75% |
| Clinic/psychiatry/rehabilitation clinic | 6 | 1,50% |
| Outpatient physiotherapy practice | 5 | 1,25% |
| Telephone prospecting of health insurance company (AOK) | 5 | 1,25% |
| Outpatient psychiatrist practice | 4 | 1,00% |
| Other (e.g. radio, socio-psychiatric service, further specialist doctors) | 4 | 1,00% |
| Self-help groups for mental health | 1 | 0,25% |

### Internal validity assessments/Process Evaluation

**Table S6.**
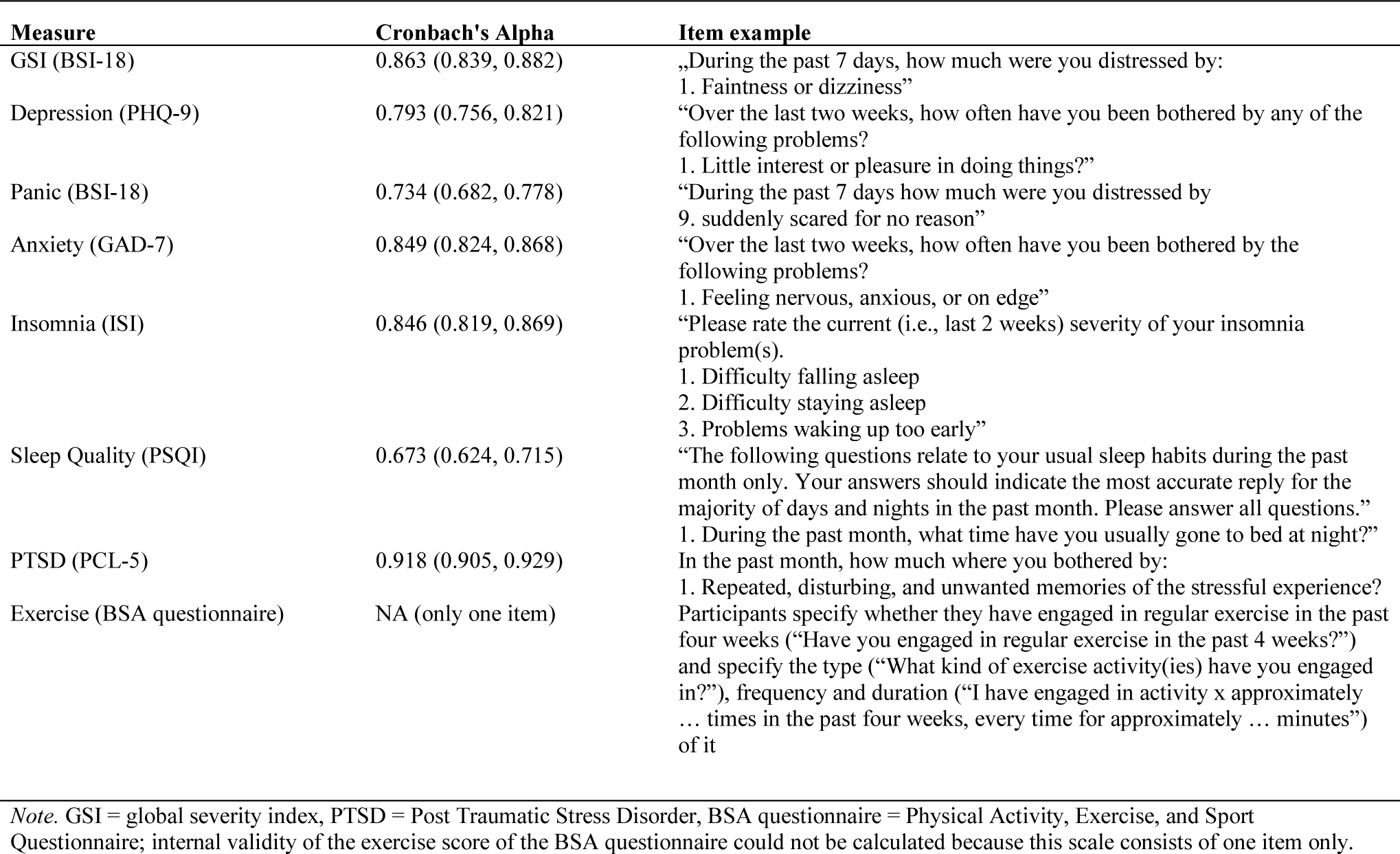
Cronbach’s Alpha at baseline of all primary and secondary assessments.

| Measure | Cronbach's Alpha | Item example |
| --- | --- | --- |
| GSI (BSI-18) | 0.863 (0.839, 0.882) | „During the past 7 days, how much were you distressed by:<br>1. Faintness or dizziness” |
| Depression (PHQ-9) | 0.793 (0.756, 0.821) | “Over the last two weeks, how often have you been bothered by any of the following problems?<br>1. Little interest or pleasure in doing things?” |
| Panic (BSI-18) | 0.734 (0.682, 0.778) | “During the past 7 days how much were you distressed by<br>9. suddenly scared for no reason” |
| Anxiety (GAD-7) | 0.849 (0.824, 0.868) | “Over the last two weeks, how often have you been bothered by the following problems?<br>1. Feeling nervous, anxious, or on edge” |
| Insomnia (ISI) | 0.846 (0.819, 0.869) | “Please rate the current (i.e., last 2 weeks) severity of your insomnia problem(s).<br>1. Difficulty falling asleep<br>2. Difficulty staying asleep<br>3. Problems waking up too early” |
| Sleep Quality (PSQI) | 0.673 (0.624, 0.715) | “The following questions relate to your usual sleep habits during the past month only. Your answers should indicate the most accurate reply for the majority of days and nights in the past month. Please answer all questions.”<br>1. During the past month, what time have you usually gone to bed at night?” |
| PTSD (PCL-5) | 0.918 (0.905, 0.929) | In the past month, how much were you bothered by:<br>1. Repeated, disturbing, and unwanted memories of the stressful experience? |
| Exercise (BSA questionnaire) | NA (only one item) | Participants specify whether they have engaged in regular exercise in the past four weeks (“Have you engaged in regular exercise in the past 4 weeks?”) and specify the type (“What kind of exercise activity(ies) have you engaged in?”), frequency and duration (“I have engaged in activity x approximately ... times in the past four weeks, every time for approximately ... minutes”) of it |

*Note.* GSI = global severity index, PTSD = Post Traumatic Stress Disorder, BSA questionnaire = Physical Activity, Exercise, and Sport Questionnaire; internal validity of the exercise score of the BSA questionnaire could not be calculated because this scale consists of one item only.

## Treatment fidelity

12 of the previously randomly selected videos had to be replaced due to one of the following reasons 1) only one participant attended the session (i.e., not fulfilling the criteria of a group-based intervention), 2) inadequate quality of the video, such as no sound, that made it impossible to rate the video, 3) parts of the session were not recorded due to technical glitches, or 4) the video was recorded, but accidentally not saved and was therefore no longer available. Another session of the respective group was randomly chosen, and if that was not possible due to one of the previously mentioned reasons, the same session for another group was randomly selected. Replacements were conducted using Excel (Microsoft Excel Version 16.75).

We estimated inter-rater reliability of the two assessors by Cohen’s kappa statistics and tested the 0-hypothesis that the extent of agreement is the same as random (kappa=0). With a Cohen’s Kappa of 0.507 (moderate agreement), our analysis revealed a significant agreement between the two raters or observers (Z = 6.8527, 95% CI 0.398 to 0.616; p< .001).

**Table S7.**
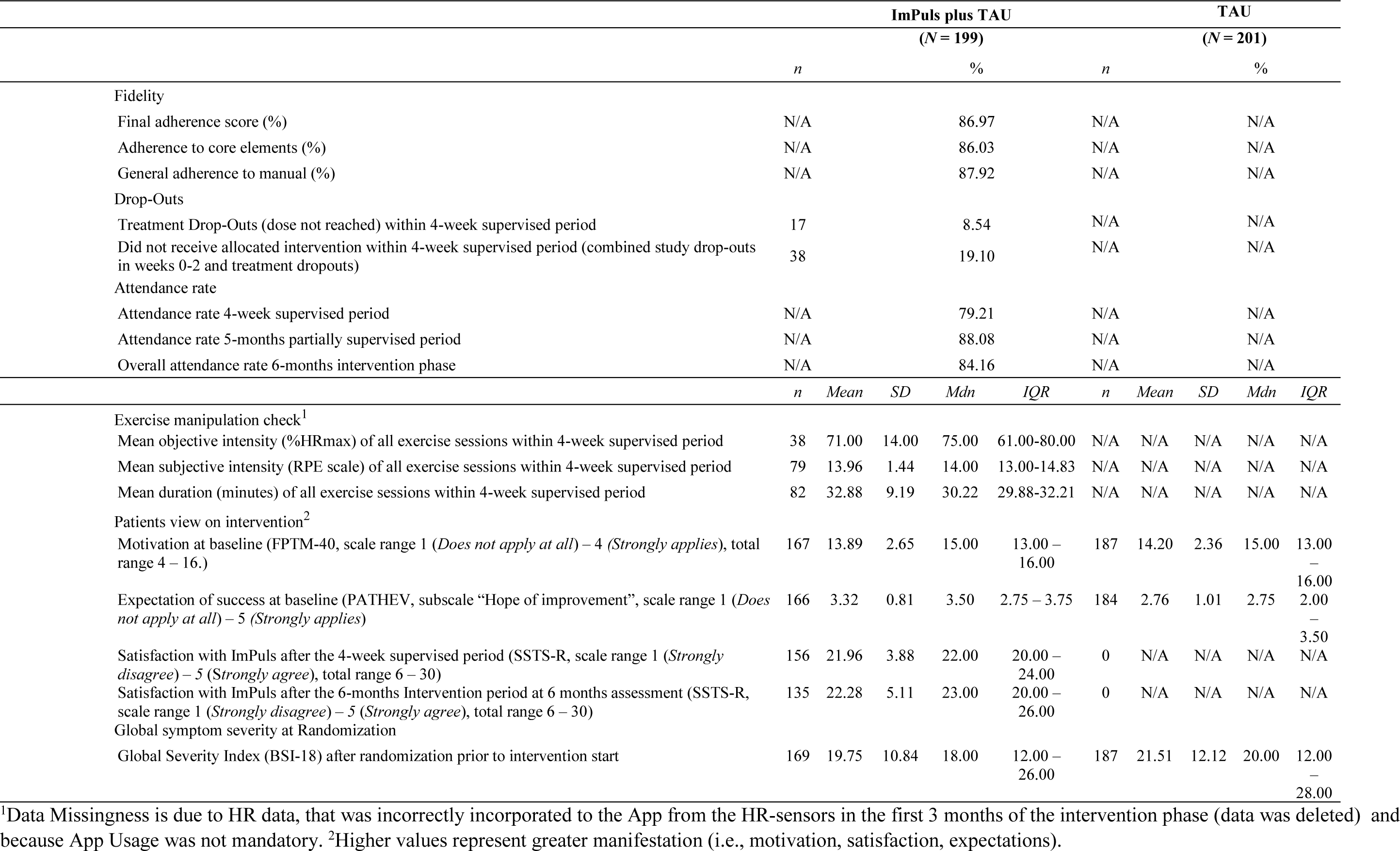
Quality assessments of Process evaluation for internal validity.

## Results of the Reliable clinical Change Analysis

**Table S8.** Reliable clinical change Index at 6- and 12-mothts assessment based on the Jacobson-Truax formulation (Complete Case Analysis)

| Group / Time | Recovered<br>N(%) | Improved<br>N(%) | Unchanged<br>N(%) | Deteriorated<br>N(%) | M-W test |
| --- | --- | --- | --- | --- | --- |
| TAU / 6<br>months | 28 (15) | 28 (13) | 118 (62) | 20 (11) | W = 11551, **p = .002\*** |
| ImPuls plus<br>TAU / 6<br>months | 42 (28) | 22 (15) | 73 (49) | 11 (7) |  |
| TAU / 12<br>months | 38 (20) | 26 (13) | 107 (55) | 23 (12) | W = 11084, **p = .009\*** |
| ImPuls plus<br>TAU / 12<br>months | 45 (33) | 14 (10) | 67 (50) | 9 (7) |  |

\*Significant result: $p < 0.05$

## Further results of the mixed models (ITT sample)

**Table S9.** Results of mixed model with global symptom severity (BSI-18) as the outcome, based on 10 multiply imputed datasets on the ITT sample.

| Predictor | Estimate | 95% CI | SE | t | p |
| --- | --- | --- | --- | --- | --- |
| Intercept | 22.02 | [20.40, 23.63] | 0.83 | 26.68 | **<0.001\*** |
| Timepoint (*ref. baseline*) |  |  |  |  |  |
| 6-months | -2.45 | [-3.94, -0.96] | 0.76 | -3.22 | **0.001\*** |
| 12-months | -3.65 | [-5.14, -2.15] | 0.76 | -4.79 | **<0.001\*** |
| Group (*ref. TAU*) |  |  |  |  |  |
| ImPuls plus TAU | 0.06 | [-2.23, 2.36] | 1.17 | 0.05 | 0.958 |
| Interaction (*ref. Baseline, TAU*) |  |  |  |  |  |
| 6-months, ImPuls plus TAU | -4.17 | [-6.32, -2.02] | 1.10 | -3.80 | **<0.001\*** |
| 12-months, ImPuls plus TAU | -3.35 | [-5.57, -1.13] | 1.13 | -2.96 | **0.003\*** |

Note. CI: confidence interval, ref.: reference category, SE: standard error. \*Significant result: $p < 0.05$

**Table S10.** Results of mixed model with Depression (PHQ-9) as the outcome, based on 10 multiply imputed datasets on the ITT sample.

| Predictor | Estimate | 95% CI | SE | t | p |
| --- | --- | --- | --- | --- | --- |
| Intercept | 13.75 | [12.98, 14.52] | 0.39 | 35.02 | **<0.001\*** |
| Timepoint (*ref. baseline*) |  |  |  |  |  |
| 6-months | -1.61 | [-2.36, -0.86] | 0.38 | -4.19 | **<0.001\*** |
| 12-months | -2.03 | [-2.80, -1.26] | 0.39 | -5.17 | **<0.001\*** |
| Group (*ref. TAU*) |  |  |  |  |  |
| ImPuls plus TAU | 0.01 | [-1.08, 1.09] | 0.55 | 0.01 | 0.988 |
| Interaction (*ref. Baseline, TAU*) |  |  |  |  |  |
| 6-months, ImPuls plus TAU | -1.27 | [-2.37, -0.17] | 0.56 | -2.25 | **0.024\*** |
| 12-months, ImPuls plus TAU | -1.48 | [-2.67, -0.29] | 0.61 | -2.43 | **0.016\*** |

Note. CI: confidence interval, ref.: reference category, SE: standard error. \*Significant result: $p < 0.05$

**Table S11.** Results of mixed model with Insomnia Symptoms (ISI) as the outcome, based on 10 multiply imputed datasets on the ITT sample.

| Predictor | Estimate | 95% CI | SE | t | p |
| --- | --- | --- | --- | --- | --- |
| Intercept | 14.55 | [13.51, 15.58] | 0.53 | 27.58 | <0.001* |
| Timepoint (*ref. baseline*) |  |  |  |  |  |
| 6-months | -1.28 | [-2.12, -0.44] | 0.43 | -2.98 | **0.003\*** |
| 12-months | -2.04 | [-2.90, -1.18] | 0.44 | -4.64 | <0.001* |
| Group (*ref. TAU*) |  |  |  |  |  |
| ImPuls plus TAU | 0.52 | [-0.68, 1.71] | 0.61 | 0.85 | 0.396 |
| Interaction (*ref. Baseline, TAU*) |  |  |  |  |  |
| 6-months, ImPuls plus TAU | -1.76 | [-3.02, -0.50] | 0.64 | -2.74 | **0.006\*** |
| 12-months, ImPuls plus TAU | -1.47 | [-2.74, -0.20] | 0.65 | -2.27 | **0.023\*** |

Note. CI: confidence interval, ref.: reference category, SE: standard error. \*Significant result: $p < 0.05$

**Table S12.** Results of mixed model with Sleep Quality (PSQI) as the outcome, based on 10 multiply imputed datasets on the ITT sample.

| Predictor | Estimate | 95% CI | SE | t | p |
| --- | --- | --- | --- | --- | --- |
| Intercept | 9.90 | [9.33, 10.47] | 0.29 | 34.29 | <0.001* |
| Timepoint (*ref. baseline*) |  |  |  |  |  |
| 6-months | -0.67 | [-1.20, -0.14] | 0.27 | -2.48 | **0.014\*** |
| 12-months | -0.74 | [-1.25, -0.24] | 0.26 | -2.88 | **0.004\*** |
| Group (*ref. TAU*) |  |  |  |  |  |
| ImPuls plus TAU | 0.17 | [-0.60, 0.94] | 0.39 | 0.44 | 0.662 |
| Interaction (*ref. Baseline, TAU*) |  |  |  |  |  |
| 6-months, ImPuls plus TAU | -0.59 | [-1.38, 0.20] | 0.40 | -1.46 | 0.146 |
| 12-months, ImPuls plus TAU | -0.61 | [-1.40, 0.18] | 0.40 | -1.51 | 0.134 |

Note. CI: confidence interval, ref.: reference category, SE: standard error. \*Significant result: $p < 0.05$

**Table S13.** Results of mixed model with general Anxiety (GAD-7) as the outcome, based on 10 multiply imputed datasets on the ITT sample.

| Predictor | Estimate | 95% CI | SE | t | p |
| --- | --- | --- | --- | --- | --- |
| Intercept | 10.66 | [9.97, 11.36] | 0.35 | 30.25 | <0.001* |
| Timepoint (*ref. baseline*) |  |  |  |  |  |
| 6-months | -1.24 | [-1.90, -0.59] | 0.33 | -3.73 | <0.001* |
| 12-months | -1.73 | [-2.38, -1.08] | 0.33 | -5.20 | <0.001* |
| Group (*ref. TAU*) |  |  |  |  |  |
| ImPuls plus TAU | -0.12 | [-1.09, 0.84] | 0.49 | -0.25 | 0.801 |
| Interaction (*ref. Baseline, TAU*) |  |  |  |  |  |
| 6-months, ImPuls plus TAU | -1.16 | [-2.11, -0.21] | 0.49 | -2.39 | **0.017\*** |
| 12-months, ImPuls plus TAU | -1.10 | [-2.12, -0.07] | 0.52 | -2.10 | **0.037\*** |

Note. CI: confidence interval, ref.: reference category, SE: standard error. \*Significant result: $p < 0.05$

**Table S14.** Results of mixed model with Panic (BSI-18) as the outcome, based on 10 multiply imputed datasets on the ITT sample.

| Predictor | Estimate | 95% CI | SE | t | p |
| --- | --- | --- | --- | --- | --- |
| Intercept | 2.59 | [2.25, 2.92] | 0.17 | 15.12 | <0.001* |
| Timepoint (*ref. baseline*) |  |  |  |  |  |
| 6-months | -0.40 | [-0.73, -0.06] | 0.17 | -2.32 | **0.020\*** |
| 12-months | -0.46 | [-0.80, -0.13] | 0.17 | -2.71 | **0.007\*** |
| Group (*ref. TAU*) |  |  |  |  |  |
| ImPuls plus TAU | 0.21 | [-0.26, 0.69] | 0.24 | 0.88 | 0.376 |
| Interaction (*ref. Baseline, TAU*) |  |  |  |  |  |
| 6-months, ImPuls plus TAU | -0.81 | [-1.31, -0.31] | 0.25 | -3.20 | **0.001\*** |
| 12-months, ImPuls plus TAU | -0.79 | [-1.28, -0.29] | 0.25 | -3.12 | **0.002\*** |

Note. CI: confidence interval, ref.: reference category, SE: standard error. \*Significant result: $p < 0.05$

**Table S15.**
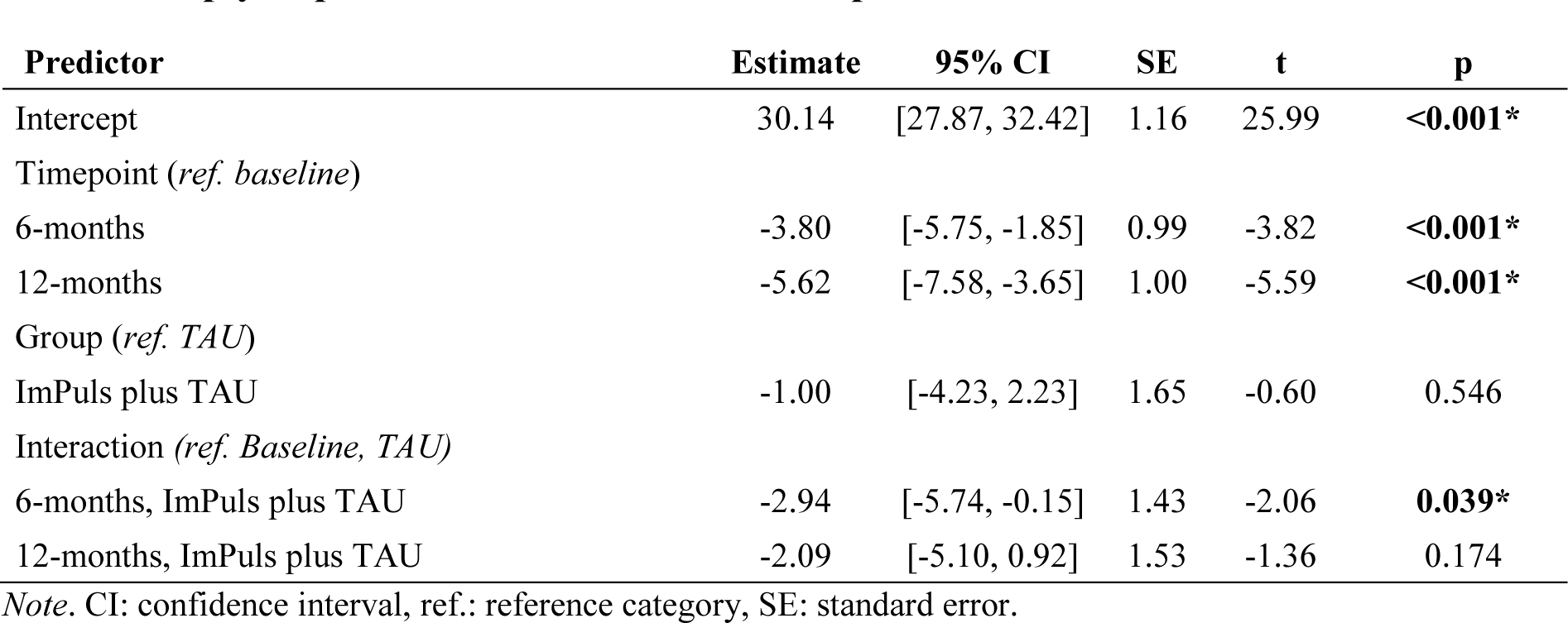
Results of mixed model with PTSD symptoms (PCL-5) as the outcome, based on 10 multiply imputed datasets on the ITT sample.

| Predictor | Estimate | 95% CI | SE | t | p |
| --- | --- | --- | --- | --- | --- |
| Intercept | 30.14 | [27.87, 32.42] | 1.16 | 25.99 | <0.001* |
| Timepoint (*ref. baseline*) |  |  |  |  |  |
| 6-months | -3.80 | [-5.75, -1.85] | 0.99 | -3.82 | <0.001* |
| 12-months | -5.62 | [-7.58, -3.65] | 1.00 | -5.59 | <0.001* |
| Group (*ref. TAU*) |  |  |  |  |  |
| ImPuls plus TAU | -1.00 | [-4.23, 2.23] | 1.65 | -0.60 | 0.546 |
| Interaction (*ref. Baseline, TAU*) |  |  |  |  |  |
| 6-months, ImPuls plus TAU | -2.94 | [-5.74, -0.15] | 1.43 | -2.06 | **0.039\*** |
| 12-months, ImPuls plus TAU | -2.09 | [-5.10, 0.92] | 1.53 | -1.36 | 0.174 |

Note. CI: confidence interval, ref.: reference category, SE: standard error.

**Table S16.** Results of mixed model with self-reported exercise as the outcome, based on 10 multiply imputed datasets on the ITT sample.

| Predictor | Estimate | 95% CI | SE | t | p |
| --- | --- | --- | --- | --- | --- |
| Intercept | 0.71 | [0.47, 0.95] | 0.12 | 5.77 | <0.001* |
| Timepoint (*ref. baseline*) |  |  |  |  |  |
| 6-months | 0.67 | [0.32, 1.03] | 0.18 | 3.74 | <0.001* |
| 12-months | 0.81 | [0.46, 1.16] | 0.18 | 4.55 | <0.001* |
| Group (*ref. TAU*) |  |  |  |  |  |
| ImPuls plus TAU | 0.02 | [-0.31, 0.36] | 0.17 | 0.13 | 0.898 |
| Interaction (*ref. Baseline, TAU*) |  |  |  |  |  |
| 6-months, ImPuls plus TAU | 1.09 | [0.59, 1.58] | 0.25 | 4.28 | <0.001* |
| 12-months, ImPuls plus TAU | 0.42 | [-0.16, 1.00] | 0.30 | 1.41 | 0.161 |

Note. CI: confidence interval, ref.: reference category, SE: standard error. \*Significant result: $p < 0.05$

**Table S17.** Results of mixed model with accelerometry based moderate to vigorous physical activity (MVPA) as the outcome, based on 10 multiply imputed datasets on the ITT sample.

| Predictor | Estimate | 95% CI | SE | t | p |
| --- | --- | --- | --- | --- | --- |
| Intercept | 5.56 | [5.44, 5.69] | 0.06 | 92.33 | <0.001 |
| Timepoint (*ref. baseline*) |  |  |  |  |  |
| 6-months | -0.02 | [-0.13, 0.09] | 0.06 | -0.31 | 0.754 |
| 12-months | 0.04 | [-0.08, 0.16] | 0.06 | 0.67 | 0.504 |
| Group (*ref. TAU*) |  |  |  |  |  |
| ImPuls plus TAU | -0.02 | [-0.18, 0.15] | 0.08 | -0.21 | 0.832 |
| Interaction (*ref. Baseline, TAU*) |  |  |  |  |  |
| 6-months, ImPuls plus TAU | 0.04 | [-0.13, 0.21] | 0.09 | 0.47 | 0.642 |
| 12-months, ImPuls plus TAU | -0.01 | [-0.17, 0.16] | 0.08 | -0.11 | 0.914 |

Note. CI: confidence interval, ref.: reference category, SE: standard error. \*Significant result: $p < 0.05$

## Mediation Analysis (ITT sample)

**Table S18.** Results of structural equation modeling with bootstrapping (5000 iterations) on the ITT sample for model on changes of global symptom severity from baseline to 6 months assessment. Missings were handled with full-information maximum likelihood estimation. The model was saturated and absolute fit indices were not available

| Causal Relationship<sup>a</sup> |  |  | *Est* | *SE* | *z* | *p* |
| --- | --- | --- | --- | --- | --- | --- |
| *direct effects* |  |  |  |  |  |  |
| Δ GSI | ← | Condition (ImPuls plus TAU vs. TAU) | -0.18 | 0.05 | -3.56 | **<0.001\*** |
| Δ GSI | ← | Δ MVPA | -0.02 | 0.07 | -0.27 | 0.788 |
| Δ GSI | ← | Δ self-reported Exercise | -0.11 | 0.06 | -1.84 | 0.067 |
| Δ MVPA | ← | Condition (ImPuls plus TAU vs. TAU) | 0.02 | 0.06 | 0.36 | 0.718 |
| Δ self-reported exercise | ← | Condition (ImPuls plus TAU vs. TAU) | 0.30 | 0.05 | 5.66 | **<0.001\*** |
| Δ MVPA | ↔ | Δ self-reported Exercise | 0.14 | 0.07 | 1.94 | 0.053 |
| *indirect effects* |  |  |  |  |  |  |
| Δ GSI | ← | Condition x Δ MVPA | 0 | 0 | -0.21 | 0.836 |
| Δ GSI | ← | Condition x Δ self-reported Exercise | -0.03 | 0.02 | -1.77 | 0.077 |
| *total effect* |  |  | -0.21 | 0.05 | -4.07 | **<0.001\*** |

Note. <sup>a</sup> = MVPA and self-reported exercise were log-transformed, Est = standardized regression coefficient, SE = standard error of the mean, Δ GSI = change in Global Symptom severity from baseline to 6-months, Δ MVPA= change in accelerometry based physical activity, TAU = treatment as usual, \*Significant result: $p < 0.05$

**Table S19.** Results of structural equation modeling with bootstrapping (5000 iterations) on the ITT sample for model on changes of global symptom severity from baseline to 12 months assessment. Missings were handled with full-information maximum likelihood estimation. The model was saturated and absolute fit indices were not available

| Causal Relationship<sup>a</sup> |  |  | Est | SE | z | p |
| --- | --- | --- | --- | --- | --- | --- |
| *direct effects* |  |  |  |  |  |  |
| Δ GSI | ← | Condition (ImPuls plus TAU vs. TAU) | -0.16 | 0.05 | -2.91 | **0.004\*** |
| Δ GSI | ← | Δ MVPA | 0.04 | 0.06 | 0.70 | 0.482 |
| Δ GSI | ← | Δ self-reported Exercise | -0.19 | 0.05 | -4.02 | **<0.001\*** |
| Δ MVPA | ← | Condition (ImPuls plus TAU vs. TAU) | 0.02 | 0.07 | 0.37 | 0.709 |
| Δ self-reported exercise | ← | Condition (ImPuls plus TAU vs. TAU) | 0.12 | 0.06 | 2.11 | **0.035\*** |
| Δ MVPA | ↔ | Δ self-reported Exercise | 0.08 | 0.07 | 1.15 | 0.252 |
| *indirect effects* |  |  |  |  |  |  |
| Δ GSI | ← | Condition x Δ MVPA | 0.00 | 0.00 | 0.34 | 0.735 |
| Δ GSI | ← | Condition x Δ self-reported Exercise | -0.02 | 0.01 | -1.89 | 0.059 |
| *total effect* |  |  | -0.18 | 0.05 | -3.28 | **0.001\*** |

Note. <sup>a</sup> = MVPA and self-reported exercise were log-transformed, Est = standardized regression coefficient, SE = standard error of the mean, Δ GSI = change in Global Symptom severity from baseline to 6-months, Δ MVPA= change in accelerometry based physical activity, TAU = treatment as usual, \*Significant result: p < 0.05

## Attrition Analysis

**Table S20.**
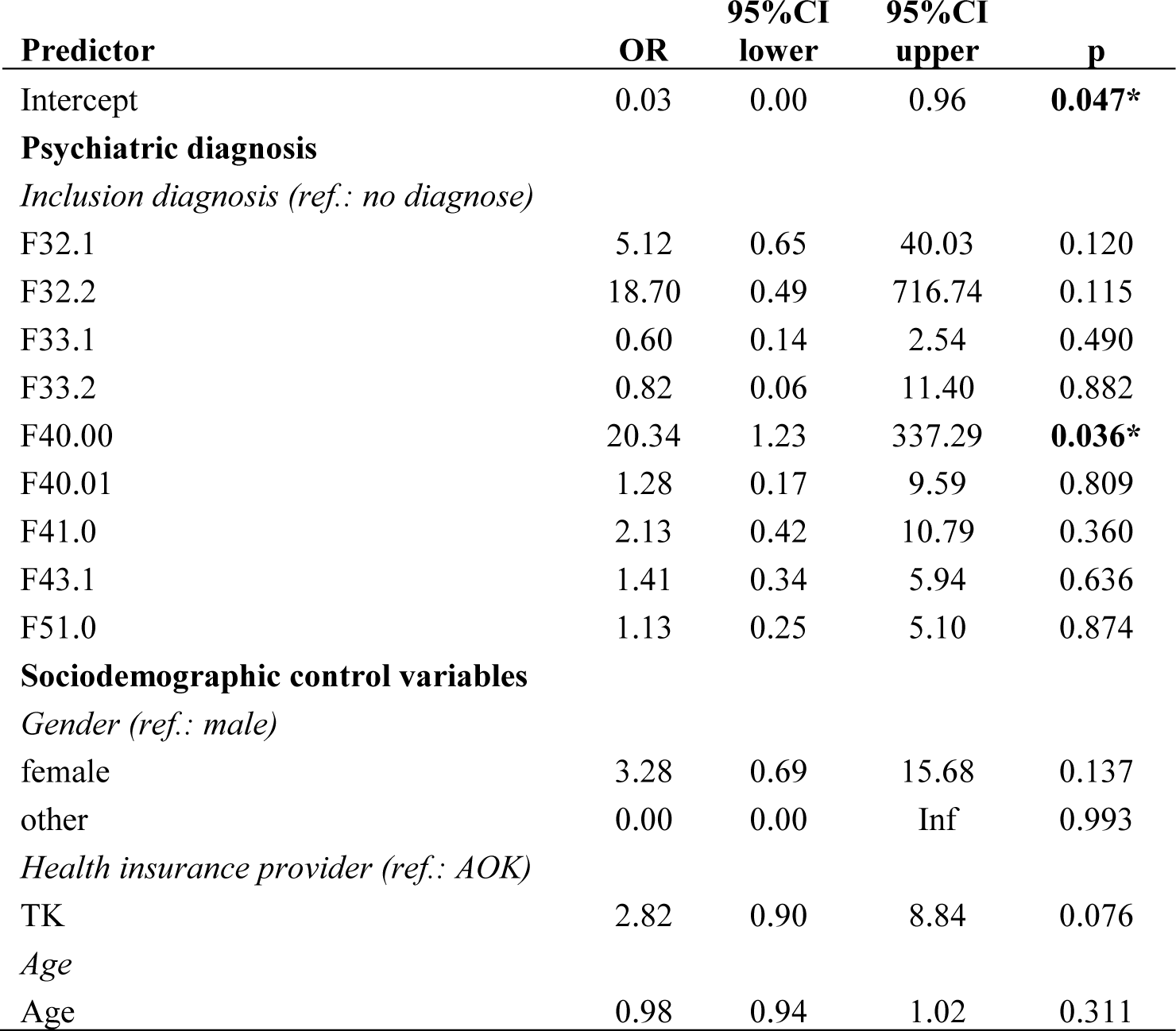
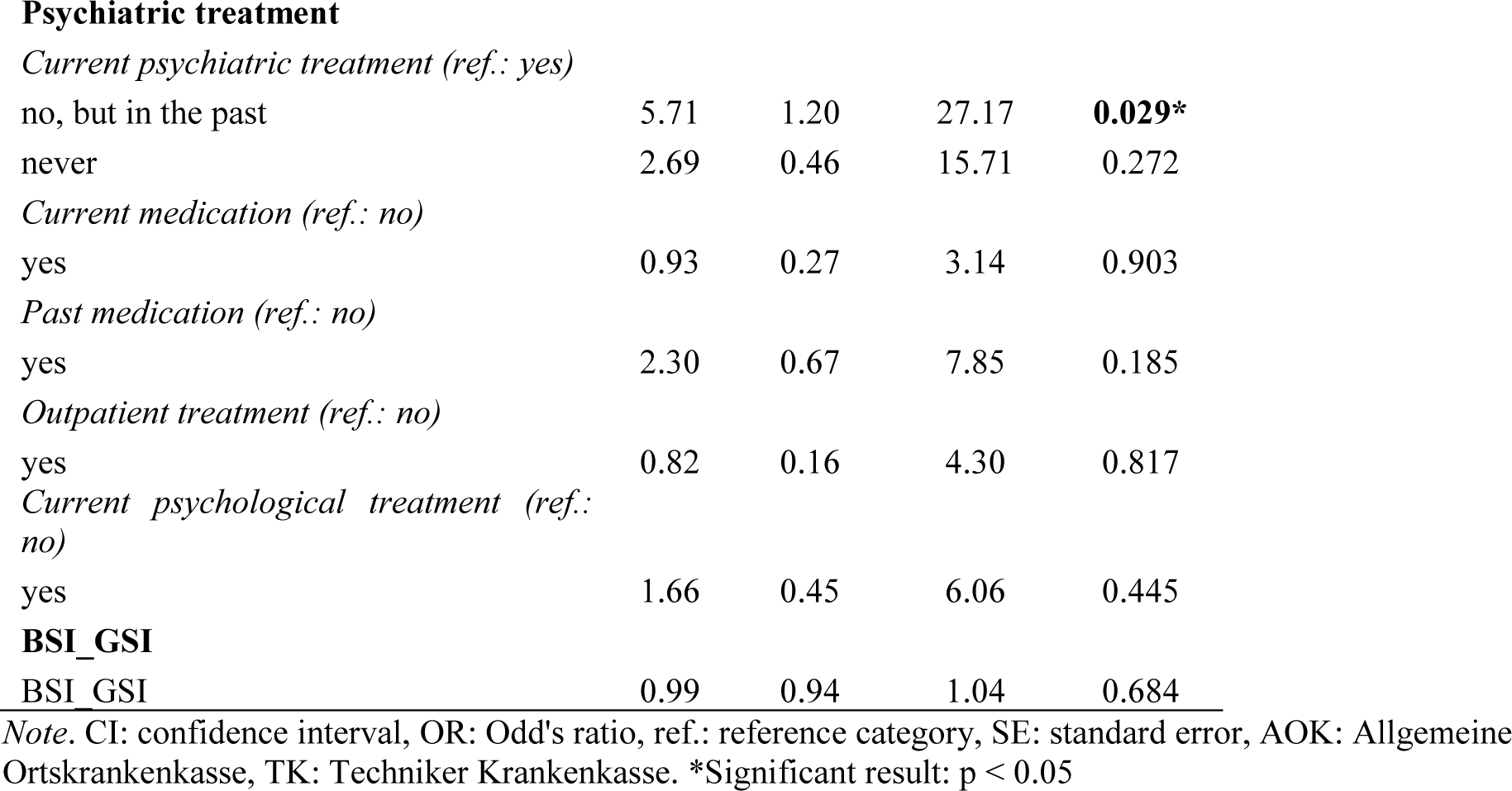
Results of Logistic regression predicting dropout (yes/no) on the ITT sample.

| Predictor | OR | 95%CI<br>lower | 95%CI<br>upper | p |
| --- | --- | --- | --- | --- |
| Intercept | 0.03 | 0.00 | 0.96 | **0.047\*** |
| **Psychiatric diagnosis** |  |  |  |  |
| *Inclusion diagnosis (ref.: no diagnose)* |  |  |  |  |
| F32.1 | 5.12 | 0.65 | 40.03 | 0.120 |
| F32.2 | 18.70 | 0.49 | 716.74 | 0.115 |
| F33.1 | 0.60 | 0.14 | 2.54 | 0.490 |
| F33.2 | 0.82 | 0.06 | 11.40 | 0.882 |
| F40.00 | 20.34 | 1.23 | 337.29 | **0.036\*** |
| F40.01 | 1.28 | 0.17 | 9.59 | 0.809 |
| F41.0 | 2.13 | 0.42 | 10.79 | 0.360 |
| F43.1 | 1.41 | 0.34 | 5.94 | 0.636 |
| F51.0 | 1.13 | 0.25 | 5.10 | 0.874 |
| **Sociodemographic control variables** |  |  |  |  |
| *Gender (ref.: male)* |  |  |  |  |
| female | 3.28 | 0.69 | 15.68 | 0.137 |
| other | 0.00 | 0.00 | Inf | 0.993 |
| *Health insurance provider (ref.: AOK)* |  |  |  |  |
| TK | 2.82 | 0.90 | 8.84 | 0.076 |
| *Age* |  |  |  |  |
| Age | 0.98 | 0.94 | 1.02 | 0.311 |
| **Psychiatric treatment***Current psychiatric treatment (ref.: yes)* |  |  |  |  |
| no, but in the past | 5.71 | 1.20 | 27.17 | **0.029\*** |
| never | 2.69 | 0.46 | 15.71 | 0.272 |
| *Current medication (ref.: no)* |  |  |  |  |
| yes | 0.93 | 0.27 | 3.14 | 0.903 |
| *Past medication (ref.: no)* |  |  |  |  |
| yes | 2.30 | 0.67 | 7.85 | 0.185 |
| *Outpatient treatment (ref.: no)* |  |  |  |  |
| yes | 0.82 | 0.16 | 4.30 | 0.817 |
| *Current psychological treatment (ref.: no)* |  |  |  |  |
| yes | 1.66 | 0.45 | 6.06 | 0.445 |
| **BSI\_GSI** |  |  |  |  |
| BSI_GSI | 0.99 | 0.94 | 1.04 | 0.684 |

*Note.* CI: confidence interval, OR: Odd's ratio, ref.: reference category, SE: standard error, AOK: Allgemeine Ortskrankenkasse, TK: Techniker Krankenkasse. \*Significant result: $p < 0.05$

## Further results on completer analysis (completer sample)

**Table S21.**
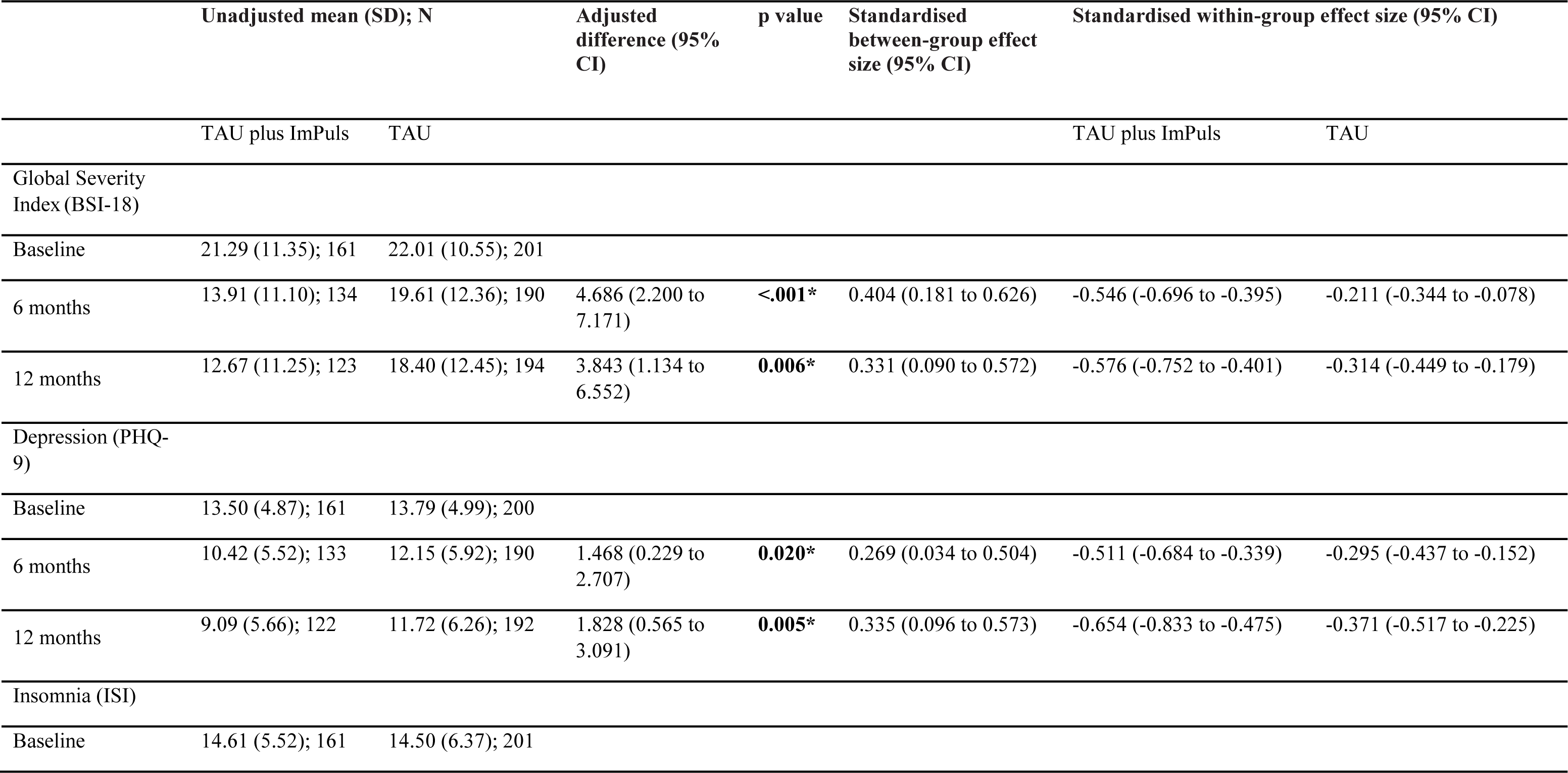
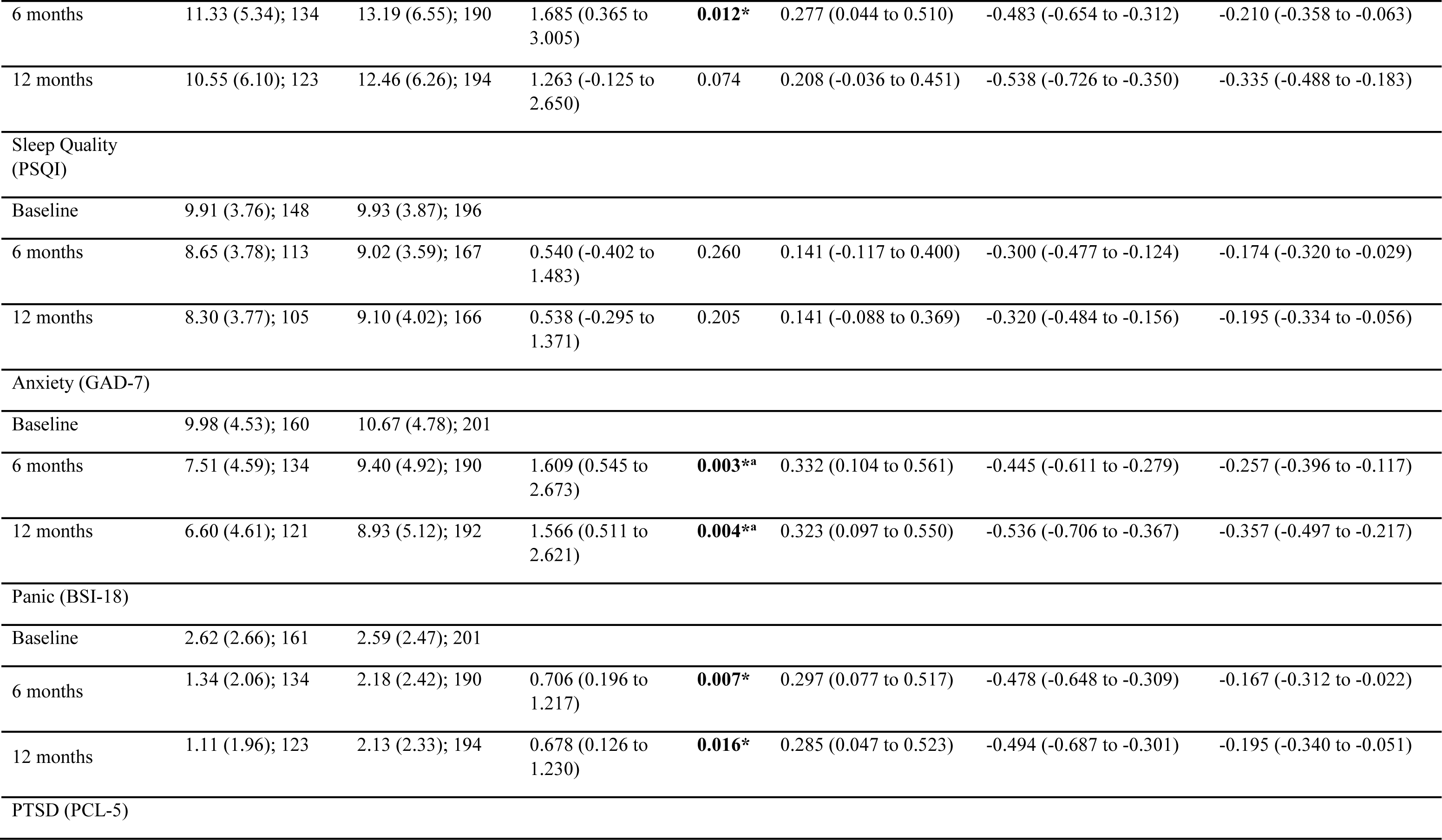
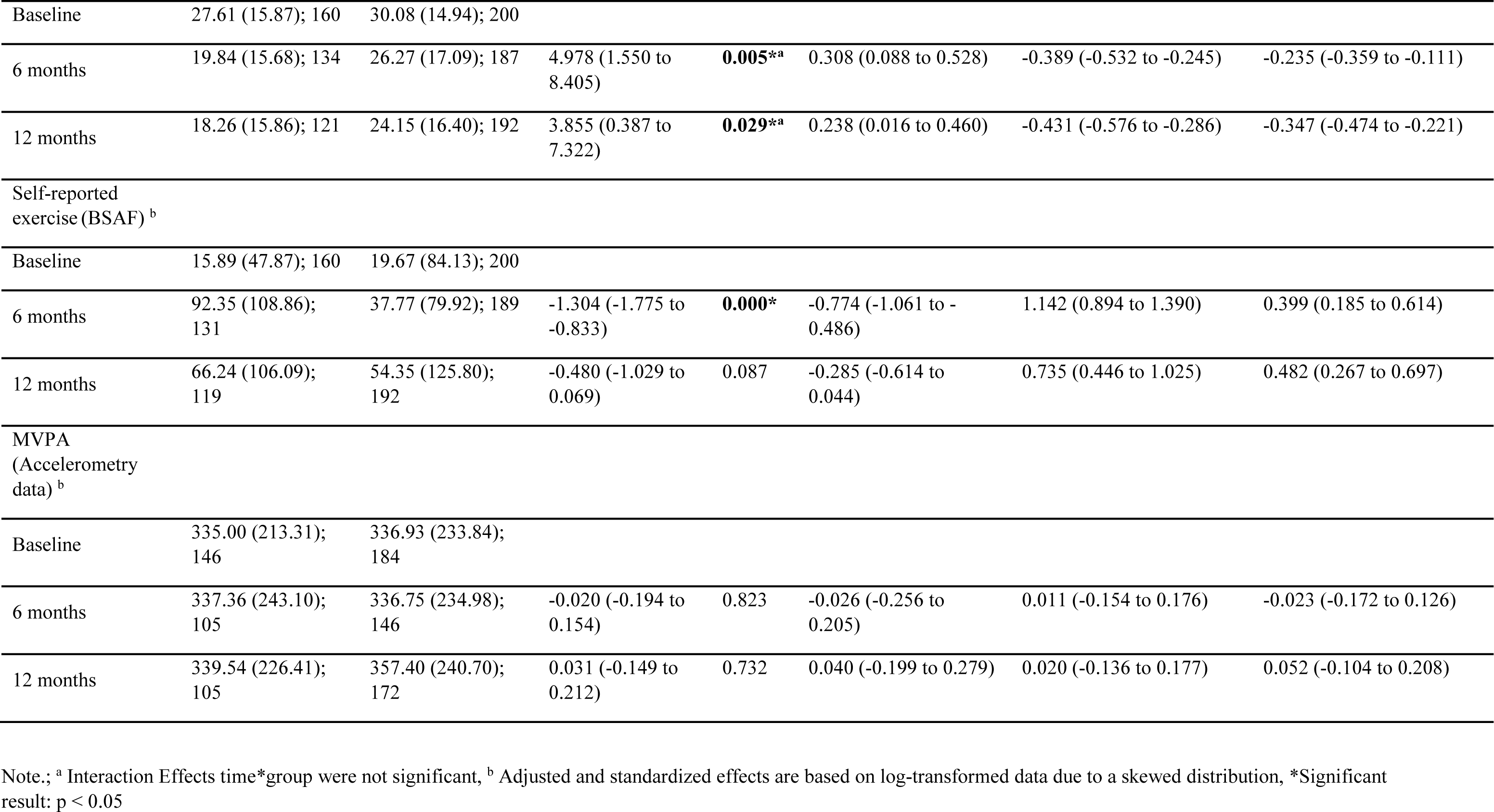
Results of the mixed models on the completer sample including unadjusted means, adjusted differences, standardized between nd within-group differences for the primary and all secondary outcomes.

**Table S22.** Results of mixed model with global symptom severity (BSI-18) as the outcome, based on 10 multiply imputed datasets on the Completer sample.

| Predictor | Estimate | 95% CI | SE | t | p |
| --- | --- | --- | --- | --- | --- |
| Intercept | 22.01 | [20.33, 23.69] | 0.86 | 25.66 | <0.001* |
| Timepoint (*ref. baseline*) |  |  |  |  |  |
| 6-months | -2.45 | [-3.93, -0.97] | 0.76 | -3.24 | **0.001\*** |
| 12-months | -3.65 | [-5.14, -2.16] | 0.76 | -4.79 | <0.001* |
| Group (*ref. TAU*) |  |  |  |  |  |
| ImPuls plus TAU | -0.80 | [-3.21, 1.61] | 1.23 | -0.65 | 0.515 |
| Interaction (*ref. Baseline, TAU*) |  |  |  |  |  |
| 6-months, ImPuls plus TAU | -3.89 | [-6.11, -1.66] | 1.14 | -3.42 | **0.001\*** |
| 12-months, ImPuls plus TAU | -3.04 | [-5.51, -0.58] | 1.26 | -2.42 | **0.016\*** |

Note. CI: confidence interval, ref.: reference category, SE: standard error. \*Significant result: $p < 0.05$

**Table S23.** Results of mixed model with Depression (PHQ-9) as the outcome, based on 10 multiply imputed datasets on the Completer sample.

| Predictor | Estimate | 95% CI | SE | t | p |
| --- | --- | --- | --- | --- | --- |
| Intercept | 13.75 | [12.95, 14.55] | 0.41 | 33.80 | <0.001* |
| Timepoint (*ref. baseline*) |  |  |  |  |  |
| 6-months | -1.61 | [-2.35, -0.87] | 0.38 | -4.25 | <0.001* |
| 12-months | -2.03 | [-2.79, -1.27] | 0.39 | -5.24 | <0.001* |
| Group (*ref. TAU*) |  |  |  |  |  |
| ImPuls plus TAU | -0.28 | [-1.42, 0.85] | 0.58 | -0.49 | 0.623 |
| Interaction (*ref. Baseline, TAU*) |  |  |  |  |  |
| 6-months, ImPuls plus TAU | -1.18 | [-2.36, 0.00] | 0.60 | -1.97 | **0.050\*** |
| 12-months, ImPuls plus TAU | -1.54 | [-2.74, -0.35] | 0.61 | -2.53 | **0.012\*** |

Note. CI: confidence interval, ref.: reference category, SE: standard error. \*Significant result: $p < 0.05$

**Table S24.** Results of mixed model with Insomnia Symptoms (ISI) as the outcome, based on 10 multiply imputed datasets on the Completer sample.

| Predictor | Estimate | 95% CI | SE | t | p |
| --- | --- | --- | --- | --- | --- |
| Intercept | 14.55 | [13.42, 15.68] | 0.58 | 25.15 | <0.001* |
| Timepoint (*ref. baseline*) |  |  |  |  |  |
| 6-months | -1.28 | [-2.11, -0.44] | 0.43 | -3.01 | **0.003\*** |
| 12-months | -2.04 | [-2.89, -1.18] | 0.44 | -4.68 | <0.001* |
| Group (*ref. TAU*) |  |  |  |  |  |
| ImPuls plus TAU | -0.03 | [-1.27, 1.21] | 0.63 | -0.05 | 0.962 |
| Interaction (*ref. Baseline, TAU*) |  |  |  |  |  |
| 6-months, ImPuls plus TAU | -1.65 | [-2.95, -0.36] | 0.66 | -2.51 | **0.012\*** |
| 12-months, ImPuls plus TAU | -1.23 | [-2.59, 0.13] | 0.69 | -1.78 | 0.076 |

Note. CI: confidence interval, ref.: reference category, SE: standard error. \*Significant result: $p < 0.05$

**Table S25.** Results of mixed model with Sleep Quality (PSQI) as the outcome, based on 10 multiply imputed datasets on the Completer sample.

| Predictor | Estimate | 95% CI | SE | t | p |
| --- | --- | --- | --- | --- | --- |
| Intercept | 9.90 | [9.29, 10.51] | 0.31 | 31.96 | **0.001\*** |
| Timepoint (*ref. baseline*) |  |  |  |  |  |
| 6-months | -0.67 | [-1.19, -0.14] | 0.27 | -2.48 | **0.014\*** |
| 12-months | -0.74 | [-1.25, -0.24] | 0.26 | -2.90 | **0.004\*** |
| Group (*ref. TAU*) |  |  |  |  |  |
| ImPuls plus TAU | -0.06 | [-0.87, 0.75] | 0.41 | -0.14 | 0.887 |
| Interaction (*ref. Baseline, TAU*) |  |  |  |  |  |
| 6-months, ImPuls plus TAU | -0.48 | [-1.33, 0.37] | 0.43 | -1.11 | 0.268 |
| 12-months, ImPuls plus TAU | -0.48 | [-1.26, 0.30] | 0.40 | -1.20 | 0.230 |

Note. CI: confidence interval, ref.: reference category, SE: standard error. \*Significant result: $p < 0.05$

**Table S26.** Results of mixed model with general Anxiety (GAD-7) as the outcome, based on 10 multiply imputed datasets on the Completer sample.

| Predictor | Estimate | 95% CI | SE | t | p |
| --- | --- | --- | --- | --- | --- |
| Intercept | 10.66 | [9.95, 11.38] | 0.36 | 29.22 | **<0.001\*** |
| Timepoint (*ref. baseline*) |  |  |  |  |  |
| 6-months | -1.24 | [-1.89, -0.60] | 0.33 | -3.77 | **<0.001\*** |
| 12-months | -1.73 | [-2.37, -1.08] | 0.33 | -5.26 | **<0.001\*** |
| Group (*ref. TAU*) |  |  |  |  |  |
| ImPuls plus TAU | -0.70 | [-1.70, 0.31] | 0.51 | -1.36 | 0.173 |
| Interaction (*ref. Baseline, TAU*) |  |  |  |  |  |
| 6-months, ImPuls plus TAU | -0.91 | [-1.92, 0.09] | 0.51 | -1.78 | 0.076 |
| 12-months, ImPuls plus TAU | -0.87 | [-1.86, 0.12] | 0.51 | -1.72 | 0.086 |

Note. CI: confidence interval, ref.: reference category, SE: standard error. \*Significant result: $p < 0.05$

**Table S27.** Results of mixed model with Panic (BSI-18) as the outcome, based on 10 multiply imputed datasets on the Completer sample.

| Predictor | Estimate | 95% CI | SE | t | p |
| --- | --- | --- | --- | --- | --- |
| Intercept | 2.59 | [2.26, 2.92] | 0.17 | 15.35 | **<0.001\*** |
| Timepoint (*ref. baseline*) |  |  |  |  |  |
| 6-months | -0.40 | [-0.73, -0.06] | 0.17 | -2.32 | **0.020\*** |
| 12-months | -0.46 | [-0.80, -0.13] | 0.17 | -2.72 | **0.007\*** |
| Group (*ref. TAU*) |  |  |  |  |  |
| ImPuls plus TAU | 0.03 | [-0.46, 0.53] | 0.25 | 0.13 | 0.894 |
| Interaction (*ref. Baseline, TAU*) |  |  |  |  |  |
| 6-months, ImPuls plus TAU | -0.74 | [-1.25, -0.23] | 0.26 | -2.83 | **0.005\*** |
| 12-months, ImPuls plus TAU | -0.71 | [-1.27, -0.16] | 0.28 | -2.52 | **0.013\*** |

Note. CI: confidence interval, ref.: reference category, SE: standard error. \*Significant result: $p < 0.05$

**Table S28.** Results of mixed model with PTSD symptoms (PCL-5) as the outcome, based on 10 multiply imputed datasets on the Completer sample.

| Predictor | Estimate | 95% CI | SE | t | p |
| --- | --- | --- | --- | --- | --- |
| Intercept | 30.15 | [27.88, 32.43] | 1.16 | 25.98 | <0.001* |
| Timepoint (*ref. baseline*) |  |  |  |  |  |
| 6-months | -3.80 | [-5.72, -1.88] | 0.98 | -3.88 | <0.001* |
| 12-months | -5.62 | [-7.55, -3.68] | 0.99 | -5.68 | <0.001* |
| Group (*ref. TAU*) |  |  |  |  |  |
| ImPuls plus TAU | -2.50 | [-5.85, 0.86] | 1.71 | -1.46 | 0.145 |
| Interaction (*ref. Baseline, TAU*) |  |  |  |  |  |
| 6-months, ImPuls plus TAU | -2.48 | [-5.39, 0.43] | 1.48 | -1.67 | 0.094 |
| 12-months, ImPuls plus TAU | -1.36 | [-4.30, 1.58] | 1.50 | -0.91 | 0.365 |

Note. CI: confidence interval, ref.: reference category, SE: standard error. \*Significant result: $p < 0.05$

**Table S29.** Results of mixed model with self-reported exercise as the outcome, based on 10 multiply imputed datasets on the Completer sample.

| Predictor | Estimate | 95% CI | SE | t | p |
| --- | --- | --- | --- | --- | --- |
| Intercept | 0.71 | [0.47, 0.96] | 0.13 | 5.68 | <0.001* |
| Timepoint (*ref. baseline*) |  |  |  |  |  |
| 6-months | 0.67 | [0.33, 1.02] | 0.18 | 3.79 | <0.001* |
| 12-months | 0.81 | [0.47, 1.16] | 0.18 | 4.59 | <0.001* |
| Group (*ref. TAU*) |  |  |  |  |  |
| ImPuls plus TAU | 0.05 | [-0.30, 0.40] | 0.18 | 0.29 | 0.769 |
| Interaction (*ref. Baseline, TAU*) |  |  |  |  |  |
| 6-months, ImPuls plus TAU | 1.25 | [0.74, 1.76] | 0.26 | 4.84 | <0.001* |
| 12-months, ImPuls plus TAU | 0.43 | [-0.15, 1.00] | 0.29 | 1.46 | 0.147 |

Note. CI: confidence interval, ref.: reference category, SE: standard error. \*Significant result: $p < 0.05$

**Table S30.** Results of mixed model with accelerometry based moderate to vigorous physical activity (MVPA) as the outcome, based on 10 multiply imputed datasets on the Completer sample.

| Predictor | Estimate | 95% CI | SE | t | p |
| --- | --- | --- | --- | --- | --- |
| Intercept | 5.56 | [5.43, 5.68] | 0.06 | 86.91 | <0.001* |
| Timepoint (*ref. baseline*) |  |  |  |  |  |
| 6-months | -0.02 | [-0.13, 0.09] | 0.06 | -0.32 | 0.753 |
| 12-months | 0.04 | [-0.08, 0.16] | 0.06 | 0.67 | 0.502 |
| Group (*ref. TAU*) |  |  |  |  |  |
| ImPuls plus TAU | -0.01 | [-0.18, 0.16] | 0.09 | -0.08 | 0.936 |
| Interaction (*ref. Baseline, TAU*) |  |  |  |  |  |
| 6-months, ImPuls plus TAU | 0.03 | [-0.14, 0.19] | 0.08 | 0.32 | 0.752 |
| 12-months, ImPuls plus TAU | -0.02 | [-0.19, 0.14] | 0.09 | -0.29 | 0.776 |

Note. CI: confidence interval, ref.: reference category, SE: standard error. \*Significant result: $p < 0.05$

## Mediation Analysis (Completer sample)

**Table S31.** Results of structural equation modeling with bootstrapping (5000 iterations) on the completer sample for model on changes of global symptom severity from baseline to 6 months assessment. Missings were handled with full-information maximum likelihood estimation. The model was saturated and absolute fit indices were not available

| Causal Relationship<sup>a</sup> |  |  | Est | SE | z | p |
| --- | --- | --- | --- | --- | --- | --- |
| *direct effects* |  |  |  |  |  |  |
| Δ GSI | ← | Condition (ImPuls plus TAU vs. TAU) | -0.14 | 0.05 | -2.77 | **0.006\*** |
| Δ GSI | ← | Δ MVPA | -0.03 | 0.07 | -0.49 | 0.623 |
| Δ GSI | ← | Δ self-reported Exercise | -0.13 | 0.06 | -2.24 | **0.025\*** |
| Δ MVPA | ← | Condition (ImPuls plus TAU vs. TAU) | 0.02 | 0.06 | 0.25 | 0.802 |
| Δ self-reported exercise | ← | Condition (ImPuls plus TAU vs. TAU) | 0.31 | 0.05 | 5.95 | **<0.001\*** |
| Δ MVPA | ~~ | Δ self-reported Exercise | 0.12 | 0.07 | 1.69 | 0.092 |
| *indirect effects<sup>b</sup>* |  |  |  |  |  |  |
| Δ GSI | ← | Condition x Δ MVPA | 0.00 | 0.00 | -0.22 | 0.828 |
| Δ GSI | ← | Condition x Δ self-reported Exercise | -0.04 | 0.02 | -2.13 | **0.033\*** |
| *total effect* |  |  | -0.18 | 0.05 | -3.41 | **<0.001** |

Note. <sup>a</sup> = MVPA and self-reported exercise were log-transformed, <sup>b</sup> = We found that the model with the indirect effect of exercise showed a better fit than the model without this effect in terms of AIC. Est = standardized regression coefficient, SE = standard error of the mean, Δ GSI = change in Global Symptom severity from baseline to 6-months, Δ MVPA= change in accelerometry based physical activity, TAU = treatment as usual; \*Significant result: $p < 0.05$

**Table S32.** Results of structural equation modeling with bootstrapping (5000 iterations) on the completer sample for model on changes of global symptom severity from baseline to 12 months assessment. Missings were handled with full-information maximum likelihood estimation. The model was saturated and absolute fit indices were not available

| Causal Relationship* |  |  | Est | SE | z | p |
| --- | --- | --- | --- | --- | --- | --- |
| *direct effects* |  |  |  |  |  |  |
| Δ GSI | ← | Condition (ImPuls plus TAU vs. TAU) | -0.13 | 0.05 | -2.34 | **0.019\*** |
| Δ GSI | ← | Δ MVPA | -0.04 | 0.06 | 0.69 | 0.492 |
| Δ GSI | ← | Δ self-reported Exercise | -0.20 | 0.05 | -4.19 | **<0.001\*** |
| Δ MVPA | ← | Condition (ImPuls plus TAU vs. TAU) | 0.02 | 0.06 | 0.29 | 0.776 |
| Δ self-reported exercise | ← | Condition (ImPuls plus TAU vs. TAU) | 0.11 | 0.06 | 1.85 | **0.065** |
| Δ MVPA | ↔ | Δ self-reported Exercise | 0.07 | 0.06 | 1.03 | 0.303 |
| *indirect effects* |  |  |  |  |  |  |
| Δ GSI | ← | Condition x Δ MVPA | 0.00 | 0.00 | 0.27 | 0.790 |
| Δ GSI | ← | Condition x Δ self-reported Exercise | -0.02 | 0.01 | -1.74 | 0.081 |
| *total effect* |  |  | -0.15 | 0.06 | -2.68 | **0.007\*** |

Note. \* = MVPA and self-reported exercise were log-transformed, Est = standardized regression coefficient, SE = standard error of the mean, Δ GSI = change in Global Symptom severity from baseline to 6-months, Δ MVPA= change in accelerometry based physical activity, TAU = treatment as usual. \*Significant result: $p < 0.05$

## Adverse and Serious Adverse Events

**Table S33.** Detailed information on serious adverse events (SAE)

|  | N SAEs (N participants with SAE) | Severity |  | Duration (days) min-max | ImPuls+TAU<br>N SAEs (N participants with SAE) | TAU<br>N SAEs (N participants with SAE) | Relatedness to intervention<br>N SAEs (N participants with SAE) |
| --- | --- | --- | --- | --- | --- | --- | --- |
|  |  | N SAE moderate | N SAE severe |  |  |  |  |
| Suicide | 0 (0) |  |  |  |  |  |  |
| Suicide attempt | 0 (0) |  |  |  |  |  |  |
| Other event that resulted in death | 0 (0) |  |  |  |  |  |  |
| Intentional serious self-injury resulting in hospitalization | 1 (1) | 0 | 1 | 47 | 0 (0) | 1 (1) | 0 (0) |
| Third party injury | 0 (0) |  |  |  |  |  |  |
| Event that is acutely life-threatening (i.e., study participant is in acute danger of death) | 0 (0) |  |  |  |  |  |  |
| Event that results in significant physical disability | 1 (1) | 1 | 0 | 39 | 1 (1) | 0 (0) | 1 (1) |
| Hospitalization due to psychiatric symptoms | 24 (23) | 2 | 22 | 2-26 | 10 (9) | 14 (14) | 0 (0) |
| Hospitalization due to somatic symptoms | 13 (13) | 4 | 9 | 1-90 | 4 (4) | 9 (9) | 0 (0) |

*Note. SAE = severe adverse event; TAU = Treatment as usual; min = minimum; max = maximum.*

**Table S34.** Detailed information on adverse events (AE)

|  | Number of<br>participants with<br>AE (%<br>*Participants*<br>*with AE*); N<br>Missing | ImPuls+TAU (*n* = 199);<br>Number of participants with<br>AE (% *Participants with*<br>*AE*)<sup>1</sup>;N Missings | TAU (*n* = 201);<br>Number of participants<br>with AE (% *Participants*<br>*with AE*)<sup>2</sup>;N Missings |
| --- | --- | --- | --- |
| Onset of new psychological symptoms | 42 (10.50); 43 | 16 (8.04); 30 | 26 (12.94); 11 |
| Significant worsening of pre-existing psychological symptoms | 71 (17.75); 45 | 23 (11.56); 31 | 48 (23.88); 12 |
| Occurrence of new physical symptoms | 102 (25.50); 44 | 38 (19.10); 30 | 64 (31.84); 12 |
| Significant worsening of pre-existing physical symptoms | 65 (16.25); 44 | 19 (9.55); 31 | 46 (22.89); 11 |
| Injuries that result in not being able to perform any sports activity for at least 2 months | 53 (13.25); 45 | 28 (14.07); 32 | 25 (12.44); 11 |

*Note.* AE = Adverse Event
<sup>1</sup>In total, *n* = 85 participants of the ImPuls+TAU group reported at least one adverse event.
<sup>2</sup>In total, *n* = 135 participants of the TAU group reported at least one adverse event.

**Table S35.** Analysis of group differences in AEs and SAEs between groups.

|  | ImPuls plus Tau | TAU | Group difference | P value |
| --- | --- | --- | --- | --- |
| AE, N (%) | 85 / 136 (62.5) | 135 / 191 (70.7) | OR = 0.69 | 0.12 |
| SAE, M (SD), N | 0.08 (0.28), 198 | 0.12 (0.35), 201 | t(381.1) = 1.36 | 0.18 |

